# The potential global impact and cost-effectiveness of next-generation influenza vaccines: a modelling analysis

**DOI:** 10.1101/2024.09.19.24313950

**Authors:** Lucy Goodfellow, Simon R Procter, Mihaly Koltai, Naomi R Waterlow, Johnny A N Filipe, Carlos K H Wong, Edwin van Leeuwen, WHO Technical Advisory Group for the Full Value of Influenza Vaccines Assessment and project team, Next-generation influenza vaccine impact modelling contributors, Rosalind M Eggo, Mark Jit

**Author notes:** Membership of WHO Technical Advisory Group for the Full Value of Influenza Vaccines Assessment and project team and Next-generation influenza vaccine impact modelling contributors are provided in the Acknowledgements.

## Abstract

**Background:** Next-generation influenza vaccines (NGIVs) are in development and have the potential to achieve greater reductions in influenza burden, with resulting widespread health and economic benefits. Understanding the prices at which their market can be sustained and which vaccination strategies may maximise impact and cost-effectiveness, particularly in low- and middle-income countries, can provide a valuable tool for vaccine development and investment decision-making at a national and global level. To address this evidence gap, we projected the health and economic impact of NGIVs in 186 countries and territories.

**Methods and Findings:** We inferred current influenza transmission parameters from World Health Organization (WHO) FluNet data in regions defined by their transmission dynamics, and projected thirty years of influenza epidemics, accounting for demographic changes. Vaccines considered included current seasonal vaccines, vaccines with increased efficacy, duration, and breadth of protection, and universal vaccines, defined in line with the WHO Preferred Product Characteristics. We estimated cost-effectiveness of different vaccination scenarios using novel estimates of key health outcomes and costs.

NGIVs have the potential to substantially reduce influenza burden: compared to no vaccination, vaccinating 50% of children aged under 18 annually prevented 1.3 (95% uncertainty range (UR): 1.2-1.5) billion infections using current vaccines, 2.6 (95% UR: 2.4-2.9) billion infections using vaccines with improved efficacy or breadth, and 3.0 (95% UR: 2.7-3.3) billion infections using universal vaccines. In many countries, NGIVs were cost-effective at higher prices than typically paid for existing seasonal vaccines. However, cross-subsidy may be necessary for improved vaccines to be cost-effective in lower income countries.

This study is limited by the availability of accurate data on influenza incidence and influenza-associated health outcomes and costs. Furthermore, the model involves simplifying assumptions around vaccination coverage and administration, and does not account for societal costs or budget impact of NGIVs. How NGIVs will compare to the vaccine types considered in this model when developed is unknown. We conducted sensitivity analyses to investigate key model parameters.

**Conclusions:** This study highlights the considerable potential health and economic benefits of NGIVs, but also the variation in cost-effectiveness between high-income and low- and middle-income countries. This work provides a framework for long-term global cost-effectiveness evaluations, and the findings can inform a pathway to developing NGIVs and rolling them out globally.

## Introduction

Globally, seasonal influenza is a substantial cause of respiratory illness, morbidity, and mortality, causing 2911,000-646,1000 deaths annually and significant economic impact through healthcare costs, costs to the individual, and productivity losses [1–3]. The burden varies between countries and wider geographical regions, due to variation in circulating influenza strains and subtypes, population age structure, and current vaccination programmes and coverage. Furthermore, the timing and regularity of influenza epidemics ranges widely around the world, as does the quality and reliability of influenza surveillance data [4].

Seasonal influenza vaccines have been available since the 1940s, and have been subject to extensive improvements and developments since their introduction [5]. However, while influenza vaccines are widely used in the Americas and some high-income countries (HICs), seasonal vaccination coverage remains low globally, and as of 2024 only 34% of low- and lower-middle income countries have a national policy for seasonal influenza vaccination [6].

Current seasonal influenza vaccines face several barriers that can limit their impact and cost-effectiveness. Their duration of protection is less than a year [7], which does not provide immunity through long or multi-peak seasons in temperate and tropical climates, and also requires annual revaccination. Current vaccines must also be reformulated annually based on early estimates of circulating influenza strains and subtypes due to the long timeframe needed for vaccine production. This can lead to very low vaccine effectiveness (VE) in some seasons, particularly in older people for whom vaccines are typically less effective [8]. Next-generation influenza vaccines (NGIVs) are in development which aim to address these limitations, with 40 vaccine candidates currently in clinical trials and over 170 preclinical candidates [9]. The World Health Organization (WHO) defines several types of NGIVs using Preferred Product Characteristics (PPCs); they are categorised as *‘improved’* vaccines, which have increased efficacy or breadth of protection and length of immunity, and *‘universal’* vaccines, which have an increased efficacy and breadth of protection, and immunity lasting up to 5 years [10].

Previous cost-effectiveness analyses conducted in Kenya, UK, and USA have found NGIVs to be cost-effective over a range of willingness-to-pay (WTP) thresholds [11,12]. However, understanding the potential cost-effectiveness of NGIVs globally, and the vaccine prices at which their market can be sustained, is key to informing the planning of possible future investments and decisions made by manufacturers, governments, and potential donors. Here, we expand on models previously used to estimate the national-level cost-effectiveness of NGIVs to generate global estimates.

## Methods

We used a modelling framework consisting of four steps (Figure 1a) to assess the future impact and cost-effectiveness of NGIVs in 186 countries and territories (hereafter referred to as just countries). The steps were: (1) epidemiological inference model (infer current influenza transmission parameters in regions with similar transmission dynamics), (2) vaccination model (project age- and vaccination status-specific populations in each country), (3) epidemic model (simulate future influenza epidemics), and (4) economic model (estimate cost-effectiveness).

**Figure 1.**
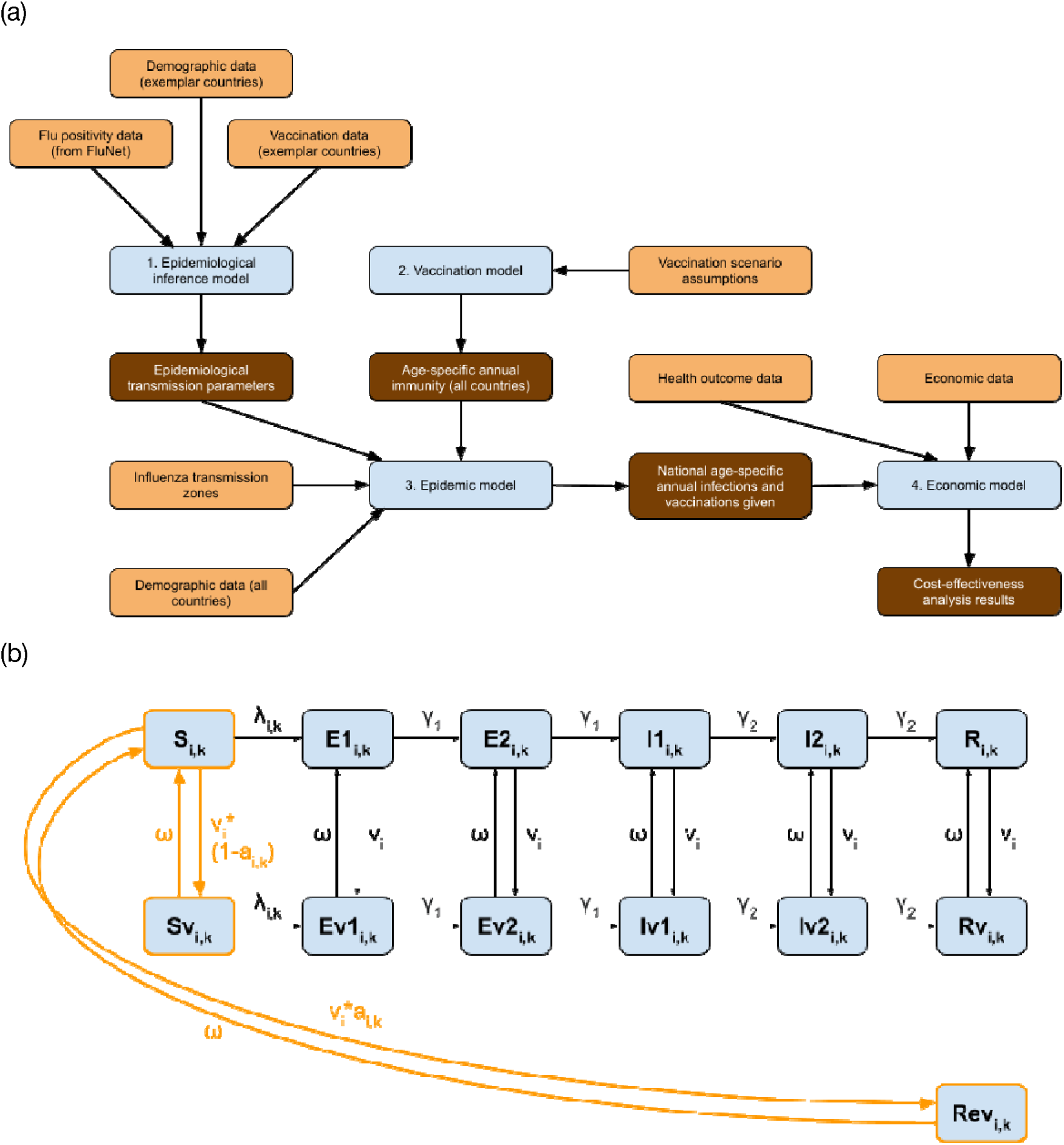
**a)** Overview of modelling steps. Orange indicates inputs, brown indicates outputs and blue shows the modelling elements. **b)** Vaccination and transmission models. Compartments outlined in orange and transitions in solid orange are included in both the vaccination and the transmission models. Transitions in black are only included in the transmission model. **v** denotes the age-specific rates of vaccination, *a* the vaccine effectiveness, which varied by age and strain and depended annually on whether the vaccine matches circulating strains in each hemisphere, and ω vaccine-derived immunity waning. Each compartment was stratified by age (i) and strain (k). Ageing, births, and age-specific mortality are not included in this diagram.

Populations were stratified into four age categories: 0-4, 5-19, 20-64, and 65+ years of age. The age-stratified transmission model used was an extension of the *FluEvidenceSynthesis* model (Figure 1b) [13], and consisted of 13 compartments: Susceptible (S), Exposed (E1, E2), Infectious (I1, I2), and Recovered (R), their ineffectively vaccinated counterparts (Sv-Rv), and Rev (individuals who were vaccinated effectively) (Figure 1b). The E and I populations were split into two sequential compartments to produce gamma-distributed latent and infectious periods. Susceptibles who were infected progressed through the E and I compartments and entered the R compartment after ceasing to be infectious, whereupon they could not be re-infected during the same epidemic. This transmission model was used for both the epidemic inference (Step 1) and epidemic model (Step 3).

### Epidemiological inference model

WHO provides national-level weekly data on laboratory-confirmed influenza through FluNet, an online tool, but the availability and consistency of this data varies widely and could not be used to inform influenza epidemiology in every country [14]. We therefore used a global categorisation of countries with similar influenza epidemiology to project characteristics of influenza transmission inferred for a limited number of countries onto the rest of the world.

We expanded the seven Influenza Transmission Zones (ITZs) produced by Chen et al. [4], which classified 109 countries with data available in FluNet using influenza season timing, laboratory-confirmed influenza positivity, and location parameters. The 77 countries not classified in Chen et al. due to insufficient influenza surveillance data were assigned to an existing ITZ based on location parameters (Supplementary Section 2b). The exemplar countries for each ITZ were chosen to maximise the number of years with available data in FluNet and number of laboratory tests performed: Argentina (Southern America), Australia (Oceania-Melanesia-Polynesia), Canada (Northern America), China (Eastern & Southern-Asia), Ghana (Africa), Turkey (Asia-Europe), United Kingdom (Europe). In each exemplar country, we identified distinct influenza A and influenza B epidemics using weekly laboratory-confirmed influenza incidence in the inference period of 1st January 2010 to 31st December 2019 (Supplementary Section 2d).

In each exemplar country, age-specific seasonal vaccination coverage was assumed to be constant over the 2010-2019 inference period based on estimates from the same time period, with the exception of the UK, where seasonal vaccination policy changed in 2013 (Supplementary Section 3d). We determined whether vaccine strains ‘matched’ or ‘mismatched’ dominant circulating strains of influenza A and B in the Northern and Southern Hemisphere in each year using peer-reviewed literature (Supplementary Section 3e). In line with existing literature, we assumed that in years in which the vaccine strains matched the circulating influenza viruses, VE against infection was 70% in under 65 year olds, and 46% in the age group 65+, compared to 42% and 28%, respectively, in mismatched years [12,15].

We fitted our model to incidence data independently for each identified epidemic in each country using the Markov Chain Monte Carlo (MCMC) algorithm in the *BayesianTools* R package [16], and obtained joint posterior samples of the reporting rate, population susceptibility, transmissibility, and initial number of infections (Supplementary Section 3f).

### Vaccination model

To ascertain the future impact of NGIVs, we used a 30 year simulation period between 1st January 2025 to 31st December 2054. The vaccination model tracked the vaccination status-specific size of each age group over time. Demographic changes (births, mortality, ageing) occurred annually on April 1st (Northern Hemisphere) or October 1st (Southern Hemisphere) using projected 2025 demographic parameters (Supplementary Section 5a). Vaccinations were given at a constant rate over a twelve-week period to accrue to the intended coverage level, beginning on October 1st (Northern Hemisphere) or April 1st (Southern Hemisphere). A proportion of those vaccinated, defined by VE, became immune to infection and entered the Rev compartment; the complement of this proportion did not develop immunity and entered the Sv compartment (Figure 1b). Individuals in Rev moved to S upon the waning of immunity. At the same rate, ineffectively vaccinated individuals returned to their unvaccinated counterpart compartment (i.e. Sv to S, Rv to R).

We considered vaccination scenarios defined by combinations of 5 vaccine types as described by WHO PPCs [10] (Table 1) and 5 age-targeting strategies: ages 0-4, 0-10, 0-17, 65+, and 0-17 combined with 65+. The three *improved* vaccines have increased duration of protection, and efficacy or breadth of protection against strains; *universal* vaccines are enhanced in all aspects. Vaccine doses were distributed at a rate determined by the mean immunity duration of the vaccine type used (i.e. fewer vaccine doses were given annually for vaccine types with longer immunity duration). Vaccine doses were distributed independently of previous vaccination and infection status, but we did not assume any increased protection upon multiple doses (Supplementary Section 6b).

**Table 1.** Vaccine types, based on WHO Preferred Product Characteristics [10]. Some vaccine types (including current) may have ‘mismatched’ seasons where their formulation does not match circulating strains.

| Vaccine type | Current seasonal vaccines | Improved (minimal) | Improved (efficacy) | Improved (breadth) | Universal vaccines |
| --- | --- | --- | --- | --- | --- |
| Mean duration of protection | 6 months | 1 year | 2 years | 3 years | 5 years |
| Vaccine effectiveness (Matched 0-64, 65+/ Mismatched 0-64, 65+) | 0.70, 0.46/<br>0.42, 0.28 | 0.70, 0.46/<br>0.42, 0.28 | 0.90, 0.70/<br>0.70, 0.46 | 0.70, 0.46/<br>0.70, 0.46 | 0.90, 0.70/<br>0.90, 0.70 |
| Mismatched seasons? | Yes | Yes | Yes | No | No |

We assumed that vaccination coverage reached 50% in each age group targeted by vaccination programmes, and conducted sensitivity analyses considering 20% or 70% coverage in each targeted age group. We also ran analyses where only duration of immunity or VE improved in NGIVs, to disentangle the combined effects of NGIVs.

### Epidemic model

In each year of the simulation period, we randomly sampled a year from the inference period, and sampled the susceptibility and transmissibility of all epidemics starting in that year from their joint posterior distributions, to produce a thirty-year period of epidemics occurring with the same frequency and intensity as the inference period in each ITZ. For each year of the simulation period, we also randomly sampled whether formulated vaccines would ‘match’ or ‘mismatch’ circulating strains of influenza A and B in both hemispheres where relevant, using probabilities in alignment with the matching frequencies found in the 2010-19 inference period (Supplementary Section 3e). We simulated epidemics in each of the 186 countries using the transmission model (Figure 1b), the sampled ITZ-specific epidemiological parameters, and the national age- and vaccination status-specific population sizes calculated in the vaccination model. This was repeated 100 times for each vaccine scenario to determine uncertainty in our estimates. Contact patterns for each country were based on those of Prem et al. [17], and reweighted to reflect annual demography (Supplementary Section 5b).

Infected individuals could experience asymptomatic infection, symptomatic but non-fever infection, fever, hospitalisation, and death. Data on seasonal influenza infection-fatality ratios (IFRs), which are highly age- and context-dependent, are sparse. We calculated national age-specific IFR estimates using data on seasonal influenza-associated respiratory deaths [1], and global age-specific infection-hospitalisation ratios (IHRs) using data from Paget et al. [18] (Supplementary Section 7a), and used these estimates to calculate the predicted number of hospitalisations and deaths. We conducted a systematised review to compare our IFR estimates against the limited literature (Supplementary Section 10). There is evidence to support the hypothesis that vaccinated individuals who develop breakthrough infections experience less severe influenza [19,20]; we conducted a sensitivity analysis in which breakthrough infections experienced a 50% reduction in both IHR and IFR, and another in which the infectiousness of individuals experiencing breakthrough infections was assumed to be 50% lower.

### Economic model

To estimate the cost-effectiveness of each vaccination scenario, we overlaid a decision tree model onto the underlying dynamic transmission model (Supplementary Figure S28) and a no-vaccination scenario as the comparator, as national-level data on current seasonal vaccination coverage is sparse and vaccination coverage is low in most of the global population. We calculated the disability-adjusted life years (DALYs) averted by estimating age-specific Years of Life Lost (YLLs) per influenza death using national life tables and combining this with Years Lived with Disability (YLDs) for symptomatic cases, fevers, and hospitalisations (Supplementary Section 7a). Future DALYs were discounted at a rate of 3%, and in a sensitivity analysis reduced to 0%, as recommended by WHO [21].

We estimated costs from a healthcare-payer perspective (Supplementary Section 7b). We estimated national costs of hospitalised cases using data from existing systematic reviews in a regression model predicted by national gross domestic product (GDP) per capita, and included the cost of outpatient visits in a sensitivity analysis. Country-level costs of vaccine dose delivery were estimated using data from a meta-regression for low- and middle-income countries [22], and extrapolated to HICs using a regression against healthcare expenditure per capita. As a sensitivity analysis, we also estimated the productivity loss of influenza deaths using a human capital approach. Future costs were discounted at a rate of 3%, and all costs were expressed in 2022 USD.

To inform the potential return on investment to NGIVs developers, we calculated threshold prices per dose for each country below which vaccination would be cost-effective. We used WTP thresholds estimated in Pichon-Riviere et al. [23], which are based on national per-capita health expenditures and life expectancy, and conducted a sensitivity analysis using WTP thresholds of 50% of GDP per capita.

## Results

The expanded ITZs and selected exemplar countries are shown in Figure 2a. In most exemplar countries, observed epidemics followed regular seasonality, but the timing of outbreaks was less regular in Ghana and China (the African and Eastern and Southern Asian ITZs, respectively; Figure 2b). Posterior estimates of susceptibility and transmissibility inferred for each epidemic were in similar ranges, and mean predicted reported cases were highly similar to the data (R^2^=0.96) (Supplementary Section 4).

**Figure 2.**
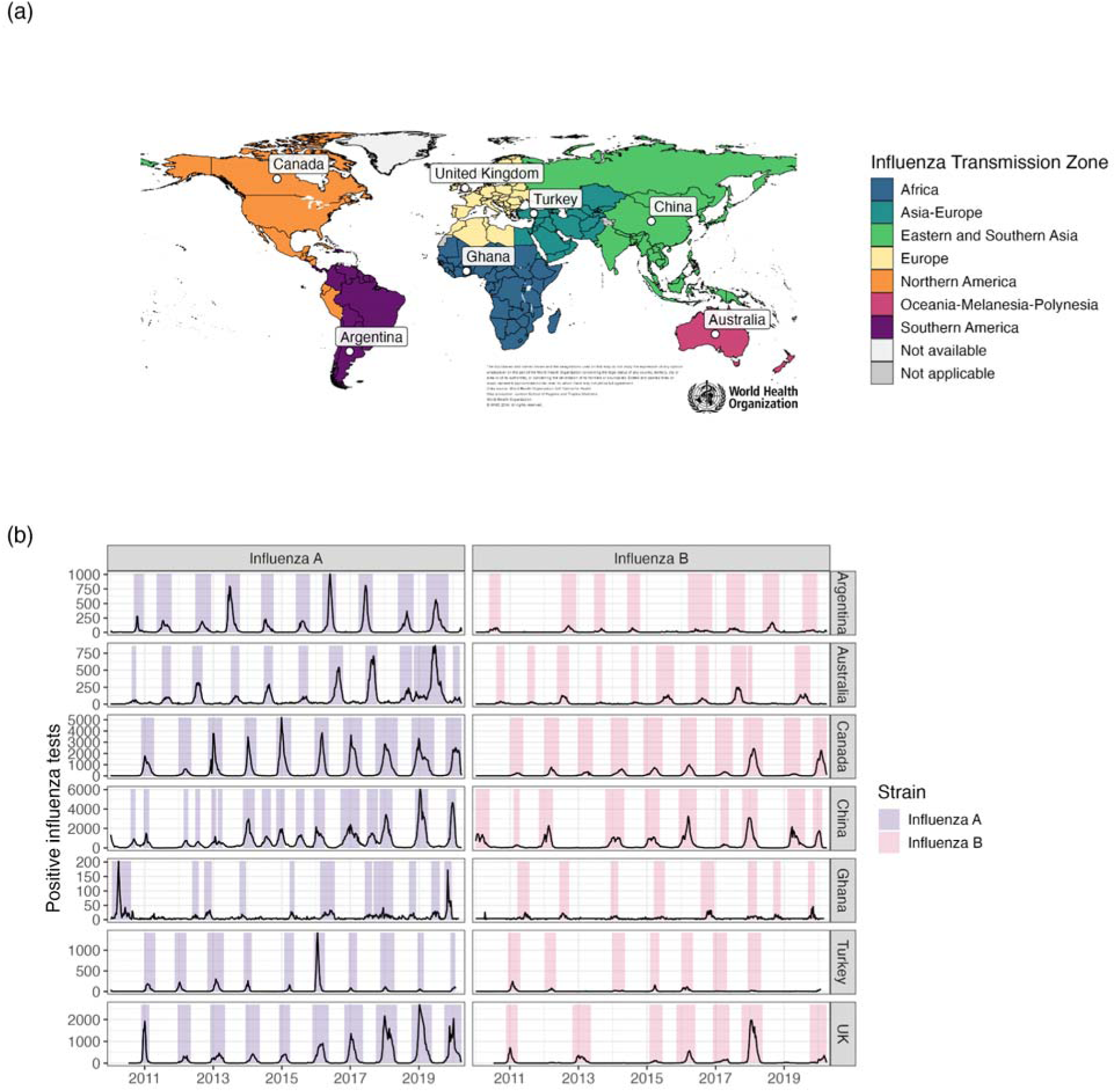
**a)** Map of influenza transmission zones. White dots show exemplar countries for each influenza transmission zone. **b)** FluNet data in each exemplar country over the inference period, stratified by influenza strain, showing total number of positive tests. Shaded time periods indicate identified epidemics. Map base layer from https://gis-who.hub.arcgis.com/pages/detailedboundary.

Globally, the number of influenza infections averted depended on the vaccine characteristics and age-targeting strategies. *Current* vaccines would prevent 1.33 (95% uncertainty range (UR): 1.20-1.48) billion, or 37% of, infections annually when vaccinating 50% of 0-17 year olds worldwide compared to no vaccinations, but only 117 (95% UR: 105-129) million, or 3% of, infections when targeting the 65+ age group. The number of infections averted increased for NGIVs, with *improved (minimal)* preventing 1.93 (95% UR: 1.72-2.11) billion and *improved (efficacy)* and *improved (breadth)* preventing 2.65 and 2.64 (95% UR: 2.39-2.93) billion annual infections respectively when targeting 0-17 year olds, while *universal* vaccines prevented 2.96 (95% UR: 2.70-3.27) billion, or 83% of, infections annually. See Table S8 for annual influenza infections averted under each vaccination scenario.

Some age-targeting strategies were clearly more effective than others: while vaccinating children aged 0-10 required approximately the same number of vaccine doses as vaccinating adults aged over 65, the former strategy prevented up to 9.5x as many infections and 2.5x as many deaths. This is likely because young children have higher contact rates, and so preventing infections in children can lead to highly effective indirect protection for unvaccinated individuals. The global number needed to vaccinate (NNV) to avert one DALY was consistently lowest in the 0-10 age-targeting strategy, and highest in the 65+ age-targeting strategy (Figure 3a), similarly for NNV against infections, hospitalisations, and deaths (Figure S33). While most infections were prevented in the 20-64 age group, averted hospitalisations were concentrated in children under age 5 and adults aged 65+, and fatalities in the 65+ age group (Figure 3b).

**Figure 3:**
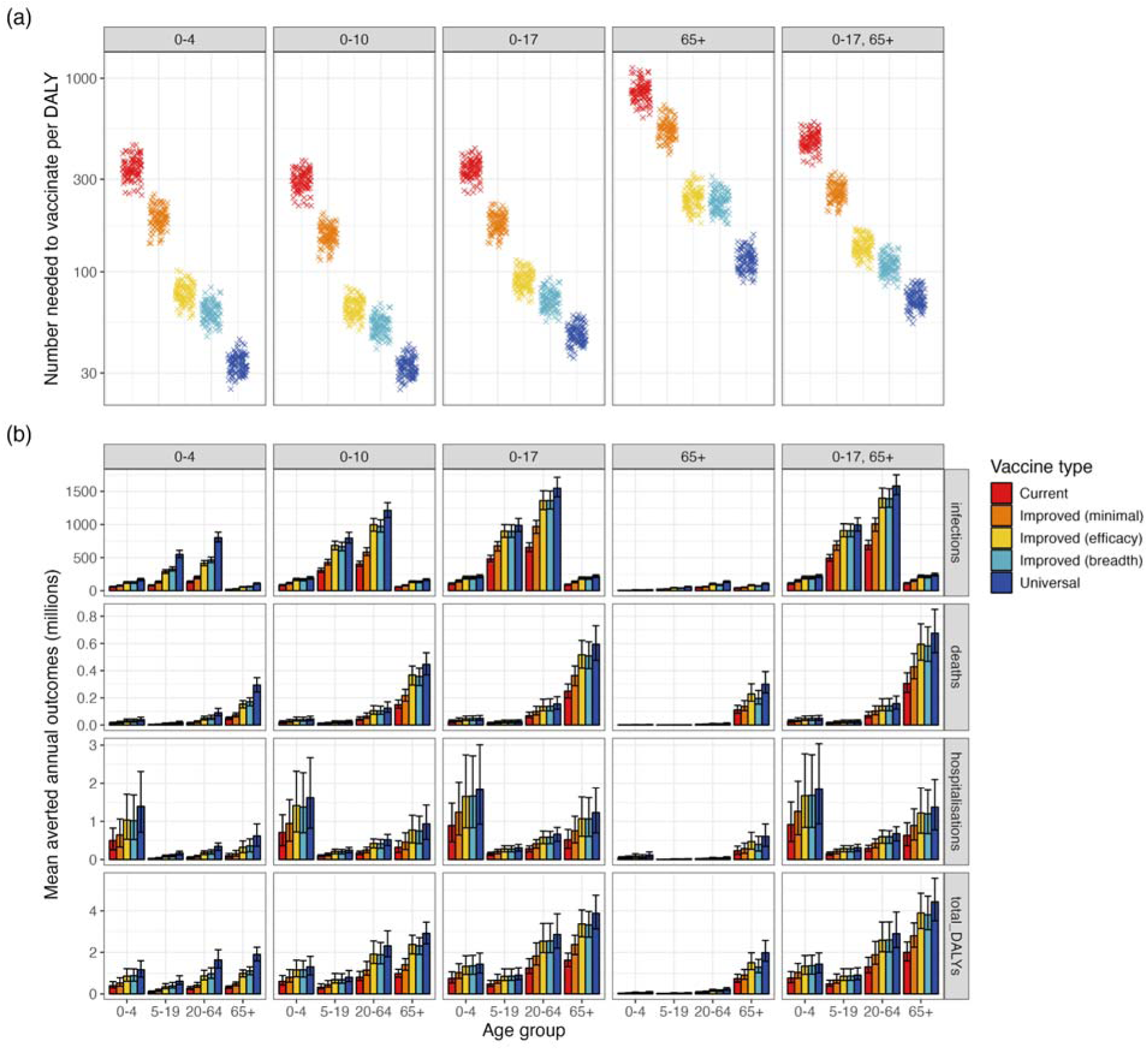
**a)** Global number needed to vaccinate to avert one DALY, for each vaccine type and age-targeting strategy, on a log scale. **b)** Global averted annual age-specific health outcomes under each age-targeting strategy and vaccine type, with 95% uncertainty ranges.

Under no vaccinations, we estimated the average annual number of hospitalisations and deaths between 2025-2054 to be 4.83 (95% UR: 2.82-7.34) million and 1.06 (95% UR: 0.80-1.42) million, respectively. This figure is higher than current estimates [1], due to the assumption of no vaccinations and since populations are predicted to grow and age in the next 30 years. Vaccinating all children under age 18 with current vaccines prevented 1.85 (95% UR: 1.06-2.82) million annual hospitalisations and 357,000 (95% UR: 279,000, 454,000) annual deaths compared to no vaccinations; these figures increased to 2.63 (95% UR: 1.53-3.95) million and 519,000 (95% UR: 401,000-653,000) under i*mproved (minimal)* vaccines, and 4.04 (95% UR: 2.33-6.12) million and 826,000 (95% UR: 641,000-1,050,000) under *universal* vaccines.

Threshold prices below which vaccines are cost-effective tended to increase with national-level income, but varied between countries with similar GDP per capita, even within the same ITZ, indicating that willingness to pay for NGIVs also depends on epidemiology and demography (Figure 4a). NGIVs were associated with higher threshold prices than current seasonal influenza vaccines under all age-targeting strategies in all World Bank income groups (Figure 4b). The 0-10 age-targeting strategy was associated with the highest threshold price across vaccine types in the majority of countries (Figure S42).

**Figure 4.**
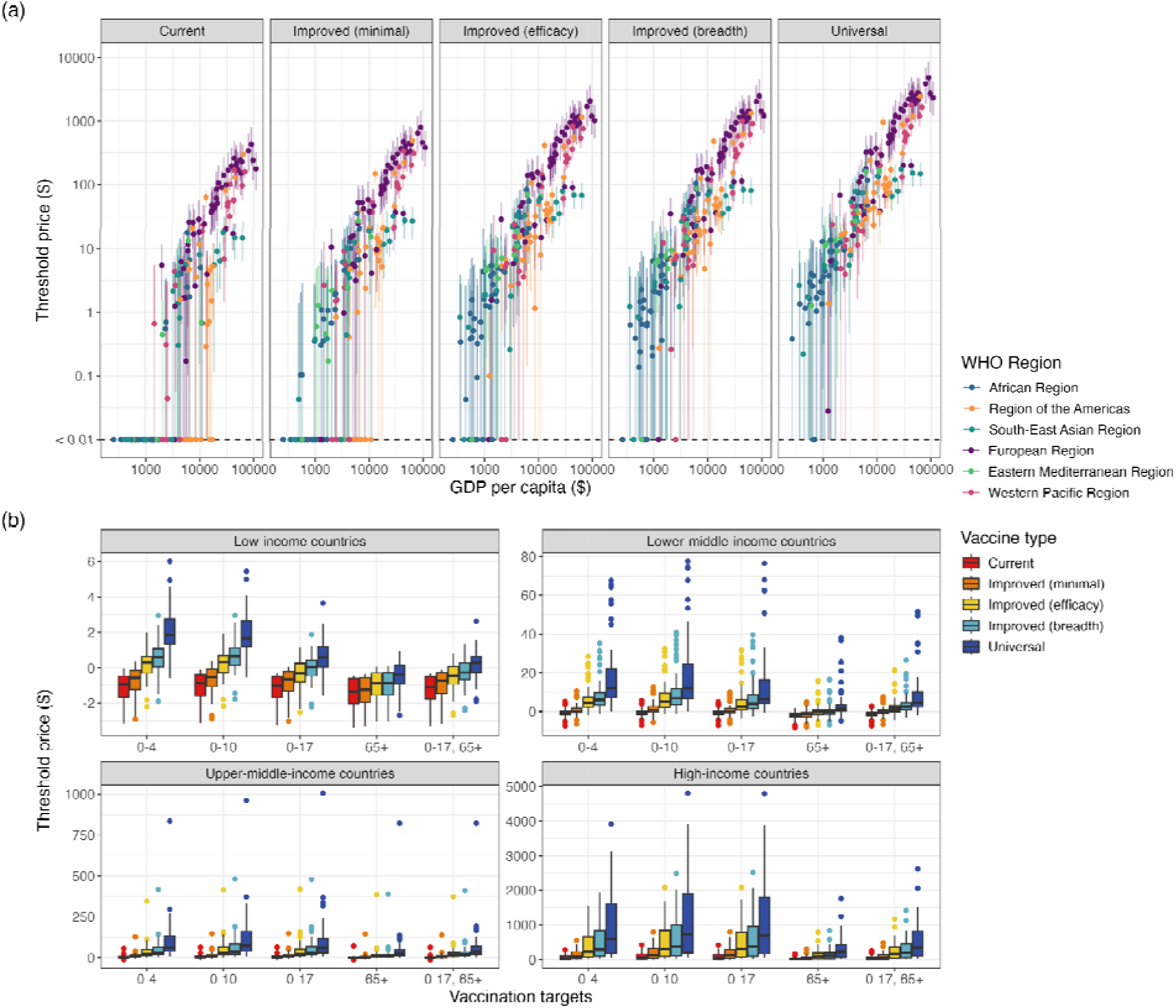
**a)** Median national threshold prices per vaccine dose and 95% uncertainty ranges for each vaccine type when vaccinating 50% of those aged 0-10, on a log-log scale. **b)** Median national threshold vaccine price in each World Bank income group, for each vaccine type and age-targeting strategy.

We found that *current* seasonal vaccines were not cost-effective in any low-income countries (LICs) under any age-targeting strategy or price, and only had positive threshold prices in 31% of lower-middle-income countries (LMICs) when vaccinating all children aged 0-10 (Table S12). In comparison, vaccinating the same age group with *current* seasonal vaccines had positive threshold prices in 98% of HICs, and reached up to $430 (95% UR: $200-$810). *Improved (minimal, efficacy, breadth)* vaccines had feasible threshold prices in very few LICs, but were associated with median threshold prices of up to $12, $32, and $41, respectively, in LMICs when vaccinating children aged 0-10. These vaccines could therefore be cost-effective in many countries, but are unlikely to be cost-effective in LICs without substantial subsidy, as the unit price of newly introduced NGIVs may be higher than the estimated threshold prices.

*Universal* vaccines had positive threshold prices in the majority of countries (184/186) under the 0-10 age-targeting strategy. Median threshold prices ranged up to $5.50 in LICs and $78 in LMICs, ranged between $7.90 and $960 in upper-middle-income countries (UMICs), and between $65 and $4800 in HICs. It is therefore likely that *universal* vaccines will be highly cost-effective in HICs, many UMICs and LMICs, but few LICs without substantial subsidy.

We conducted a range of sensitivity analyses (Supplementary Section 9). More infections, hospitalisations, and deaths were prevented when 70% coverage was achieved in targeted age groups, instead of 50%, similarly less for 20%, but increasing vaccination coverage from 50% to 70% had a diminishing marginal impact of each vaccine dose in terms of reduction of infection (Table S14). Reduced relative infectiousness or disease modifying in breakthrough infections was associated with a small further reduction in the number of infections over the thirty-year period, although the impact on threshold prices for each vaccine type was small. When comparing the effects of increasing VE or the length of immunity provided, more benefits were found to be due to increased length of immunity. In a sensitivity analysis, we estimated the productivity gains from NGIVs, which demonstrate the potential additional economic benefit of NGIVs from a societal perspective (Supplementary Section 9g).

## Discussion

We found that using NGIVs could have a dramatic impact on global influenza burden and be cost-effective in many parts of the world even if prices are high, however affordability is likely to be a barrier to adoption in lower income countries. Vaccinating children aged under 18 years old with currently licensed vaccines could prevent 37% of influenza infections (1.33 billion infections) when compared to no influenza vaccinations; this increased to 53% using *minimally improved* vaccines and 83% using *universal* vaccines. However, for all vaccine types, we found less impact per dose in extending coverage above age ten.

The unit price at which NGIVs could be cost-effective varied widely. In many countries, NGIVs are likely to be cost-effective if they were to become available at prices similar to or higher than other recently introduced vaccines [24]. *Universal* influenza vaccines could become one of the highest value vaccines available in some HICs, with threshold prices reaching thousands of dollars. Conversely, in some LICs, only slightly *improved* vaccines might not be cost-effective from a health-service perspective even if donated for free, and *universal* vaccines would not be cost-effective in any LICs if they were priced at over $6. Our findings highlight the likely need for financial subsidies for procurement and delivery (e.g. through aggressively tiered prices or support from Gavi, the Vaccine Alliance) for such vaccines to be accessible globally. These results are consistent with previous country-level analyses’ findings that *universal* vaccines would likely be cost-effective in the UK, and in Kenya if priced less than $4.94 per dose using a WTP threshold of 45% of GDP per capita [11,12].

We developed novel approaches to simulating future influenza epidemics globally, which allowed us to account for the impact of future demographic changes. A limitation of our data sources was that the model could only be fitted using ten years of influenza data, was subject to simplifying assumptions such as age-consistent reporting rates, and assumed broadly consistent epidemiology across wide regions of the world. We also did not capture within-country variation in vaccine policy and epidemiology, which may be important in geographically large countries such as Canada and China.

The vaccine types considered in this study were guided by WHO PPCs, which are based on expert opinion from 2017 but may not reflect the current state of vaccine development. Using no vaccinations as a comparator scenario overestimates NGIV cost-effectiveness in settings where current seasonal vaccination coverage is high, but these countries make up the minority of the global population as coverage is globally low. Epidemic inference in exemplar countries where coverage is high may have overlooked epidemics that would have occurred without any vaccinations; this effect is likely small, as we observed relatively consistent seasonality in these countries over the inference period.

Many of our simplifying assumptions cause the cost-effectiveness of NGIVs to be underestimated. The assumption that vaccine doses were delivered independently of vaccination and infection history could lead to underestimation of the benefits of NGIVs, since doses could be targeted at individuals with the longest interval since their last dose. Administering vaccines with longer duration of protection is likely to differ from current seasonal vaccination programmes, as populations could receive vaccinations all year round, as opposed to in a pre-epidemic period, or as part of a routine immunisation program, which could lead to further cost-saving. Fixed seasonal timing for vaccination programs may have a diminished impact in subtropical and equatorial countries with multiple epidemic peaks or undefined influenza seasons, particularly for current seasonal vaccines where immunity wanes during the year; previous research has found that while there is no optimal vaccination timing in no-seasonality settings, the timings chosen in this study closely reflect optimal programs in subtropical and temperate settings [25]. Vaccine wastage in the delivery process may also be lower for NGIVs, which we did not consider in this analysis. We did not consider potential future changes in the prevalence of chronic diseases which may be exacerbated by, or exacerbate, influenza burden. Influenza-associated mortality data used in this study only does not account for non-respiratory deaths, for which data is sparse, particularly in LMICs; further discussion of the limitations of this data can be found in [1]. Our estimated threshold prices were influenced by assumptions about the willingness to pay for improvements in health, for which we have used empirical estimates of the opportunity cost of alternative uses of the healthcare budget [23]. The analysis was performed using a healthcare-payer perspective, which does not account for wider economic costs such as out-of-pocket healthcare payments, time spent on informal care-giving, and lost income and improved productivity. Conversely, depending on the market price, there could be a substantial budget impact of NGIVs, particularly if they were to lead to a large expansion in existing influenza vaccine coverage, potentially decreasing cost-effectiveness.

In conclusion, NGIVs have the potential to significantly improve global health if made widely available, and could command higher prices than current seasonal vaccines in many countries, due to their higher VE and reduced need for re-vaccination. Given the high prices achievable in HICs, there may be potential for cross-subsidy in the vaccine market to enhance affordability in LICs and LMICs, for example via aggressively tiered pricing and pooled procurement schemes. While these NGIVs are not yet available, our findings have also shown the health and economic benefits of currently licensed seasonal influenza vaccines in many countries when targeted at children and adolescents.

## Abbreviations

DALY: Disability-Adjusted Life Years
GDP: Gross Domestic Product
HIC: High-Income Country
IFR: Infection-Fatality Ratio
IHR: Infection-Hospitalisation Ratio
ITZ: Influenza Transmission Zone
LIC: Low-Income Country
LMIC: Lower-Middle-Income Country
MCMC: Markov Chain Monte Carlo
NGIV: Next-Generation Influenza Vaccine
NNV: Number Needed to Vaccinate
PPC: Preferred Product Characteristic
UMIC: Upper-Middle-Income Country
UR: Uncertainty Range
VE: Vaccine Effectiveness
WHO: World Health Organization
WTP: Willingness-To-Pay
YLL: Year of life lost

## Data Availability Statement

All analysis code is available at https://github.com/lucy-gf/flu_model_LG. All analyses were conducted using R (version 4.3.1) and RStudio (version 2024.04.1+748) software.

## Acknowledgements

LG, SRP, NRW, RME and MJ were funded through the Task Force for Global Health in collaboration with Partnership for Influenza Vaccine Introduction (PIVI), Ready2Respond, Wellcome Trust, Centers for Disease Control and Prevention (CDC), and by the World Health Organization. JF and CW were funded by AIR@InnoHK administered by Innovation and Technology Commission, Government of Hong Kong Special Administrative Region, China, as part of the Laboratory of Data Discovery for Health (D^2^4H).

EvL, RME, and MJ were also supported by the National Institute for Health Research (NIHR) Health Protection Research Unit (HPRU) in Modelling and Health Economics, a partnership between UK HSA, Imperial College London, and LSHTM (grant number NIHR200908). The views expressed are those of the authors and not necessarily those of the UK Department of Health and Social Care (DHSC), NIHR, or UKHSA. The funders had no role in study design, data collection and analysis, decision to publish, or preparation of the manuscript.

WHO Technical Advisory Group for the Full Value of Influenza Vaccines Assessment and project team:

*<u>WHO FVIVA Technical Advisory Group members:</u>*
*Jon Abramson, Salah Al Awaidy, Silvia Bino, Rebecca Jane Cox, Luzhao Feng, Jodie McVernon, Harish Nair, Anthony T Newall, Punnee Pitisuttithum*

*<u>WHO FVIVA project team members:</u>*
*Philipp Lambach, Mitsuki Koh, Joseph Bresee, Stefano Malvolti, Carsten Mantel, Sara Sá Silva, Adam Soble, Carlo Federici*

Next-generation influenza vaccine impact modelling contributors:

Paula Barbosa, Shawn Gilchrist, Dafrossa Lyimo, Rajinder Suri, Joseph T Wu

We also thank Eduardo Azziz-Baumgartner and Kathryn Lafond for helpful discussions.

## Author Contributions

SRP, NRW, MK, RME, and MJ conceived the study. LG, SRP, MK, NRW, EvL, RME, and MJ designed the methodology. LG, SRP, and MK analysed the data. LG and SRP built the model. LG made the figures. CW and JF conducted the systematised review. LG and SRP wrote the original draft. All authors interpreted the results, contributed to reviewing and editing of the manuscript and have approved the final version.

## Supporting Information legends

**Table S1:** Model parameters, used in the epidemic inference, vaccination, and epidemic models (steps 1-3).

**Figure S1:** Geographical distribution of the seven ITZs produced by Chen et al. [5]. Map base layer from https://gis-who.hub.arcgis.com/pages/detailedboundary.

**Figure S2: (a)** The longitude and latitude of the capital cities of each country in the ITZs, and each ITZ’s cluster centroid (marked as X). **(b)** World map of all countries included in this analysis. Map base layer from https://gis-who.hub.arcgis.com/pages/detailedboundary.

**Table S2:** Influenza Transmission Zone assignments of 186 countries, assigned either by Chen et al. [5] or added based on geographical parameters.

**Table S3:** Summary of FluNet data for each of the chosen exemplar countries between January 2010 and December 2019.

**Figure S3:** Epidemic model for inference, with no underlying vaccination model. Vaccinated individuals were assigned Rev with probability equal to vaccine efficiency.

**Table S4:** Vaccination coverage levels used for inference in exemplar countries between 2010 and 2019 in each of the model age groups.

**Table S5:** Matching (M) and mismatched (U) vaccinations in each year of the inference period, for influenza A and B, in both hemispheres.

**Figure S4:** Posterior distributions of population-level susceptibility and influenza transmissibility in each epidemic used for inference.

**Figure S5:** Validation of inference model’s goodness of fit, comparing reported influenza cases and mean predicted influenza cases across all epidemics, stratified by influenza strains and using log scales on both axes. Dotted line indicates x=y.

**Figure S6: (a)** Posterior distributions of the initial number of infections, reporting rate, population-level susceptibility, and influenza transmissibility in each epidemic used for inference (Argentina, Influenza A). **(b)** Model fits using parameter posteriors.

**Figure S7: (a)** Posterior distributions of the initial number of infections, reporting rate, population-level susceptibility, and influenza transmissibility in each epidemic used for inference (Argentina, Influenza B). **(b)** Model fits using parameter posteriors.

**Figure S8: (a)** Posterior distributions of the initial number of infections, reporting rate, population-level susceptibility, and influenza transmissibility in each epidemic used for inference (Australia, Influenza A). **(b)** Model fits using parameter posteriors.

**Figure S9: (a)** Posterior distributions of the initial number of infections, reporting rate, population-level susceptibility, and influenza transmissibility in each epidemic used for inference (Australia, Influenza B). **(b)** Model fits using parameter posteriors.

**Figure S10: (a)** Posterior distributions of the initial number of infections, reporting rate, population-level susceptibility, and influenza transmissibility in each epidemic used for inference (Canada, Influenza A). **(b)** Model fits using parameter posteriors.

**Figure S11: (a)** Posterior distributions of the initial number of infections, reporting rate, population-level susceptibility, and influenza transmissibility in each epidemic used for inference (Canada, Influenza B). **(b)** Model fits using parameter posteriors.

**Figure S12: (a)** Posterior distributions of the initial number of infections, reporting rate, population-level susceptibility, and influenza transmissibility in each epidemic used for inference (China, Influenza A). **(b)** Model fits using parameter posteriors.

**Figure S13: (a)** Posterior distributions of the initial number of infections, reporting rate, population-level susceptibility, and influenza transmissibility in each epidemic used for inference (China, Influenza B). **(b)** Model fits using parameter posteriors.

**Figure S14: (a)** Posterior distributions of the initial number of infections, reporting rate, population-level susceptibility, and influenza transmissibility in each epidemic used for inference (Ghana, Influenza A). **(b)** Model fits using parameter posteriors.

**Figure S15: (a)** Posterior distributions of the initial number of infections, reporting rate, population-level susceptibility, and influenza transmissibility in each epidemic used for inference (Ghana, Influenza B). **(b)** Model fits using parameter posteriors.

**Figure S16: (a)** Posterior distributions of the initial number of infections, reporting rate, population-level susceptibility, and influenza transmissibility in each epidemic used for inference (Turkey, Influenza A). **(b)** Model fits using parameter posteriors.

**Figure S17: (a)** Posterior distributions of the initial number of infections, reporting rate, population-level susceptibility, and influenza transmissibility in each epidemic used for inference (Turkey, Influenza B). **(b)** Model fits using parameter posteriors.

**Figure S18: (a)** Posterior distributions of the initial number of infections, reporting rate, population-level susceptibility, and influenza transmissibility in each epidemic used for inference (United Kingdom, Influenza A). **(b)** Model fits using parameter posteriors.

**Figure S19: (a)** Posterior distributions of the initial number of infections, reporting rate, population-level susceptibility, and influenza transmissibility in each epidemic used for inference (United Kingdom, Influenza B). **(b)** Model fits using parameter posteriors.

**Figure S20:** The vaccination model, example shown for the first two years of the simulation period. The whole population begins as unvaccinated. On the ageing date, individuals were removed from the model at age-specific mortality rates (μ_i_), and aged into the next age groups at rates proportional to their size. Susceptible newborns were introduced at a rate proportional to the crude birth rate (CBR). Over the vaccination period (12 weeks), individuals were moved into the vaccinated compartment at age-specific rates which depend on vaccination coverage and efficacy (v_i_). After the vaccination period, individuals lost their vaccine-induced immunity and moved back into the unvaccinated compartment at rate ω, which varies by vaccine type. The ageing, waning, and vaccination occurs again annually.

**Figure S21:** Example vaccination coverage in the 0-4 age group in a Northern Hemisphere country, under 70% vaccination coverage in the 0-4 age group.

**Table S6:** Global average annual vaccine doses given over the 30-year projection period under each age-targeting strategy and vaccine type, under 50% vaccination coverage.

**Figure S22:** Annual age-specific vaccine doses given under each age-targeting strategy and vaccine type, assuming 50% vaccination coverage.

**Figure S23:** Annual vaccine doses given worldwide, stratified by vaccination status of the recipient, under 50% vaccination coverage of 0-17 and 65+ age groups.

**Figure S24:** Proportion of annual age-specific vaccine doses given to already-vaccinated individuals (‘null’), assuming 50% vaccination coverage of 0-17 and 65+ age groups.

**Figure S25:** Overlay of ten simulations of influenza incidence in each exemplar country with no vaccination coverage, stratified by strain.

**Table S7:** Annual influenza infections under each vaccine type and age-targeting strategy, assuming 50% vaccination coverage (median, 95% uncertainty intervals).

**Figure S26:** Median distribution of influenza infections across age groups under no vaccinations in each WHO region (shown as crosses), compared to distribution of the 2025 population (shown as triangles).

**Table S8:** Annual influenza infections averted under each vaccine type and age-targeting strategy, assuming 50% vaccination coverage (median, 95% uncertainty intervals), and median percentage of influenza infections averted, compared to under no vaccinations.

**Figure S27:** Number needed to vaccinate, stratified by WHO region, under each age-targeting strategy and vaccine type.

**Figure S28:** Overview of the economic decision tree model.

**Table S9:** Probabilities of symptomatic influenza and fever upon infection.

**Figure S29:** Age-specific national IFRs, per 100,000 infections. Map base layer from https://gis-who.hub.arcgis.com/pages/detailedboundary.

**Table S10:** Calculated age-specific infection hospitalisation ratios.

**Table S11:** Influenza disability weights for each health outcome [54].

**Figure S30:** Mean estimated costs of care for adult, children, and elderly hospitalisations and outpatient visits, with GDP per capita shown on a log scale. GDP per capita and costs of care in 2022 USD. Data points shown are estimates from the literature.

**Figure S31**: National willingness-to-pay thresholds [63] and 50% of 2022 GDP per capita. Dotted line indicates y=x.

**Figure S32:** Costs of vaccine dose delivery in LMICs from Portnoy et al. [64] with 95% uncertainty intervals (black), and additional HIC data for regression (red), against healthcare expenditure per capita, on a log-log scale.

**Figure S33:** Global number needed to vaccinate to prevent one influenza-associated infection, hospitalisation, or death, for each vaccine type and age-targeting strategy, on a log scale.

**Figure S34:** Number needed to vaccinate to avert one DALY in each WHO region, for each vaccine type and age-targeting strategy, on a log scale.

**Figure S35:** Averted annual age-specific health outcomes under each age-targeting strategy and vaccine type, in the African Region.

**Figure S36:** Averted annual age-specific health outcomes under each age-targeting strategy and vaccine type, in the Region of the Americas.

**Figure S37:** Averted annual age-specific health outcomes under each age-targeting strategy and vaccine type, in the Eastern Mediterranean Region.

**Figure S38:** Averted annual age-specific health outcomes under each age-targeting strategy and vaccine type, in the European Region.

**Figure S39:** Averted annual age-specific health outcomes under each age-targeting strategy and vaccine type, in the South-East Asian Region.

**Figure S40:** Averted annual age-specific health outcomes under each age-targeting strategy and vaccine type, in the Western Pacific Region.

**Figure S41:** Median national threshold vaccine prices in each WHO Region, for each vaccine type and age-targeting strategy.

**Table S12:** Regional minimum and maximum annual averted outcomes per 100,000 population between 2025-2029 (inclusive), under 50% vaccination coverage in under 18-year-olds (median and 95% uncertainty ranges), for each vaccine type. Range of years chosen for increased comparability with current population sizes.

**Figure S42:** Number of countries in which each age-targeting strategy has the highest median threshold price, under each vaccine type.

**Table S13:** Minimum and maximum national threshold prices in each World Bank income group, assuming 50% vaccination coverage, under each age-targeting strategy and vaccine type, and proportion of countries in which the median threshold cost is above $0.

**Figure S43:** Global annual averted age-specific health outcomes under each age-targeting strategy and vaccine type, under 20%, 50%, and 70% vaccination coverage.

**Table S14:** Annual global averted infections, hospitalisations, and deaths under 20%, 50%, and 70% coverage, under the 0-10 age-targeting strategy (median and 95% uncertainty ranges).

**Figure S44:** Median national threshold vaccine prices in each World Bank income group, for each vaccine type and age-targeting strategy, with reduced relative infectiousness in vaccinated individuals.

**Figure S45:** Median national threshold vaccine prices in each World Bank income group, for each vaccine type and age-targeting strategy, with disease modification in vaccinated individuals.

**Figure S46:** Number needed to vaccinate associated with original vaccine mechanisms and with reduced relative infectiousness of vaccinated individuals, under each age-targeting strategy and vaccine type, with 50% and 95% uncertainty intervals.

**Table S15:** Vaccine characteristics under the base case, breath, and depth scenarios.

**Figure S47:** Number needed to vaccinate for each original and modified vaccine type, under each age-targeting strategy and vaccine type, with 50% and 95% uncertainty intervals.

**Figure S48:** Median national threshold vaccine prices in each World Bank income group, for each vaccine type and age-targeting strategy, with willingness-to-pay thresholds set as 50% of GDP per capita.

**Figure S49:** Median national threshold vaccine prices in each World Bank income group, for each vaccine type and age-targeting strategy, with discount rates for DALYs set at 0%.

**Figure S50:** Median national threshold vaccine prices in each World Bank income group, for each vaccine type and age-targeting strategy, with the inclusion of outpatient visits and their associated costs.

**Table S16:** Productivity costs saved in 2025-2050, inclusive, by 50% vaccination coverage in individuals aged 0-17, for various influenza vaccines, in each WHO region. Costs presented in $2022, discounted at a rate of 3%.

**Figure S51:** PRISMA flow diagram of the selection of studies reporting infection-fatality ratios. **Table S17:** Characteristics of the studies included in the review.

**Figure S52:** Forest plot of seasonal influenza IFR estimates from the Hong Kong study and from the model. The empirical estimates are from three different periods during 2009 through to 2011 and from two influenza strains, A(H3N2) and A(H1N1) 2009.

**Figure S53:** Forest plot of A(H1N1) 2009 pandemic influenza IFR estimates from empirical studies and from the seasonal influenza model.

