## Supplementary Material for "The potential global impact and cost-effectiveness of next-generation influenza vaccines: a modelling analysis"

**Contents**

**1. Model parameters	4**

**2. Data preparation	5**

a. Influenza data selection	5

b. Influenza Transmission Zone classification	5

c. Exemplar country selection	14

d. Epidemic identification algorithm	15

**3. Inference methodology	15**

a. Demography	15

b. Contact matrices	15

c. The transmission model	16

d. Vaccination coverage	18

e. Matching of years	21

f. The MCMC and inference model	21

**4. Inference outputs	22**

**5. Simulating epidemics	31**

a. Demographic changes	31

b. Contact matrices	32

c. Epidemic timing	32

d. Epidemic simulation	32

**6. Simulation outputs	34**

a. Population-level immunity	34

b. Vaccine doses	34

c. Influenza incidence	37

d. Infections averted	40

e. Number needed to vaccinate (NNV)	41

f. Scaling of contact matrices	42

**7. Economic modelling	43**

a. Health outcomes data	43

Symptomatic and fever probabilities	43

Infection-fatality ratios	44

Infection-hospitalisation ratios	45

Outpatient visit rates	46

Disability-adjusted life years	46

b. Economic inputs	46

Cost of hospitalisation/outpatient	46

Willingness-to-pay	47

Vaccine delivery cost	48

c. Health outcomes	49

**8. Threshold prices	56**

**9. Sensitivity analyses	62**

a. Coverage levels	62

b. Vaccine mechanisms	64

c. Breadth and depth	65

d. Willingness-to-pay at 50% of GDP per capita	67

e. DALY discount rate at 0%	68

f. Outpatient inclusion	69

g. Productivity costs	70

**10. Systematised review of estimates of seasonal influenza infection-fatality risk	71**

a. Objective	71

b. Methods	72

Eligibility criteria	72

Information sources	72

Search strategy	72

Selection process	73

Data collection and analysis	73

c. Results	73

Studies selected	73

Study characteristics	73

Results of individuals studies	73

Comparison with the model	73

d. Conclusion	74

Range of the studies found	74

Comparison with the model	74

**References	79**

### 1. Model parameters

| **Parameter** | **Definition** | **Model assumption** | **Value (if fixed)** |
| --- | --- | --- | --- |
| 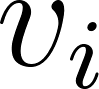<https://www.codecogs.com/eqnedit.php?latex=v_i#0> | Age-specific vaccination rate | Dependent on coverage and mean vaccine-induced immunity duration | - |
| 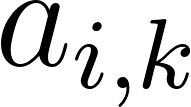<https://www.codecogs.com/eqnedit.php?latex=a_%7Bi%2Ck%7D#0> | Vaccine efficacy | Vaccine assumption | See Table 1 |
| 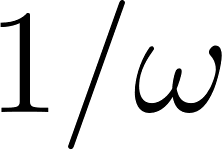<https://www.codecogs.com/eqnedit.php?latex=1%2F%5Comega#0> | Mean vaccine-induced immunity duration | Vaccine assumption | See Table 1 |
| 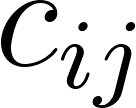<https://www.codecogs.com/eqnedit.php?latex=c_%7Bij%7D#0> | Contact rates between age groups i and j | Based on [1], reweighted for annual age structure | - |
| 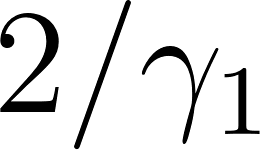<https://www.codecogs.com/eqnedit.php?latex=2%2F%5Cgamma_%7B1%7D#0> | Latency period | Fixed, based on literature [2] | 0.8 days |
| 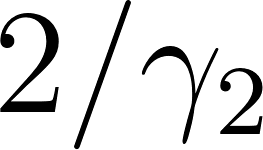<https://www.codecogs.com/eqnedit.php?latex=2%2F%5Cgamma_%7B2%7D#0> | Infectious period | Fixed, based on literature [2] | 1.8 days |
| 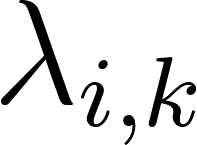<https://www.codecogs.com/eqnedit.php?latex=%5Clambda_%7Bi%2Ck%7D#0> | Age- and epidemic-specific force of infection | Depends on epidemic-specific susceptibility and transmissibility, and age-specific contact rates | - |
| 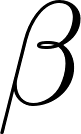<https://www.codecogs.com/eqnedit.php?latex=%5Cbeta#0> | Transmissibility | Posterior estimated from Flunet data | - |
| 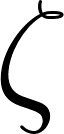<https://www.codecogs.com/eqnedit.php?latex=%5Czeta#0> | Susceptibility | Posterior estimated from Flunet data | - |

**Table S1:** Model parameters, used in the epidemic inference, vaccination, and epidemic models (steps 1-3).

### 2. Data preparation

#### a. Influenza data selection

We used global influenza surveillance data from FluNet (https://www.who.int/tools/flunet) to measure the reported incidence of influenza in an inference period of 1st January 2010 to 31st December 2019. This time period was bracketed by the A/H1N1pdm and COVID-19 pandemic, both of which disrupted influenza epidemiology. For each country, we extracted the weekly positive number of laboratory-confirmed influenza A viruses and influenza B viruses, and total number of samples processed. We used data collected from non-sentinel sources (e.g. universal testing, testing at point of care).

#### b. Influenza Transmission Zone classification

Because only a few countries had a long time series of influenza data from FluNet, we projected characteristics of influenza transmission inferred for a limited number of countries onto the rest of the world. To do this, we used a global categorisation of countries by relatively similar influenza transmission patterns. The WHO first classified 18 influenza transmission zones (ITZs) in 2009 based on influenza transmission patterns; several papers have since attempted to reclassify and recategorise the ITZs using various epidemic parameters and classification methods [3,4].

We use the classifications of ITZs produced by Chen et al. [5], as this study used a largely similar dataset to our inference (the epidemiological parameters were calculated using FluNet data from July 2010 to June 2019) and validated their results. Chen et al. used a k-means clustering approach to classify 109 countries into seven ITZs which reflect common influenza characteristics. The parameters used for the clustering were: start week, peak week, primary week (determined by detrending the time series), and positivity rate for total influenza virus, influenza H1, H3, and B viruses, and location information (latitude and longitude of the capital city). The countries used for the clustering analysis were chosen based on the availability of FluNet data in the chosen time period, and the consistency of the ITZs was verified by hierarchical clustering methods. The resulting ITZs are: Northern America, Eastern & Southern-Asia, Europe, Asia-Europe, Southern-America, Oceania-Melanesia-Polynesia, and Africa. 72 countries were included in FluNet’s database but not included in the clustering produced in [5] due to insufficient surveillance data in FluNet.

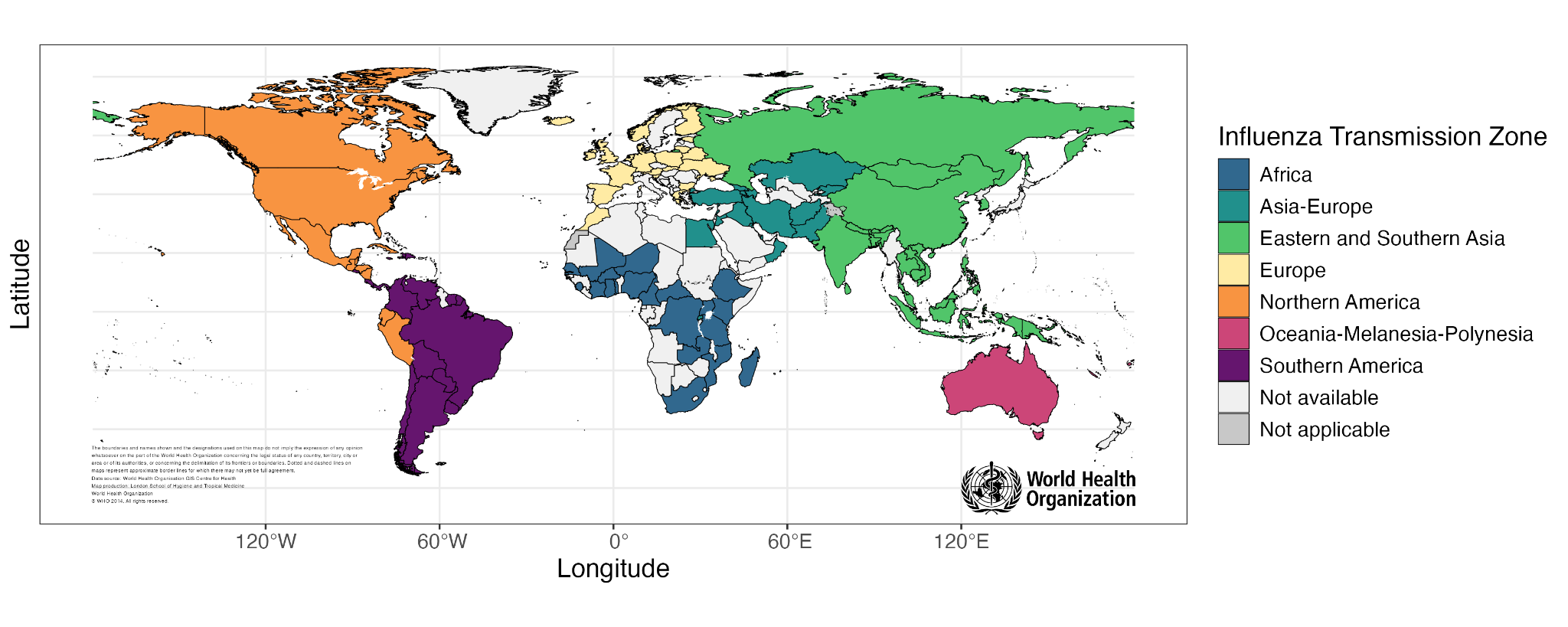

**Figure S1:** Geographical distribution of the seven ITZs produced by Chen et al. [5]. Map base layer from <https://gis-who.hub.arcgis.com/pages/detailedboundary>.

We expanded the list of countries included in this project by also considering all countries for which contact matrices were produced in Prem et al. [1], and any countries in the World Population Prospects which are projected to have at least one million residents by 2025. 186 countries were in the final set (109 in Chen et al., 177 in Prem et al., 4 in neither).

We assigned each country to an existing ITZ based solely on their geographical parameters (latitude and longitude of the capital city), using the assignment step of k-means clustering without updating any pre-assigned countries. We first calculated the geographical cluster centroids of each existing ITZ *C_i* using the capital cities *c* of the countries assigned by [5]:

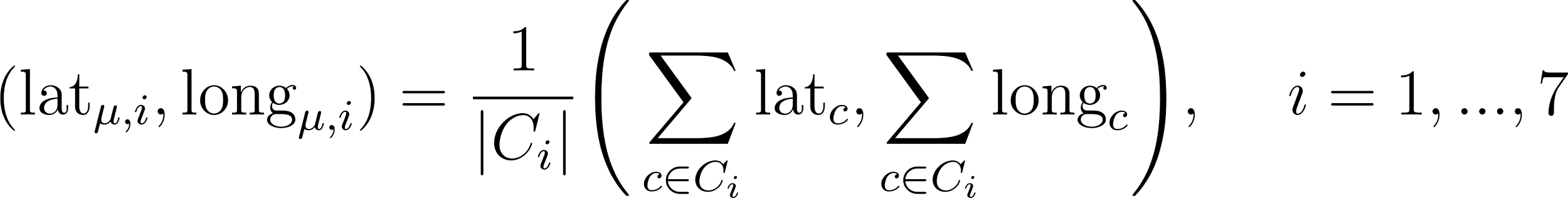

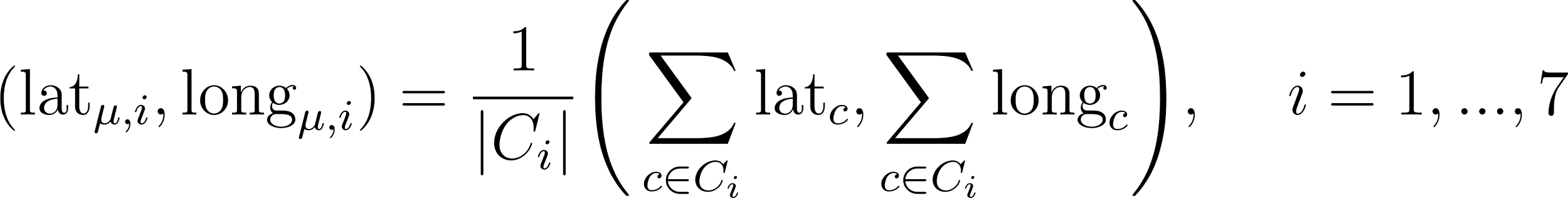
<https://www.codecogs.com/eqnedit.php?latex=(%5Ctext%7Blat%7D_%7B%5Cmu%2C%20i%7D%2C%20%5Ctext%7Blong%7D_%7B%5Cmu%2C%20i%7D)%20%3D%20%5Cfrac%7B1%7D%7B%7CC_i%7C%7D%5CBigg(%5Csum_%7Bc%20%5Cin%20C_i%7D%20%5Ctext%7Blat%7D_%7Bc%7D%2C%20%5Csum_%7Bc%20%5Cin%20C_i%7D%20%5Ctext%7Blong%7D_%7Bc%7D%5CBigg)%2C%20%5Chspace%7B5mm%7D%20i%20%3D%201%2C...%2C7#0>

We then assigned a country with a capital city of latitude *x* and longitude *y* to the cluster which minimised the Euclidean distance from the cluster centroid to the capital city:

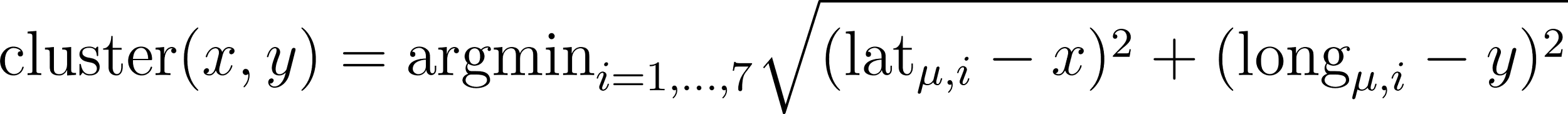

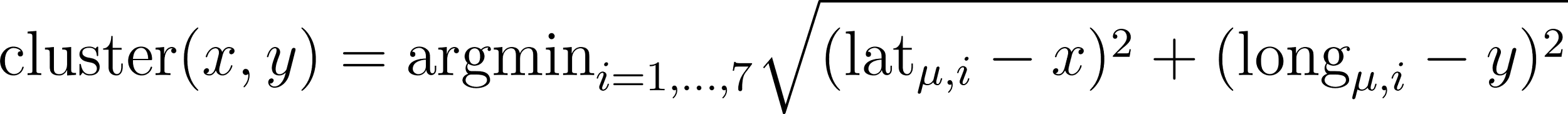
<https://www.codecogs.com/eqnedit.php?latex=%5Ctext%7Bcluster%7D(x%2Cy)%20%3D%20%5Ctext%7Bargmin%7D_%7Bi%3D1%2C...%2C7%7D%20%5Csqrt%7B(%5Ctext%7Blat%7D_%7B%5Cmu%2C%20i%7D%20-%20x)%5E2%20%2B%20(%5Ctext%7Blong%7D_%7B%5Cmu%2C%20i%7D%20-%20y)%5E2%7D#0>

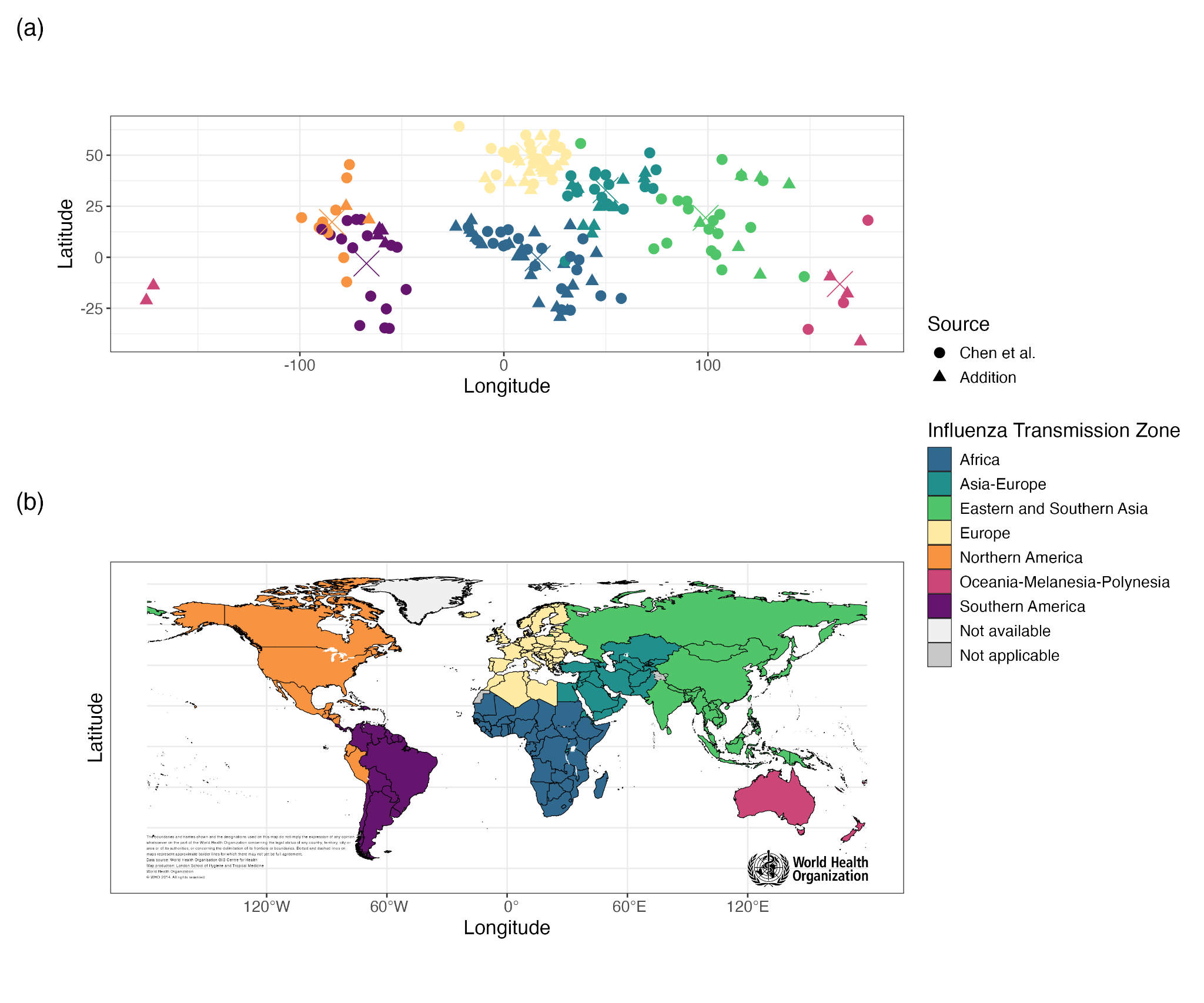

**Figure S2: (a)** The longitude and latitude of the capital cities of each country in the ITZs, and each ITZ’s cluster centroid (marked as X). **(b)** World map of all countries included in this analysis. Map base layer from <https://gis-who.hub.arcgis.com/pages/detailedboundary>.

In order to validate the use of geographical parameters alone to assign the remaining 77 countries to pre-existing ITZs, we also investigated which ITZs the countries assigned by [5] would be assigned to under this new metric. There is a clear geographical association between the countries in each pre-existing ITZ (Figure S1), which is confirmed by this investigation - only 11 of the 109 countries would be assigned to a new cluster (Costa Rica, Dominican Republic, Ecuador, El Salvador, Haiti, Jamaica, Panama, Papua New Guinea, Peru, Russian Federation, Rwanda), 8 of which involve re-classifications between Northern America and Southern America.

| **ISO3C Code** | **Name** | **ITZ** | **Source** |
| --- | --- | --- | --- |
| AFG | Afghanistan | Asia-Europe | Chen et al. |
| AGO | Angola | Africa | Addition |
| ALB | Albania | Europe | Addition |
| ARE | United Arab Emirates | Asia-Europe | Addition |
| ARG | Argentina | Southern America | Chen et al. |
| ARM | Armenia | Asia-Europe | Chen et al. |
| AUS | Australia | Oceania-Melanesia-Polynesia | Chen et al. |
| AUT | Austria | Europe | Chen et al. |
| AZE | Azerbaijan | Asia-Europe | Chen et al. |
| BDI | Burundi | Africa | Addition |
| BEL | Belgium | Europe | Addition |
| BEN | Benin | Africa | Addition |
| BFA | Burkina Faso | Africa | Chen et al. |
| BGD | Bangladesh | Eastern and Southern Asia | Chen et al. |
| BGR | Bulgaria | Europe | Chen et al. |
| BHR | Bahrain | Asia-Europe | Chen et al. |
| BHS | Bahamas | Northern America | Addition |
| BIH | Bosnia & Herzegovina | Europe | Addition |
| BLR | Belarus | Europe | Chen et al. |
| BLZ | Belize | Northern America | Chen et al. |
| BOL | Bolivia | Southern America | Chen et al. |
| BRA | Brazil | Southern America | Chen et al. |
| BRB | Barbados | Southern America | Addition |
| BRN | Brunei | Eastern and Southern Asia | Addition |
| BTN | Bhutan | Eastern and Southern Asia | Chen et al. |
| BWA | Botswana | Africa | Addition |
| CAF | Central African Republic | Africa | Chen et al. |
| CAN | Canada | Northern America | Chen et al. |
| CHE | Switzerland | Europe | Addition |
| CHL | Chile | Southern America | Chen et al. |
| CHN | China | Eastern and Southern Asia | Chen et al. |
| CIV | Côte d'Ivoire | Africa | Chen et al. |
| CMR | Cameroon | Africa | Chen et al. |
| COD | Congo - Kinshasa | Africa | Chen et al. |
| COG | Congo - Brazzaville | Africa | Addition |
| COL | Colombia | Southern America | Chen et al. |
| COM | Comoros | Africa | Addition |
| CPV | Cape Verde | Africa | Addition |
| CRI | Costa Rica | Southern America | Chen et al. |
| CUB | Cuba | Northern America | Chen et al. |
| CYP | Cyprus | Asia-Europe | Addition |
| CZE | Czechia | Europe | Addition |
| DEU | Germany | Europe | Chen et al. |
| DJI | Djibouti | Asia-Europe | Addition |
| DNK | Denmark | Europe | Chen et al. |
| DOM | Dominican Republic | Southern America | Chen et al. |
| DZA | Algeria | Europe | Addition |
| ECU | Ecuador | Northern America | Chen et al. |
| EGY | Egypt | Asia-Europe | Chen et al. |
| ERI | Eritrea | Asia-Europe | Addition |
| ESP | Spain | Europe | Chen et al. |
| EST | Estonia | Europe | Chen et al. |
| ETH | Ethiopia | Africa | Chen et al. |
| FIN | Finland | Europe | Chen et al. |
| FJI | Fiji | Oceania-Melanesia-Polynesia | Chen et al. |
| FRA | France | Europe | Chen et al. |
| GAB | Gabon | Africa | Addition |
| GBR | United Kingdom | Europe | Chen et al. |
| GEO | Georgia | Asia-Europe | Chen et al. |
| GHA | Ghana | Africa | Chen et al. |
| GIN | Guinea | Africa | Addition |
| GMB | Gambia | Africa | Addition |
| GNB | Guinea-Bissau | Africa | Addition |
| GNQ | Equatorial Guinea | Africa | Addition |
| GRC | Greece | Europe | Chen et al. |
| GTM | Guatemala | Northern America | Chen et al. |
| GUF | French Guiana | Southern America | Chen et al. |
| GUY | Guyana | Southern America | Addition |
| HKG | Hong Kong SAR China | Eastern and Southern Asia | Addition |
| HND | Honduras | Northern America | Chen et al. |
| HRV | Croatia | Europe | Addition |
| HTI | Haiti | Southern America | Chen et al. |
| HUN | Hungary | Europe | Addition |
| IDN | Indonesia | Eastern and Southern Asia | Chen et al. |
| IND | India | Eastern and Southern Asia | Chen et al. |
| IRL | Ireland | Europe | Chen et al. |
| IRN | Iran | Asia-Europe | Chen et al. |
| IRQ | Iraq | Asia-Europe | Chen et al. |
| ISL | Iceland | Europe | Chen et al. |
| ISR | Israel | Asia-Europe | Addition |
| ITA | Italy | Europe | Addition |
| JAM | Jamaica | Southern America | Chen et al. |
| JOR | Jordan | Asia-Europe | Chen et al. |
| KAZ | Kazakhstan | Asia-Europe | Chen et al. |
| KEN | Kenya | Africa | Chen et al. |
| KGZ | Kyrgyzstan | Asia-Europe | Chen et al. |
| KHM | Cambodia | Eastern and Southern Asia | Chen et al. |
| KOR | South Korea | Eastern and Southern Asia | Chen et al. |
| KWT | Kuwait | Asia-Europe | Chen et al. |
| LAO | Laos | Eastern and Southern Asia | Chen et al. |
| LBN | Lebanon | Asia-Europe | Chen et al. |
| LBR | Liberia | Africa | Addition |
| LBY | Libya | Europe | Addition |
| LCA | St. Lucia | Southern America | Addition |
| LKA | Sri Lanka | Eastern and Southern Asia | Chen et al. |
| LSO | Lesotho | Africa | Addition |
| LTU | Lithuania | Europe | Chen et al. |
| LUX | Luxembourg | Europe | Addition |
| LVA | Latvia | Europe | Addition |
| MAC | Macao SAR China | Eastern and Southern Asia | Addition |
| MAR | Morocco | Europe | Chen et al. |
| MDA | Moldova | Europe | Addition |
| MDG | Madagascar | Africa | Chen et al. |
| MDV | Maldives | Eastern and Southern Asia | Chen et al. |
| MEX | Mexico | Northern America | Chen et al. |
| MKD | North Macedonia | Europe | Addition |
| MLI | Mali | Africa | Chen et al. |
| MLT | Malta | Europe | Chen et al. |
| MMR | Myanmar (Burma) | Eastern and Southern Asia | Addition |
| MNE | Montenegro | Europe | Addition |
| MNG | Mongolia | Eastern and Southern Asia | Chen et al. |
| MOZ | Mozambique | Africa | Chen et al. |
| MRT | Mauritania | Africa | Addition |
| MUS | Mauritius | Africa | Chen et al. |
| MWI | Malawi | Africa | Addition |
| MYS | Malaysia | Eastern and Southern Asia | Chen et al. |
| NAM | Namibia | Africa | Addition |
| NCL | New Caledonia | Oceania-Melanesia-Polynesia | Chen et al. |
| NER | Niger | Africa | Chen et al. |
| NGA | Nigeria | Africa | Chen et al. |
| NIC | Nicaragua | Northern America | Chen et al. |
| NLD | Netherlands | Europe | Chen et al. |
| NOR | Norway | Europe | Chen et al. |
| NPL | Nepal | Eastern and Southern Asia | Chen et al. |
| NZL | New Zealand | Oceania-Melanesia-Polynesia | Addition |
| OMN | Oman | Asia-Europe | Chen et al. |
| PAK | Pakistan | Asia-Europe | Chen et al. |
| PAN | Panama | Southern America | Chen et al. |
| PER | Peru | Northern America | Chen et al. |
| PHL | Philippines | Eastern and Southern Asia | Chen et al. |
| PNG | Papua New Guinea | Eastern and Southern Asia | Chen et al. |
| POL | Poland | Europe | Chen et al. |
| PRI | Puerto Rico | Northern America | Addition |
| PRK | North Korea | Eastern and Southern Asia | Addition |
| PRT | Portugal | Europe | Addition |
| PRY | Paraguay | Southern America | Chen et al. |
| PSE | Palestine | Asia-Europe | Addition |
| QAT | Qatar | Asia-Europe | Chen et al. |
| ROU | Romania | Europe | Addition |
| RUS | Russia | Eastern and Southern Asia | Chen et al. |
| RWA | Rwanda | Asia-Europe | Chen et al. |
| SAU | Saudi Arabia | Asia-Europe | Addition |
| SDN | Sudan | Africa | Addition |
| SEN | Senegal | Africa | Chen et al. |
| SGP | Singapore | Eastern and Southern Asia | Chen et al. |
| SLB | Solomon Islands | Oceania-Melanesia-Polynesia | Addition |
| SLE | Sierra Leone | Africa | Chen et al. |
| SLV | El Salvador | Southern America | Chen et al. |
| SRB | Serbia | Europe | Addition |
| SSD | South Sudan | Africa | Addition |
| STP | São Tomé and Príncipe | Africa | Addition |
| SUR | Suriname | Southern America | Chen et al. |
| SVK | Slovakia | Europe | Chen et al. |
| SVN | Slovenia | Europe | Chen et al. |
| SWE | Sweden | Europe | Addition |
| SWZ | Eswatini | Africa | Addition |
| SYR | Syria | Asia-Europe | Addition |
| TCD | Chad | Africa | Addition |
| TGO | Togo | Africa | Chen et al. |
| THA | Thailand | Eastern and Southern Asia | Chen et al. |
| TJK | Tajikistan | Asia-Europe | Addition |
| TKM | Turkmenistan | Asia-Europe | Addition |
| TLS | Timor-Leste | Eastern and Southern Asia | Addition |
| TON | Tonga | Oceania-Melanesia-Polynesia | Addition |
| TTO | Trinidad & Tobago | Southern America | Addition |
| TUN | Tunisia | Europe | Addition |
| TUR | Turkey | Asia-Europe | Chen et al. |
| TZA | Tanzania | Africa | Chen et al. |
| UGA | Uganda | Africa | Chen et al. |
| UKR | Ukraine | Europe | Chen et al. |
| URY | Uruguay | Southern America | Chen et al. |
| USA | United States | Northern America | Chen et al. |
| UZB | Uzbekistan | Asia-Europe | Addition |
| VCT | St. Vincent & Grenadines | Southern America | Addition |
| VEN | Venezuela | Southern America | Chen et al. |
| VNM | Vietnam | Eastern and Southern Asia | Chen et al. |
| VUT | Vanuatu | Oceania-Melanesia-Polynesia | Addition |
| WSM | Samoa | Oceania-Melanesia-Polynesia | Addition |
| YEM | Yemen | Asia-Europe | Addition |
| ZAF | South Africa | Africa | Chen et al. |
| ZMB | Zambia | Africa | Chen et al. |
| ZWE | Zimbabwe | Africa | Addition |
| JPN | Japan | Eastern and Southern Asia | Addition |
| TWN | Taiwan | Eastern and Southern Asia | Addition |
| SOM | Somalia | Africa | Addition |
| XKX | Kosovo | Europe | Addition |

**Table S2:** Influenza Transmission Zone assignments of 186 countries, assigned either by Chen et al. [5] or added based on geographical parameters.

#### c. Exemplar country selection

Exemplar countries for each ITZ chosen on the basis of data availability (number of years of data in the coverage, average annual number of tests processed by strain and data source type) in FluNet over the inference period of January 2010 – December 2019:

Argentina - Southern-America

Australia - Oceania-Melanesia-Polynesia

Canada - Northern America

China - Eastern & Southern-Asia

Ghana – Africa

Turkey - Asia-Europe

United Kingdom (totalled over England, Scotland, Wales, Northern Ireland) – Europe

| **Country** | **Number of weeks of data** | **Mean number of specimens processed per week** | **Mean number of confirmed Influenza A infections per week** | **Mean number of confirmed Influenza B infections per week** |
| --- | --- | --- | --- | --- |
| **China** | 517 | 6620 | 639 | 355 |
| **Turkey** | 441 | 150 | 30.7 | 9.73 |
| **Argentina** | 521 | 1430 | 68.9 | 14.7 |
| **Ghana** | 520 | 75.4 | 8.65 | 3.18 |
| **Canada** | 519 | 4220 | 489 | 154 |
| **Australia** | 520 | 706 | 79.5 | 21.7 |
| **United Kingdom** | 521 | 960 | 225 | 75.4 |

**Table S3:** Summary of FluNet data for each of the chosen exemplar countries between January 2010 and December 2019.

#### d. Epidemic identification algorithm

In each of the chosen exemplar countries, we identified distinct epidemics in the inference period using weekly confirmed cases counts for both influenza A and influenza B viruses from non-sentinel data sources in the FluNet data.

A time period is defined as a distinct epidemic if:

(i) it is 8 weeks or longer;

(ii) it includes at least one week with reported positivity greater than the 65th percentile (in Australia, Canada, China) or 80th percentile (in Argentina, Ghana, Turkey, United Kingdom) of the inference period’s positivity (within that country and strain);

(iii) all weeks have a reported positivity greater than the 50th percentile of the inference period’s positivity (within that country and strain).

The necessary peak percentile threshold varied by countries on account of increased noise in the data in some time series. An added condition was in place when analysing the data in Australia and China, to ensure the uncoupling of separate epidemics occurring close together:

(iv) At no point after the first 8 weeks falls below the 64th (Australia) or 60th (China) percentile of the decade’s positivity, in which case the epidemic ends.

### 3. Inference methodology

#### a. Demography

The demography of each exemplar country was extracted from the World Population Prospects population data using the *socialmixr* R package [6]. We used data from 2015, as the dataset only provides population estimates every 5 years and this falls in the middle of our inference period. We then aggregated the population of each country into the model age groups of [0,5), [5,20), [20,65), [65, 120], producing the demography n=(n_1_, n_2_, n_3_, n_4_).

#### b. Contact matrices

To quantify contact patterns for inference in each of the exemplar countries, we used contact matrices derived in Prem et al. [1], which constructed synthetic contact matrices over 16 age groups in 177 countries using setting-specific survey data including household, school, classroom, and workplace composition. Australia, which is not included in Prem et al.’s output, was conducted using the underlying contact patterns of New Zealand.

We aggregated the daily age-specific contacts into the model age groups to produce a contact matrix:

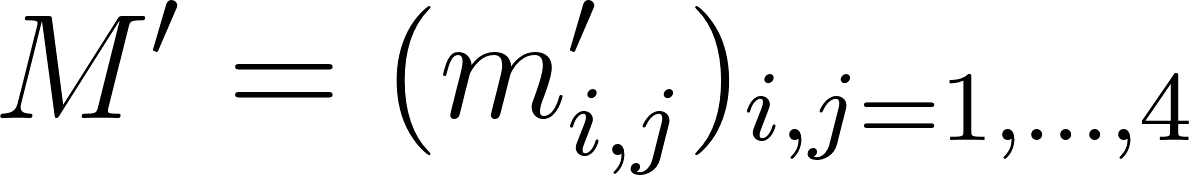
<https://www.codecogs.com/eqnedit.php?latex=M'%20%3D%20(m'_%7Bi%2Cj%7D)_%7Bi%2Cj%20%3D%201%2C...%2C4%7D%20#0>

We then ensures reciprocity with respect to the 2015 population (*n*) by constructing a reciprocal contact matrix:

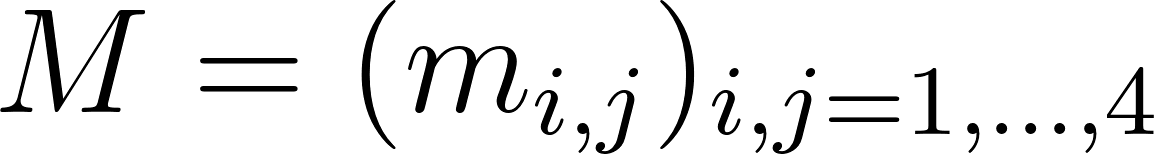
<https://www.codecogs.com/eqnedit.php?latex=M%20%3D%20(m_%7Bi%2Cj%7D)_%7Bi%2Cj%20%3D%201%2C...%2C4%7D%20#0>

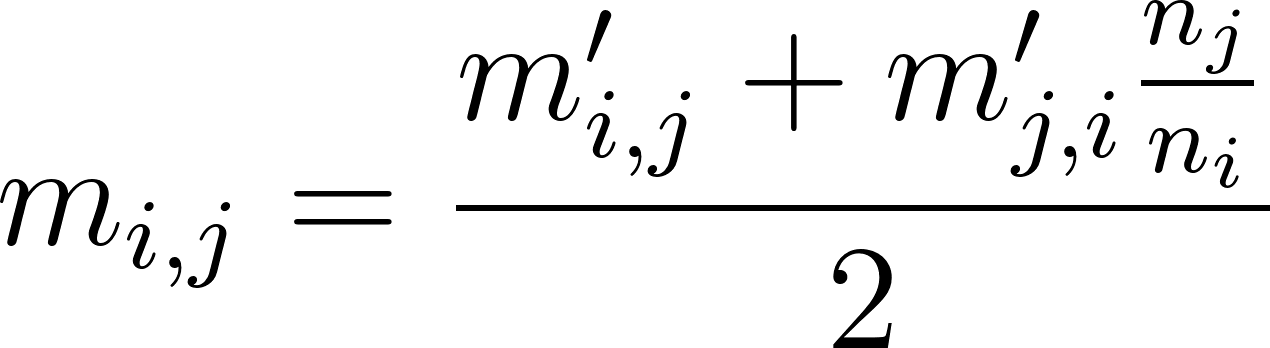
<https://www.codecogs.com/eqnedit.php?latex=m_%7Bi%2Cj%7D%20%3D%20%5Cfrac%7Bm'_%7Bi%2Cj%7D%20%2B%20m'_%7Bj%2Ci%7D%5Cfrac%7Bn_j%7D%7Bn_i%7D%7D%7B2%7D%20#0>

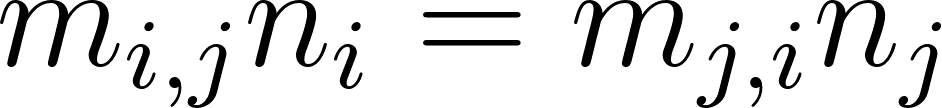
<https://www.codecogs.com/eqnedit.php?latex=m_%7Bi%2Cj%7D%20n_i%20%3D%20m_%7Bj%2Ci%7D%20n_j%20#0>

We constructed a matrix of per-capita probabilities of contact with a specific individual in an age group, by dividing through by the 2015 population:

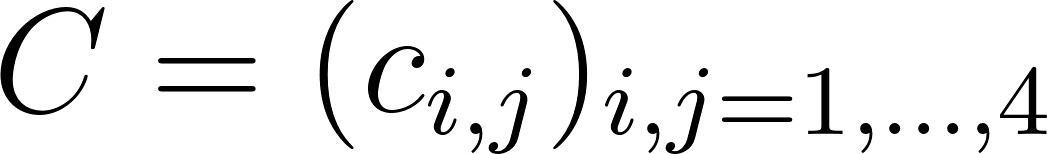
<https://www.codecogs.com/eqnedit.php?latex=C%20%3D%20(c_%7Bi%2Cj%7D)_%7Bi%2Cj%20%3D%201%2C...%2C4%7D%20#0>

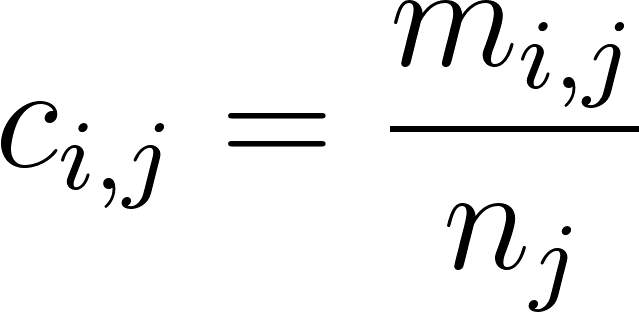
<https://www.codecogs.com/eqnedit.php?latex=c_%7Bi%2Cj%7D%20%3D%20%5Cfrac%7Bm_%7Bi%2Cj%7D%7D%7Bn_j%7D%20#0>

The matrix C was used in the transmission model.

#### c. The transmission model

The epidemic inference used a transmission model with no general vaccination model. Vaccination coverage was applied across the whole time period for simplicity (see Section 3d).

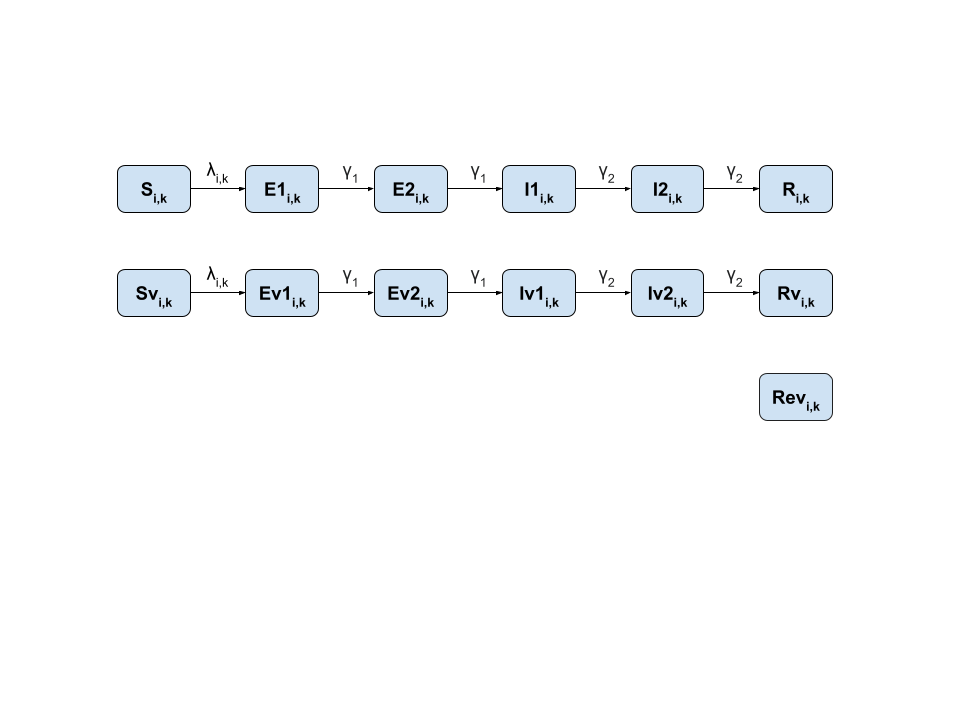

**Figure S3:** Epidemic model for inference, with no underlying vaccination model. Vaccinated individuals were assigned Rev with probability equal to vaccine efficiency.

Equations:

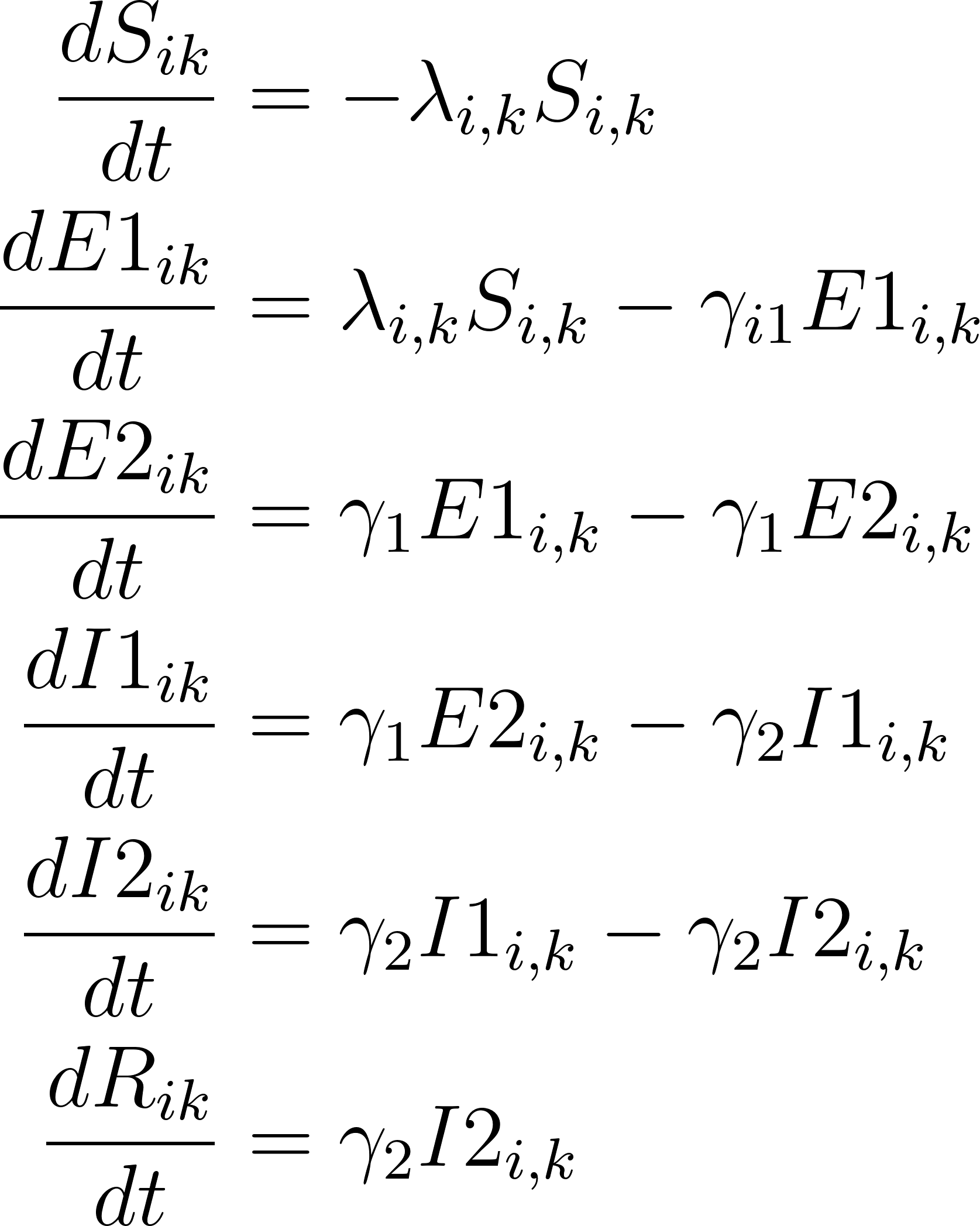
<https://www.codecogs.com/eqnedit.php?latex=%5Cbegin%7Baligned%7D%20%5Cfrac%7BdS_%7Bik%7D%7D%7Bdt%7D%20%26%3D%20-%20%5Clambda_%7Bi%2Ck%7D%20S_%7Bi%2Ck%7D%20%5C%5C%5C%5C%20%5Cfrac%7BdE1_%7Bik%7D%7D%7Bdt%7D%20%26%3D%20%5Clambda_%7Bi%2Ck%7D%20S_%7Bi%2Ck%7D%20-%20%5Cgamma_%7Bi1%7D%20E1_%7Bi%2Ck%7D%20%20%5C%5C%5C%5C%20%5Cfrac%7BdE2_%7Bik%7D%7D%7Bdt%7D%20%26%3D%20%5Cgamma_%7B1%7D%20E1_%7Bi%2Ck%7D%20-%20%5Cgamma_%7B1%7D%20E2_%7Bi%2Ck%7D%20%5C%5C%5C%5C%20%5Cfrac%7BdI1_%7Bik%7D%7D%7Bdt%7D%20%26%3D%20%5Cgamma_%7B1%7D%20E2_%7Bi%2Ck%7D%20-%20%5Cgamma_%7B2%7D%20I1_%7Bi%2Ck%7D%20%20%20%5C%5C%5C%5C%20%5Cfrac%7BdI2_%7Bik%7D%7D%7Bdt%7D%20%26%3D%20%5Cgamma_%7B2%7D%20I1_%7Bi%2Ck%7D%20-%20%5Cgamma_%7B2%7D%20I2_%7Bi%2Ck%7D%20%20%5C%5C%5C%5C%20%5Cfrac%7BdR_%7Bik%7D%7D%7Bdt%7D%20%26%3D%20%5Cgamma_%7B2%7D%20I2_%7Bi%2Ck%7D%20%5Cend%7Baligned%7D#0>

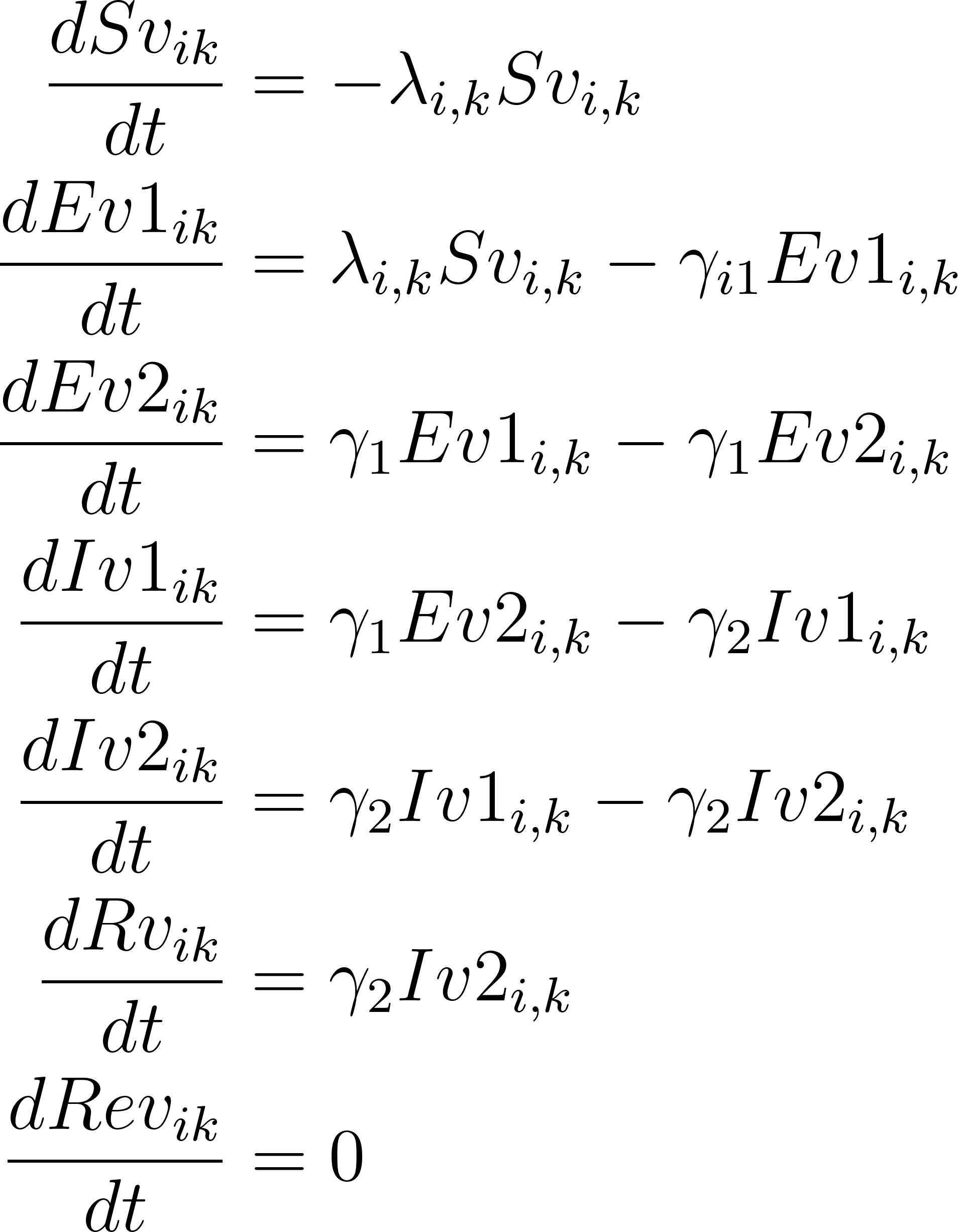
<https://www.codecogs.com/eqnedit.php?latex=%5Cbegin%7Baligned%7D%20%5Cfrac%7BdSv_%7Bik%7D%7D%7Bdt%7D%20%26%3D%20-%20%5Clambda_%7Bi%2Ck%7D%20Sv_%7Bi%2Ck%7D%20%5C%5C%5C%5C%20%5Cfrac%7BdEv1_%7Bik%7D%7D%7Bdt%7D%20%26%3D%20%5Clambda_%7Bi%2Ck%7D%20Sv_%7Bi%2Ck%7D%20-%20%5Cgamma_%7Bi1%7D%20Ev1_%7Bi%2Ck%7D%20%20%5C%5C%5C%5C%20%5Cfrac%7BdEv2_%7Bik%7D%7D%7Bdt%7D%20%26%3D%20%5Cgamma_%7B1%7D%20Ev1_%7Bi%2Ck%7D%20-%20%5Cgamma_%7B1%7D%20Ev2_%7Bi%2Ck%7D%20%5C%5C%5C%5C%20%5Cfrac%7BdIv1_%7Bik%7D%7D%7Bdt%7D%20%26%3D%20%5Cgamma_%7B1%7D%20Ev2_%7Bi%2Ck%7D%20-%20%5Cgamma_%7B2%7D%20Iv1_%7Bi%2Ck%7D%20%20%20%5C%5C%5C%5C%20%5Cfrac%7BdIv2_%7Bik%7D%7D%7Bdt%7D%20%26%3D%20%5Cgamma_%7B2%7D%20Iv1_%7Bi%2Ck%7D%20-%20%5Cgamma_%7B2%7D%20Iv2_%7Bi%2Ck%7D%20%20%5C%5C%5C%5C%20%5Cfrac%7BdRv_%7Bik%7D%7D%7Bdt%7D%20%26%3D%20%5Cgamma_%7B2%7D%20Iv2_%7Bi%2Ck%7D%20%20%5C%5C%5C%5C%20%5Cfrac%7BdRev_%7Bik%7D%7D%7Bdt%7D%20%26%3D%200%20%5Cend%7Baligned%7D#0>

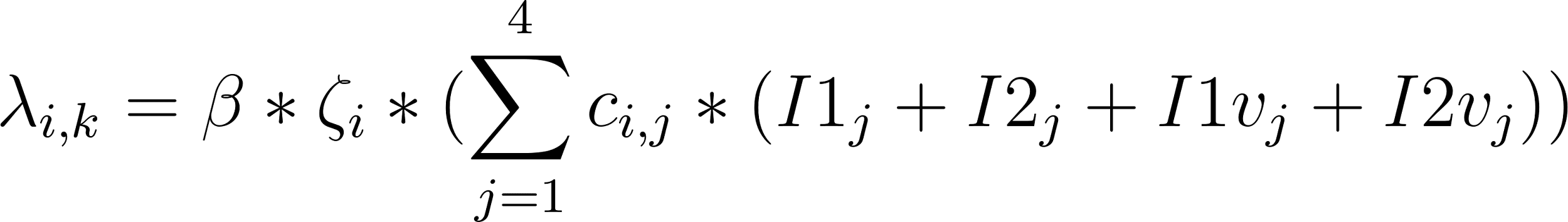
<https://www.codecogs.com/eqnedit.php?latex=%20%5Clambda_%7Bi%2Ck%7D%20%3D%20%5Cbeta%20*%20%5Czeta_i%20*%20(%5Csum_%7Bj%3D1%7D%5E%7B4%7D%20c_%7Bi%2Cj%7D%20*%20(I1_j%20%2B%20I2_j%20%2B%20I1v_j%20%2B%20I2v_j))%20#0>

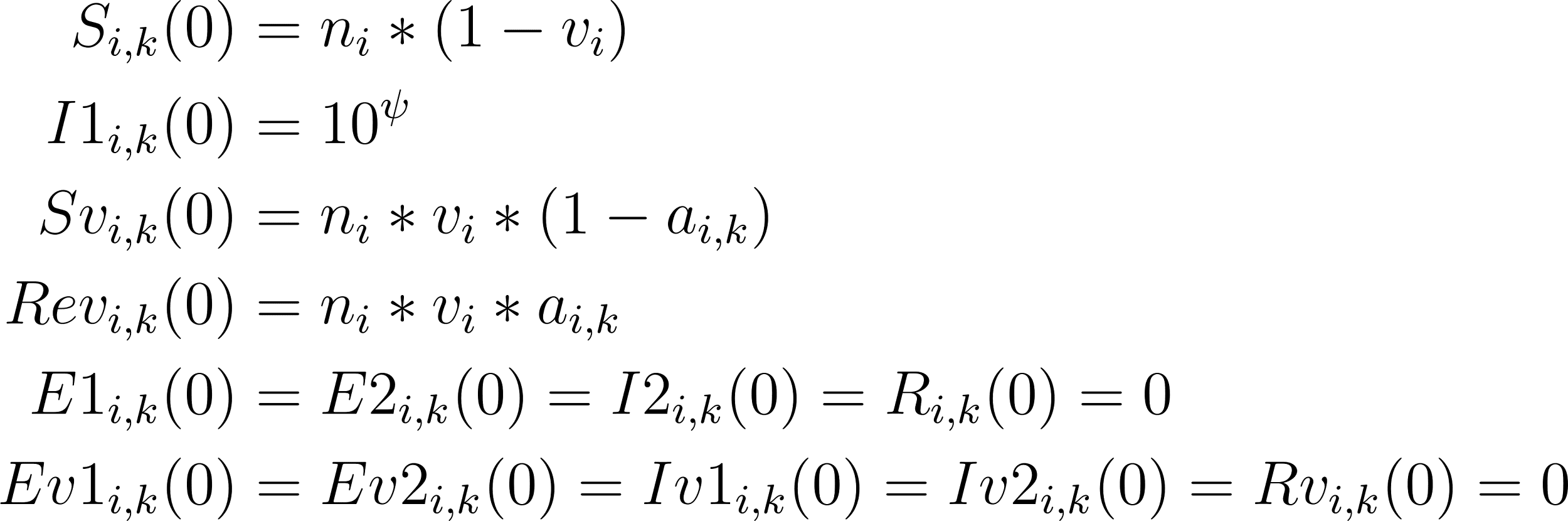
<https://www.codecogs.com/eqnedit.php?latex=%20%5Cbegin%7Baligned%7D%20%20S_%7Bi%2Ck%7D(0)%20%26%3D%20n_i%20*%20(1%20-%20v_i)%20%5C%5CI1_%7Bi%2Ck%7D(0)%20%26%3D%2010%5E%5Cpsi%20%5C%5CSv_%7Bi%2Ck%7D(0)%20%26%3D%20n_i%20*%20v_i%20*%20(1-a_%7Bi%2Ck%7D)%20%5C%5CRev_%7Bi%2Ck%7D(0)%20%26%3D%20n_i%20*%20v_i%20*%20a_%7Bi%2Ck%7D%20%5C%5CE1_%7Bi%2Ck%7D(0)%20%26%3D%20E2_%7Bi%2Ck%7D(0)%20%3D%20I2_%7Bi%2Ck%7D(0)%20%3D%20R_%7Bi%2Ck%7D(0)%20%3D%200%20%5C%5CEv1_%7Bi%2Ck%7D(0)%20%26%3D%20Ev2_%7Bi%2Ck%7D(0)%20%3D%20Iv1_%7Bi%2Ck%7D(0)%20%3D%20Iv2_%7Bi%2Ck%7D(0)%20%3D%20Rv_%7Bi%2Ck%7D(0)%20%3D%200%20%5Cend%7Baligned%7D%20#0>

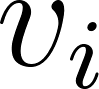
<https://www.codecogs.com/eqnedit.php?latex=v_i#0> was vaccination coverage in age group i,
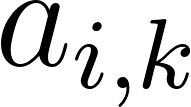
<https://www.codecogs.com/eqnedit.php?latex=a_%7Bi%2Ck%7D#0> was the vaccine efficacy, which varies by age and strain (which depended annually on whether the vaccine matches circulating strains in each hemisphere),
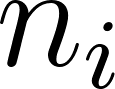
<https://www.codecogs.com/eqnedit.php?latex=n_i#0> is the age-specific population.
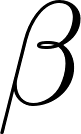
<https://www.codecogs.com/eqnedit.php?latex=%5Cbeta#0> was the transmissibility parameter, and
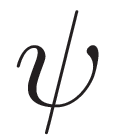
<http://www.sciweavers.org/tex2img.php?bc=Transparent&fc=Black&im=jpg&fs=100&ff=modern&edit=0&eq=%5Cpsi#0> the ‘initial infected’ parameter.
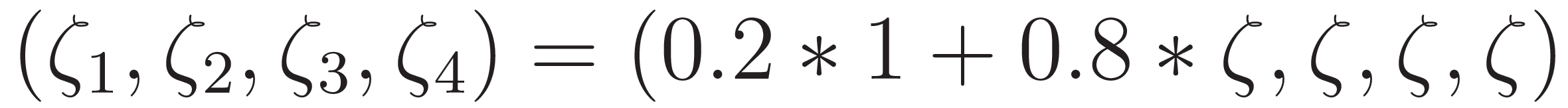
<http://www.sciweavers.org/tex2img.php?bc=Transparent&fc=Black&im=jpg&fs=100&ff=modern&edit=0&eq=(%5Czeta_1%2C%20%5Czeta_2%2C%20%5Czeta_3%2C%20%5Czeta_4)%20%3D%20(0.2*1%20%2B%200.8*%5Czeta%2C%20%5Czeta%2C%20%5Czeta%2C%20%5Czeta)#0> was the age-specific susceptibility based on the susceptibility parameter
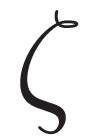
<http://www.sciweavers.org/tex2img.php?bc=Transparent&fc=Black&im=jpg&fs=100&ff=modern&edit=0&eq=%5Czeta#0>, as we assumed all children aged under 1 year old (i.e. 20% of the youngest model age group) were fully susceptible to influenza infection.

#### d. Vaccination coverage

Vaccine doses were assumed to have been distributed before any epidemic started, instead of during an epidemic. Therefore, vaccination coverage determined the size of compartments S, Sv and Rev at the start of each epidemic period, where the distribution of vaccinated individuals between Sv and Rev was determined by age-specific vaccine efficacy and annual hemisphere-specific matching of vaccines and circulating strains. No further vaccinations or loss of immunity occurred in the inference model.

The following sources detail seasonal influenza vaccination coverage from 2010-2019 in the exemplar countries.

1. **Argentina**

Urueña et al. (2021) [7] summarises coverage levels in the age groups 6mo - 2Y, 2 - 4Y, 5 - 14Y, 15 - 64Y, 65Y+. By recategorising the coverage levels into our model age groups, we determined age-specific coverage levels of:

0-4: (0*1/2 + 0.75*1.5 + 0.83*3)/5 = 72.3%

5-19: 50%

20-64: 50%

65+: 55%

These coverage levels closely agree with the limited coverage levels reported in a 2013 National Report published by the Health Ministry [8]:

20-64: (5*0.580 + 10*0.535 + 15*0.458 + 15*0.447)/45 = 48.5%

>65: 55.3%

2. **Australia**

We used data from the Australian Immunisation Register for the number of vaccinations given between 1st March and 3rd October 2023 [9] to estimate vaccination coverage levels:

0-5: 493688 /1517693 = 32.5%

5-64: 5553632/20425419 = 27.2%

65+: 3252809/ 4518800 = 72.0%

These coverage levels are approximately consistent with the 2009 Adult Vaccination Survey, which reported 22.8% coverage in the 18-64 age group, and 74.6% in the 65+ age group [10]. We therefore used vaccination coverage levels of 0.32 (0-4), 0.30 (5-19), 0.23 (20-64), 0.72 (65+).

3. **Canada**

Some Canadian provinces (but not all) have introduced universal influenza vaccination [11,12], meaning that seasonal influenza vaccination coverage is likely to vary between provinces. Due to the complexity of incorporating this into our national-level model, we used national-level coverage estimates.

We used reported 2013-2014 vaccination coverage levels where possible [13], providing estimates of coverage in the 12-17 age group (23%), 18-64 age group ((0.17*17 + 0.22*10 + 0.32*20)/47 = 24%) and the 65+ age group (64%). We assumed coverage levels in children under the age of 5 to be similar to in other HICs, so assumed a vaccination coverage level of 32% in the age group 0-4.

4. **China**

There is no routine influenza vaccination program in China, and a national cross-sectional survey found an overall influenza vaccination coverage of 2.4% in adults over the age of 40 in 2014-15 [14]. We therefore used 0% vaccination coverage at all ages in China between 2010 and 2019 for simplicity.

5. **Ghana**

There is currently no seasonal influenza vaccination program in Ghana [15]. We therefore assumed 0% vaccination coverage at all ages in Ghana between 2010 and 2019.

6. **Turkey**

A 2018 cross-sectional survey found that only 13.4% of the study population were occasionally vaccinated for influenza, and 8.1% received regular annual vaccination. For simplicity, we used 0% vaccination coverage at all ages in Turkey between 2010 and 2019.

7. **United Kingdom**

As the policy for seasonal influenza vaccination in the United Kingdom was changed in 2013 [16], we used two sets of vaccination coverage levels. 2010-2012 levels used were as in Baguelin et al. (2013) [2], and 2013-2019 levels as in Waterlow et al. (2023) [17]:

2010-2012:

0-5: (0.0011*0.979*1 + 0.031*0.021*1 + 0.0022*0.945*4 + 0.1827*0.055*4)/5 = 1.0%

5-20: (0.0048*10*0.902 + 0.223*10*0.098 + 0.0087*5*0.913 + 0.4728*5*0.087)/15 = 3.4%

20-65: (0.0087*0.913*5 + 0.4728*0.087*5 + 0.0172*0.908*20 + 0.486*0.092*20 + 0.0789*0.817*15 + 0.486*0.183*15)/45 = 8.3%

65+: 73.5%

We assumed coverage levels to be 0% in the 0-5 and 5-20 age groups for simplicity.

2013-2019:

0-5: (0.449*0.021*1 + 0.438*0.945*4 + 0.449*0.055*4)/5 = 35.3%

5-20: (0.604*10*0.902 + 0.449*10*0.098 + 0.604*2*0.913 + 0.449*5*0.087)/15 = 47.9%

20-65: (0*0.91*30 + 0.449*0.09*30 + 0.352*0.817*15 + 0.449*0.183*15)/45 = 15.0%

65+: 72.4%

| **Country** | **Years** | **0-4** | **5-19** | **20-64** | **65+** |
| --- | --- | --- | --- | --- | --- |
| **Argentina** | 2010 - 2019 | 0.72 | 0.50 | 0.50 | 0.55 |
| **Australia** | 2010 - 2019 | 0.32 | 0.30 | 0.23 | 0.72 |
| **Canada** | 2010 - 2019 | 0.32 | 0.23 | 0.24 | 0.64 |
| **China** | 2010 - 2019 | 0.00 | 0.00 | 0.00 | 0.00 |
| **Ghana** | 2010 - 2019 | 0.00 | 0.00 | 0.00 | 0.00 |
| **Turkey** | 2010 - 2019 | 0.00 | 0.00 | 0.00 | 0.00 |
| **United Kingdom** | 2010 - 2012 | 0.00 | 0.00 | 0.08 | 0.73 |
|  | 2013 - 2019 | 0.35 | 0.48 | 0.15 | 0.73 |

**Table S4:** Vaccination coverage levels used for inference in exemplar countries between 2010 and 2019 in each of the model age groups.

#### e. Matching of years

Estimated annual vaccine efficacy in the Northern and Southern Hemisphere between 2010 and 2019 using peer-reviewed literature. Based on either analysis of the antigenic similarity between circulating virus strains and the vaccine strains, or estimates of vaccine efficacy, we determined the years in which the vaccine strains were likely to have matched or mismatched circulating strains of influenza A and B in the Northern and Southern Hemisphere.

| **Year** | **Northern Hemisphere** | | | **Southern Hemisphere** | | |
| --- | --- | --- | --- | --- | --- | --- |
|  | **Influenza A** | **Influenza B** | **Source** | **Influenza A** | **Influenza B** | **Source** |
| **2010** | M | M | [18] | M | M | [19,20] |
| **2011** | U | M | [21] | M | M | [22] |
| **2012** | U | M | [23] | M | M | [24] |
| **2013** | M | M | [25,26] | M | M | [27] |
| **2014** | U | U | [28] | M | M | [29,30] |
| **2015** | M | M | [31] | U | M | [32] |
| **2016** | U | M | [33,34] | M | U | [35,36] |
| **2017** | U | U | [37] | U | M | [38,39] |
| **2018** | M | M | [40,41] | M | M | [42,43] |
| **2019** | M | M | [44] | U | M | [45,46] |

**Table S5:** Matching (M) and mismatched (U) vaccinations in each year of the inference period, for influenza A and B, in both hemispheres.

#### f. The MCMC and inference model

We estimated the reporting rate, susceptibility, transmissibility, and the initial number of infections in each age group at the start of each identified epidemic period by fitting to the FluNet data using a binomial likelihood for the weekly number of reported cases. Parameters were estimated independently for each epidemic. Therefore, we did not assume the same influenza reporting rate between epidemics, strains, or countries. We used the Markov chain Monte Carlo (MCMC) algorithm in the *BayesianTools* R package [47] to obtain 10,000 joint samples of the parameters for each epidemic (with thinning of 50 and a 500,000 burn-in).

Priors

Transmissibility:
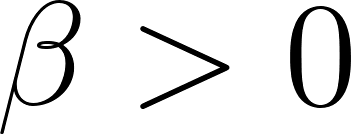
<https://www.codecogs.com/eqnedit.php?latex=%5Cbeta%20%3E%200#0>

Susceptibility:
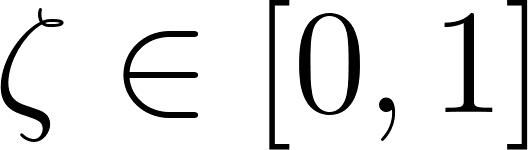
<https://www.codecogs.com/eqnedit.php?latex=%5Czeta%20%5Cin%20%5B0%2C1%5D#0>,
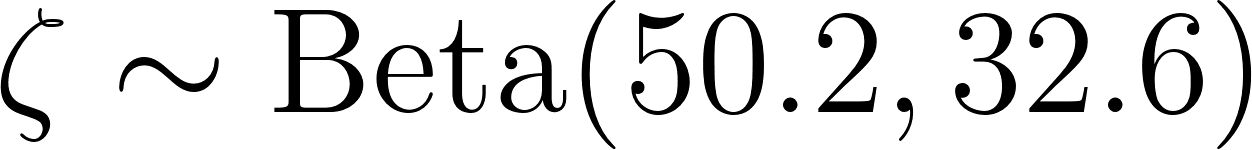
<https://www.codecogs.com/eqnedit.php?latex=%5Czeta%20%5Csim%20%5Ctext%7BBeta%7D(50.2%2C%2032.6)%20#0>

Initial infected:
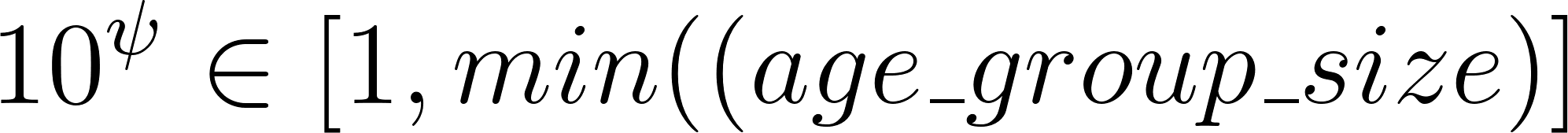
<https://www.codecogs.com/eqnedit.php?latex=10%5E%5Cpsi%20%5Cin%20%5B1%2C%20min(%5Ctext(age%5C_group%5C_size)%5D#0>

Reporting rate:
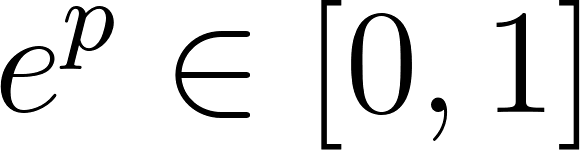
<https://www.codecogs.com/eqnedit.php?latex=e%5Ep%20%5Cin%20%5B0%2C1%5D#0>

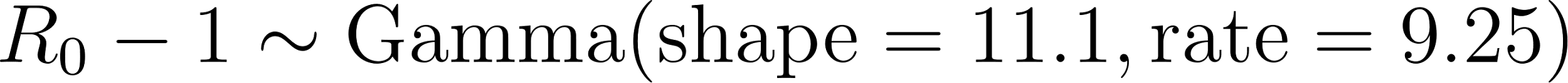
<https://www.codecogs.com/eqnedit.php?latex=R_0%20-%201%20%5Csim%20%5Ctext%7BGamma%7D(%5Ctext%7Bshape%7D%20%3D%2011.1%2C%20%5Ctext%7Brate%7D%20%3D%209.25)%20#0>

### 4. Inference outputs

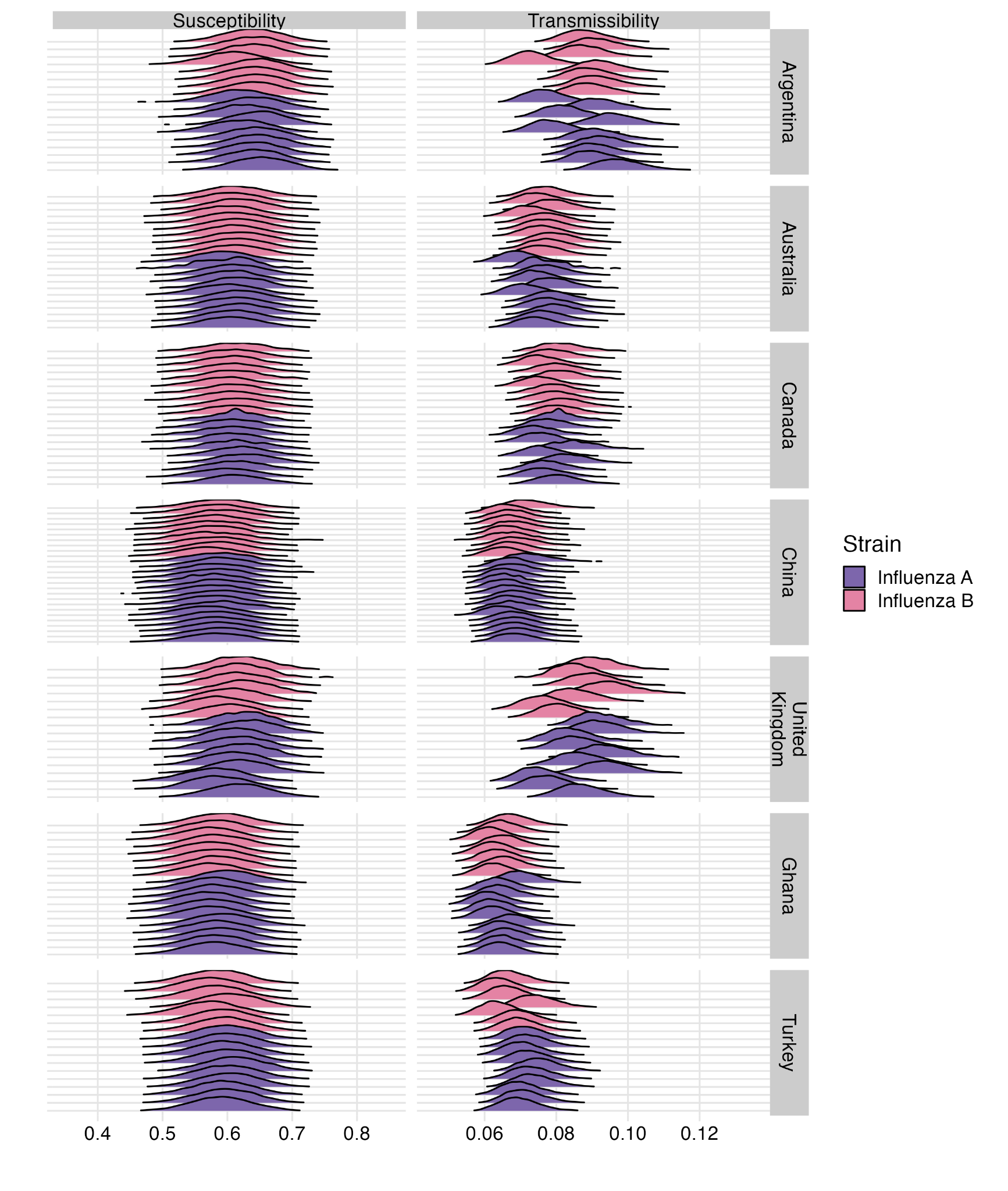

**Figure S4:** Posterior distributions of population-level susceptibility and influenza transmissibility in each epidemic used for inference.

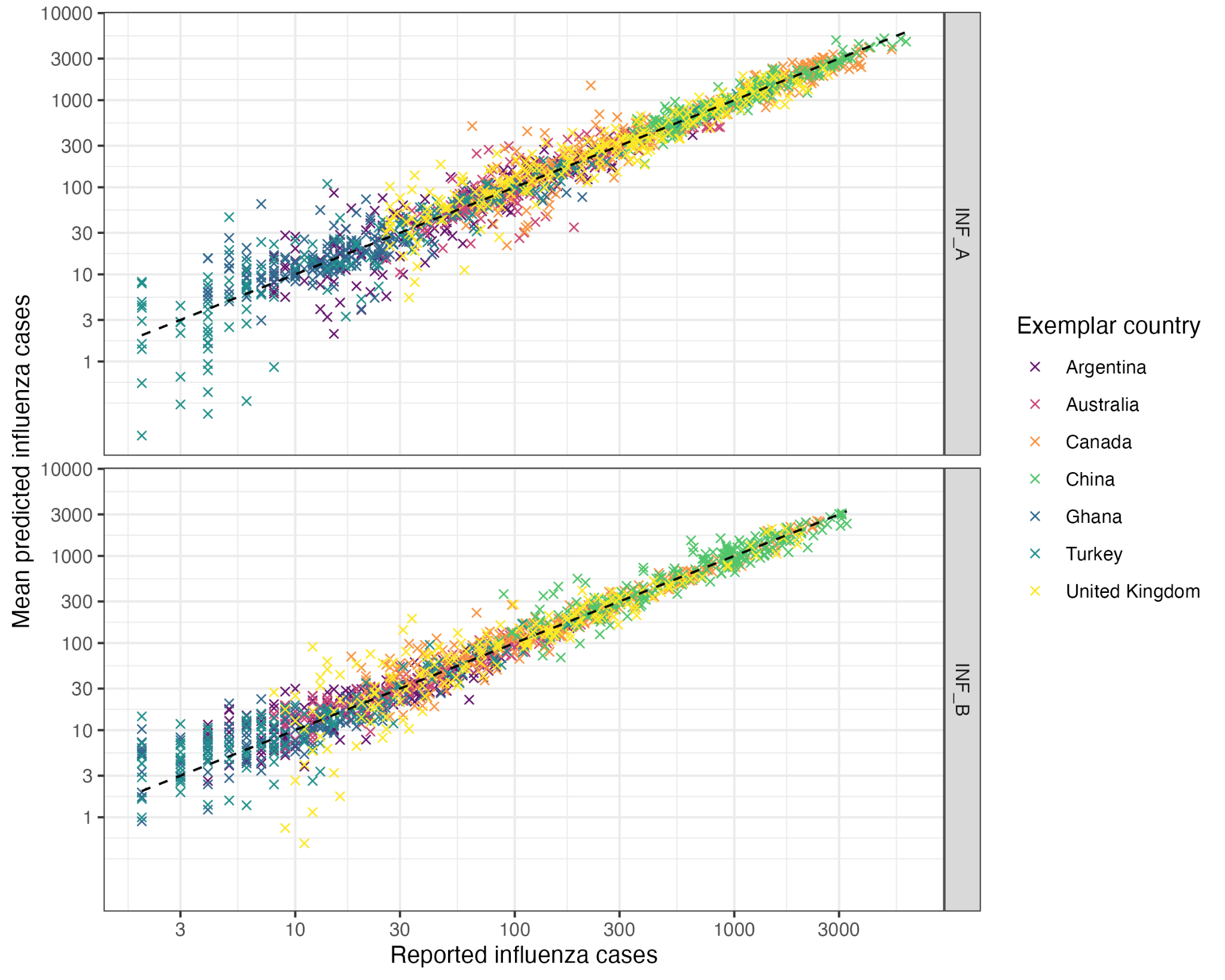

**Figure S5:** Validation of inference model’s goodness of fit, comparing reported influenza cases and mean predicted influenza cases across all epidemics, stratified by influenza strains and using log scales on both axes. Dotted line indicates x=y.

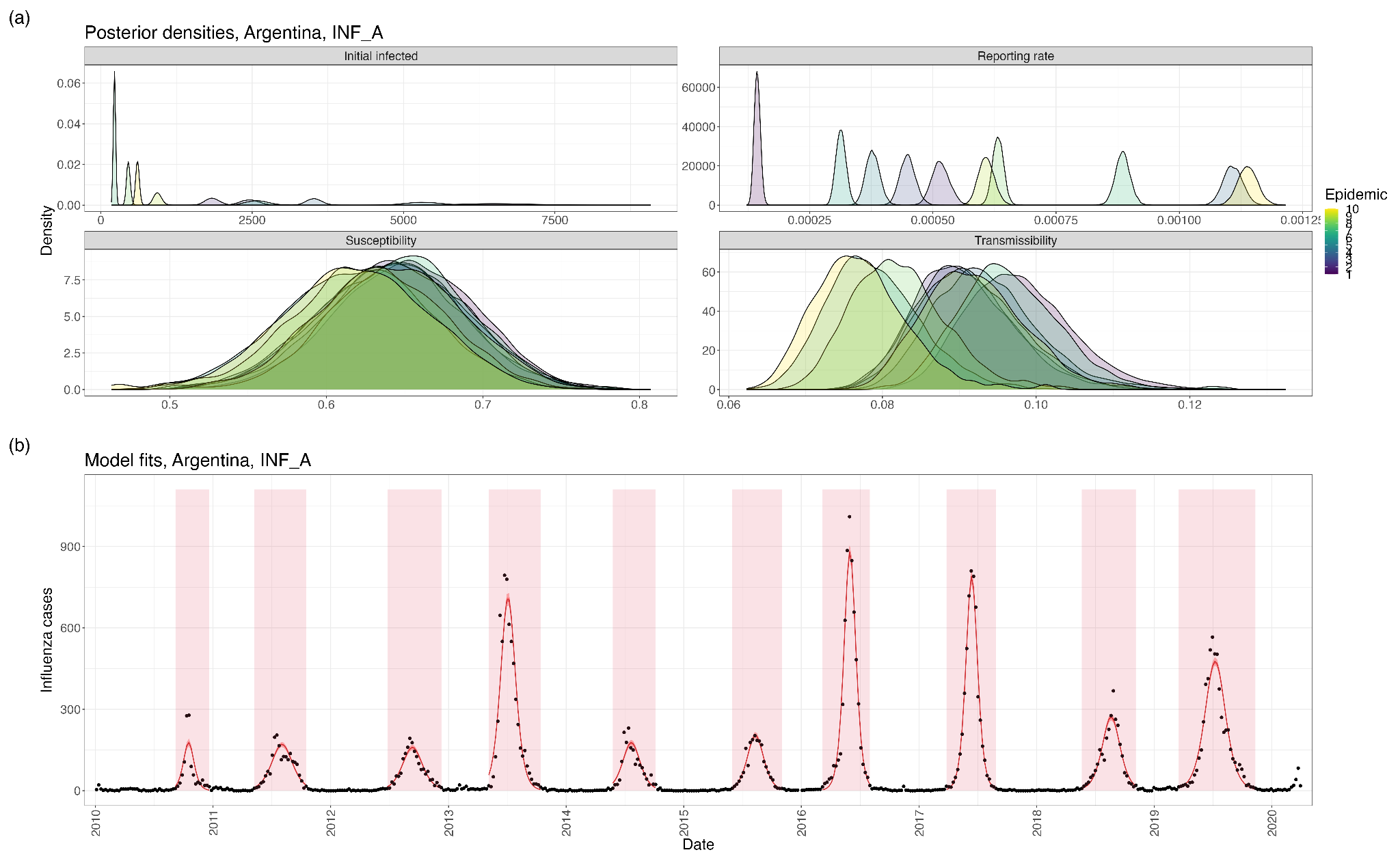

**Figure S6: (a)** Posterior distributions of the initial number of infections, reporting rate, population-level susceptibility, and influenza transmissibility in each epidemic used for inference (Argentina, Influenza A). **(b)** Model fits using parameter posteriors.

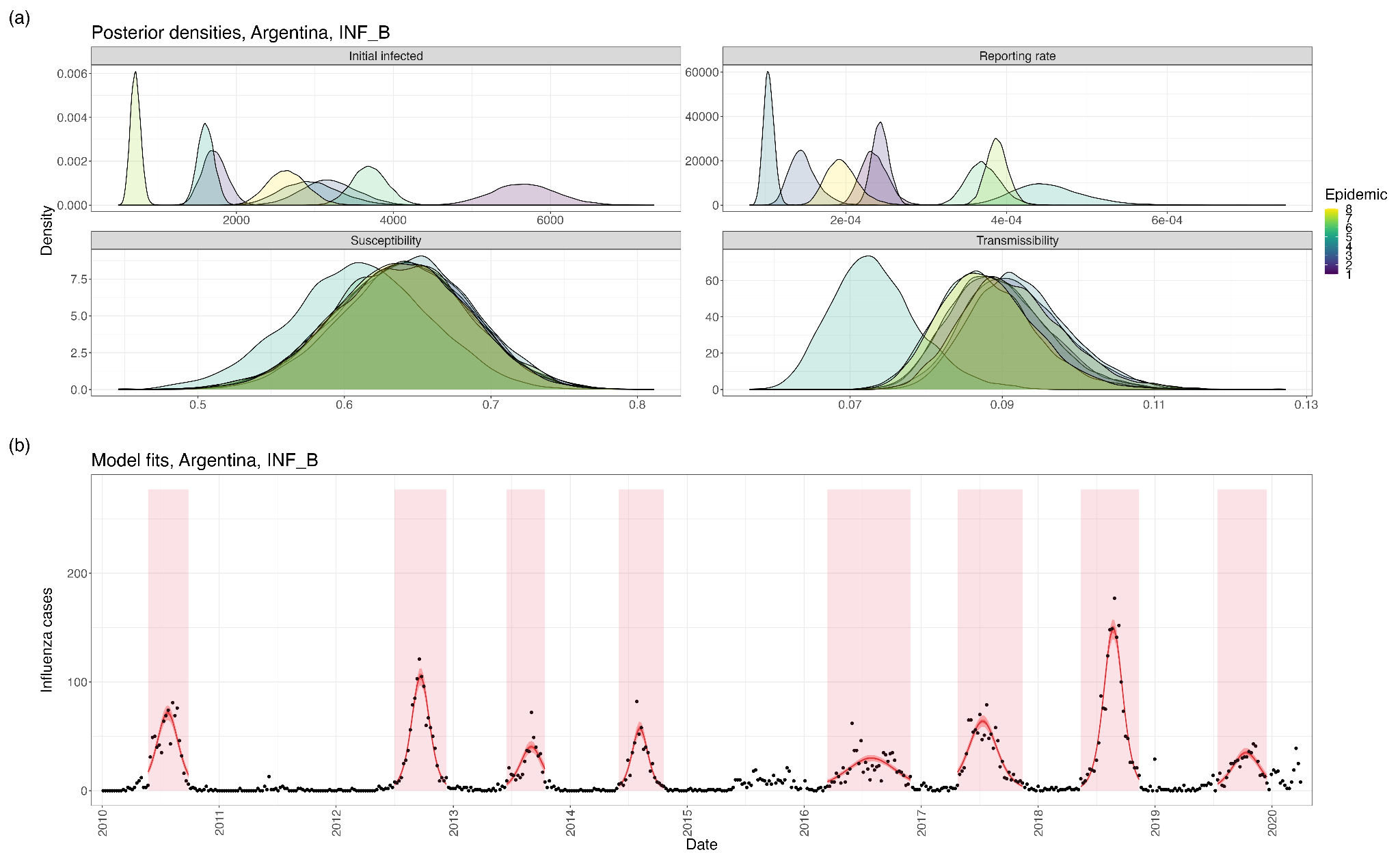

**Figure S7: (a)** Posterior distributions of the initial number of infections, reporting rate, population-level susceptibility, and influenza transmissibility in each epidemic used for inference (Argentina, Influenza B). **(b)** Model fits using parameter posteriors.

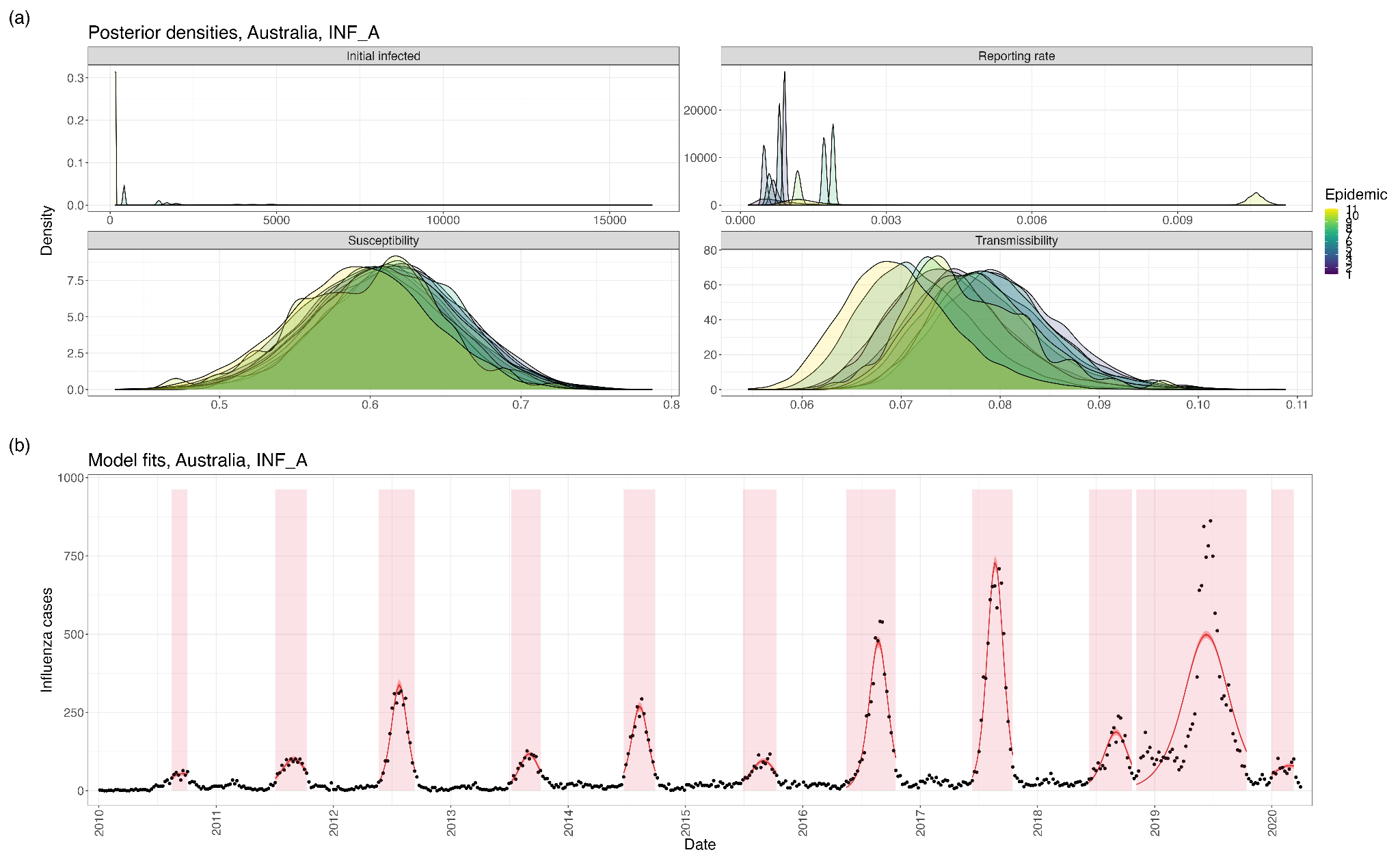

**Figure S8: (a)** Posterior distributions of the initial number of infections, reporting rate, population-level susceptibility, and influenza transmissibility in each epidemic used for inference (Australia, Influenza A). **(b)** Model fits using parameter posteriors.

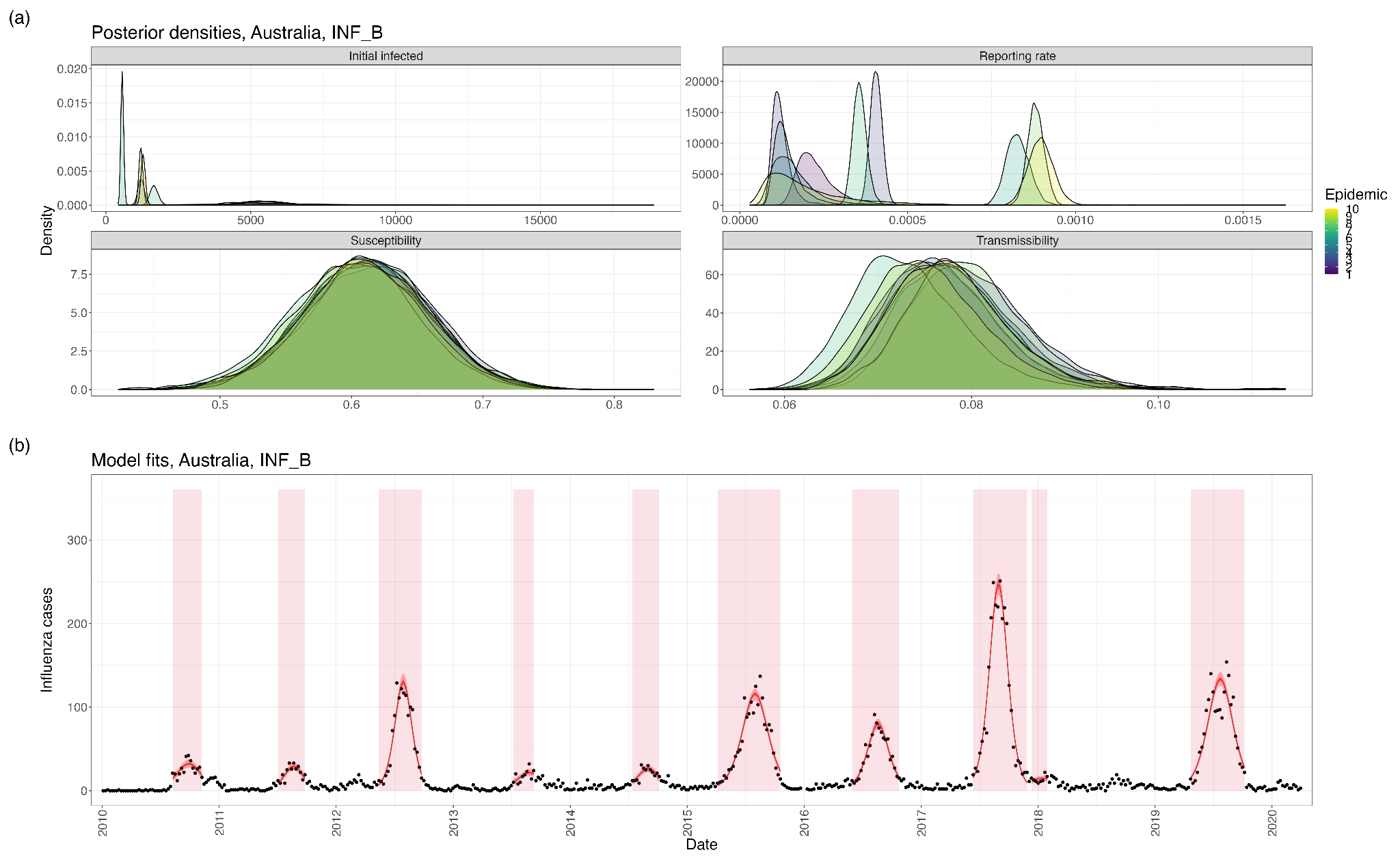

**Figure S9: (a)** Posterior distributions of the initial number of infections, reporting rate, population-level susceptibility, and influenza transmissibility in each epidemic used for inference (Australia, Influenza B). **(b)** Model fits using parameter posteriors.

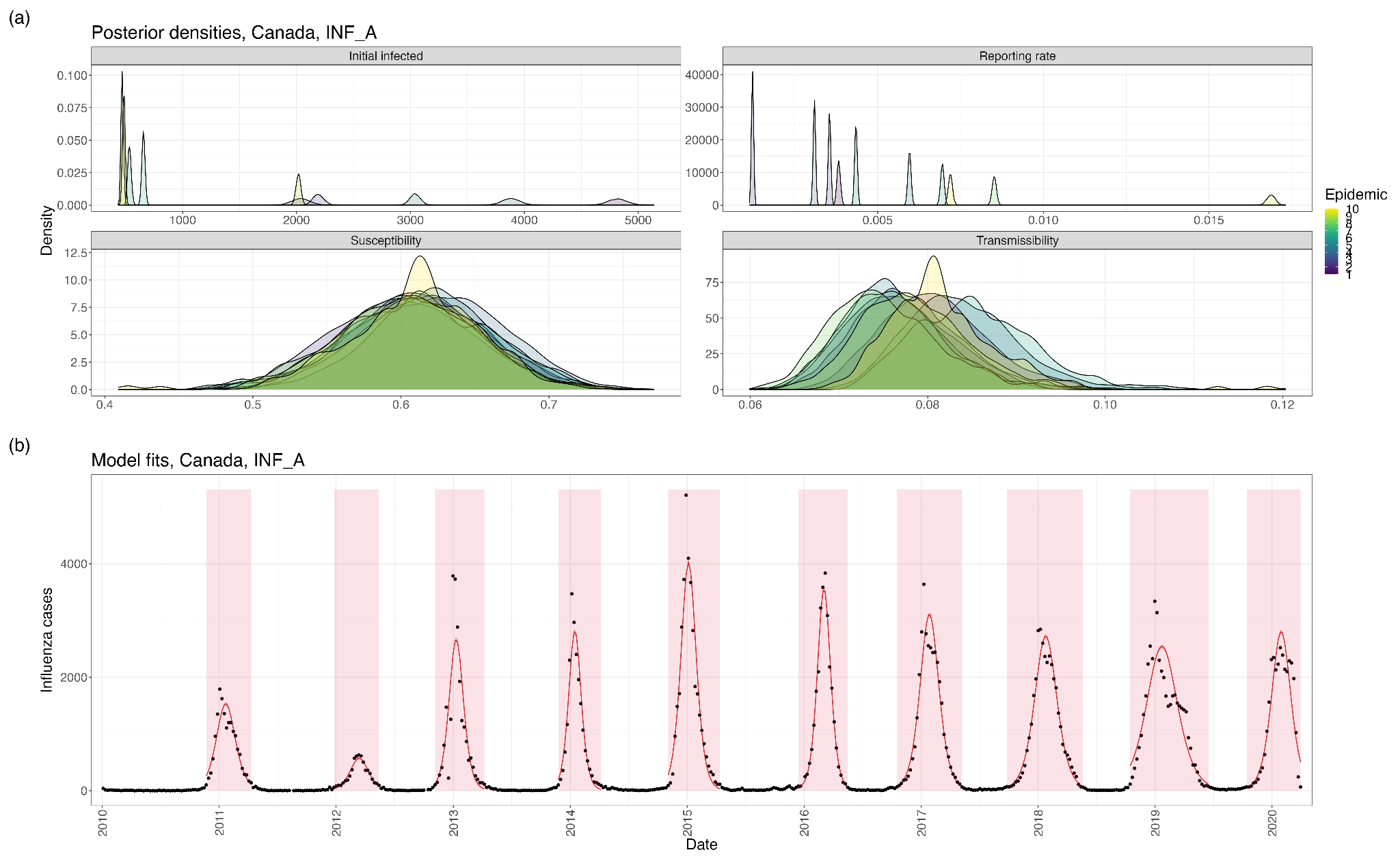

**Figure S10: (a)** Posterior distributions of the initial number of infections, reporting rate, population-level susceptibility, and influenza transmissibility in each epidemic used for inference (Canada, Influenza A). **(b)** Model fits using parameter posteriors.

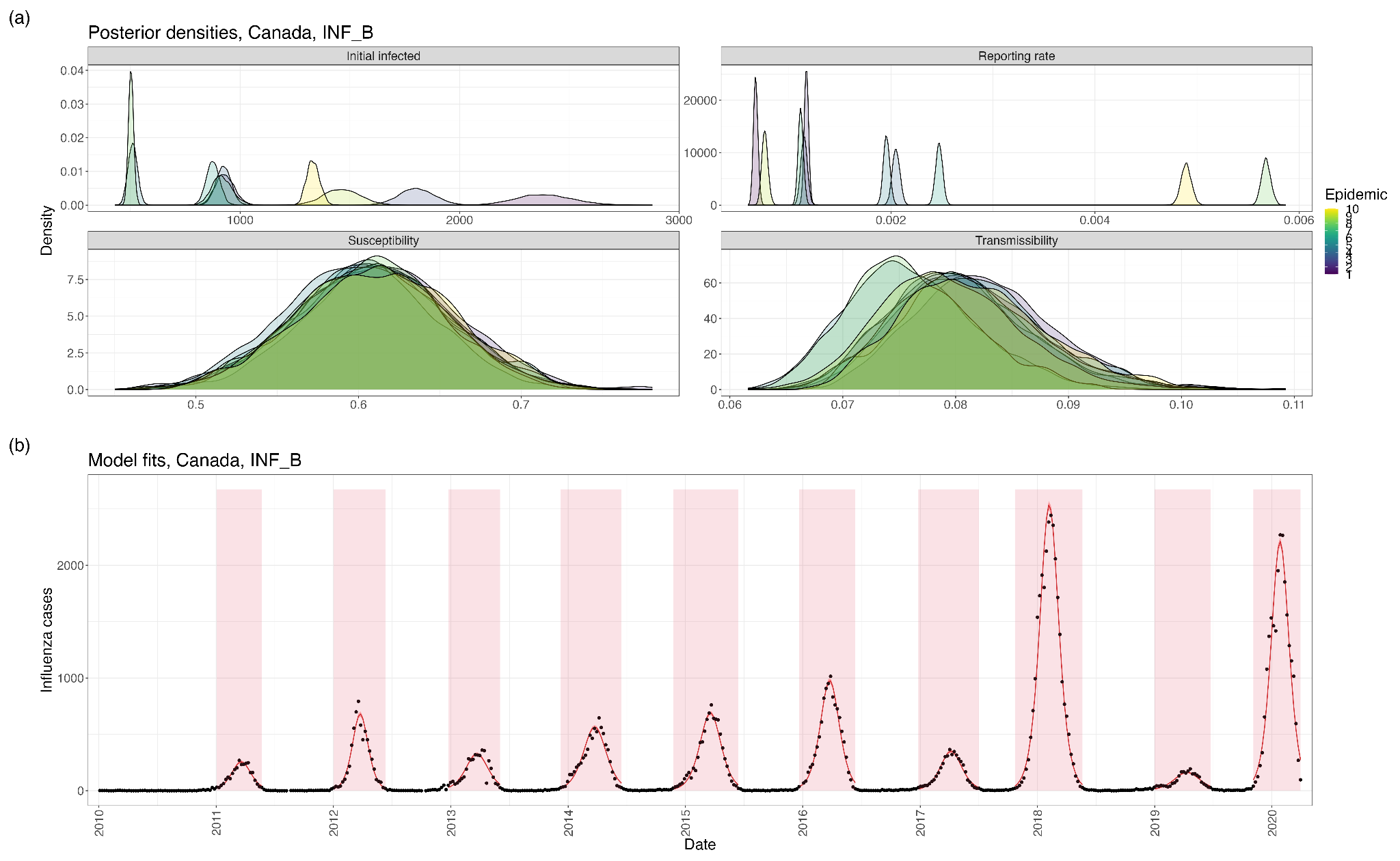

**Figure S11: (a)** Posterior distributions of the initial number of infections, reporting rate, population-level susceptibility, and influenza transmissibility in each epidemic used for inference (Canada, Influenza B). **(b)** Model fits using parameter posteriors.

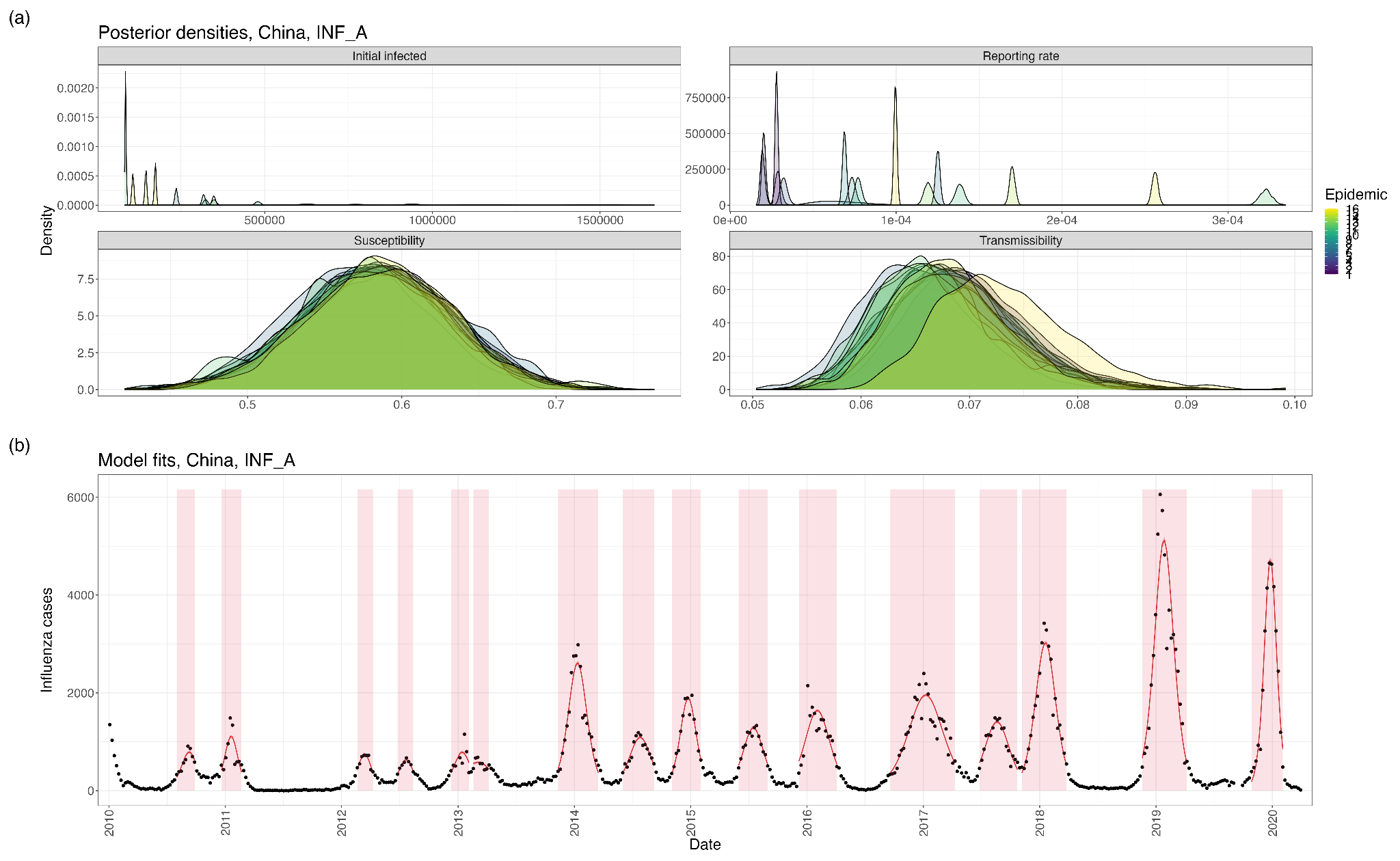

**Figure S12: (a)** Posterior distributions of the initial number of infections, reporting rate, population-level susceptibility, and influenza transmissibility in each epidemic used for inference (China, Influenza A). **(b)** Model fits using parameter posteriors.

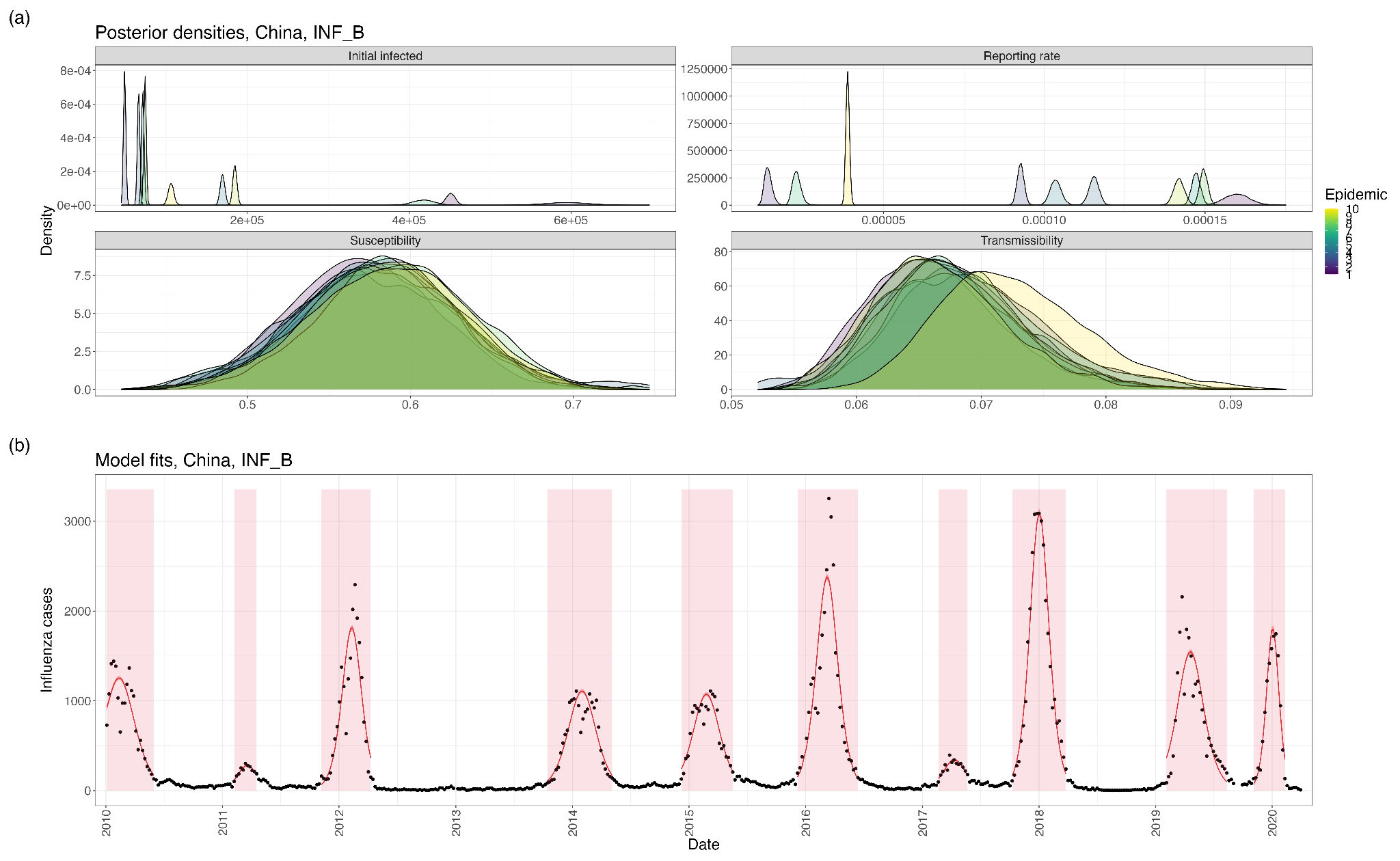

**Figure S13: (a)** Posterior distributions of the initial number of infections, reporting rate, population-level susceptibility, and influenza transmissibility in each epidemic used for inference (China, Influenza B). **(b)** Model fits using parameter posteriors.

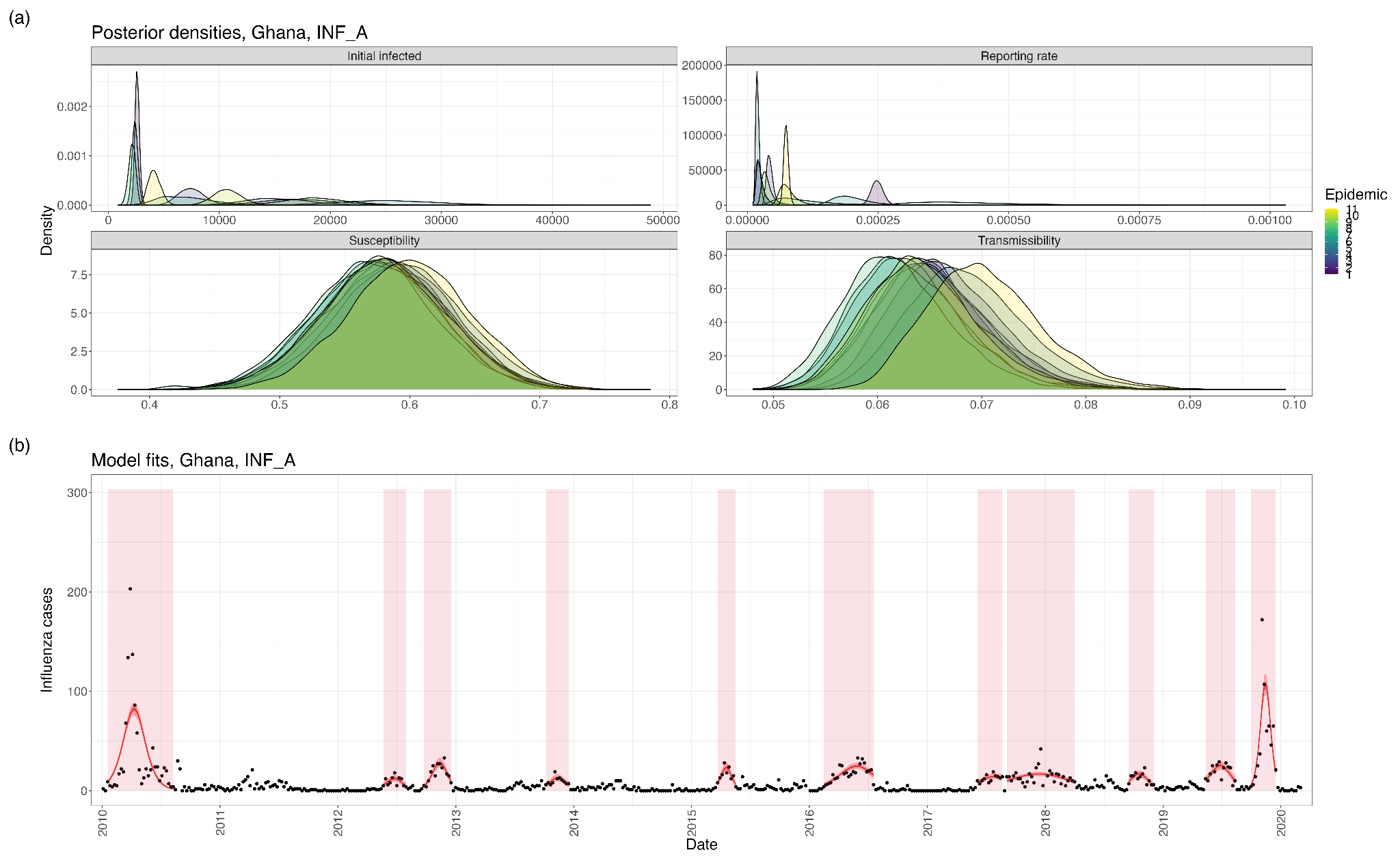

**Figure S14: (a)** Posterior distributions of the initial number of infections, reporting rate, population-level susceptibility, and influenza transmissibility in each epidemic used for inference (Ghana, Influenza A). **(b)** Model fits using parameter posteriors.

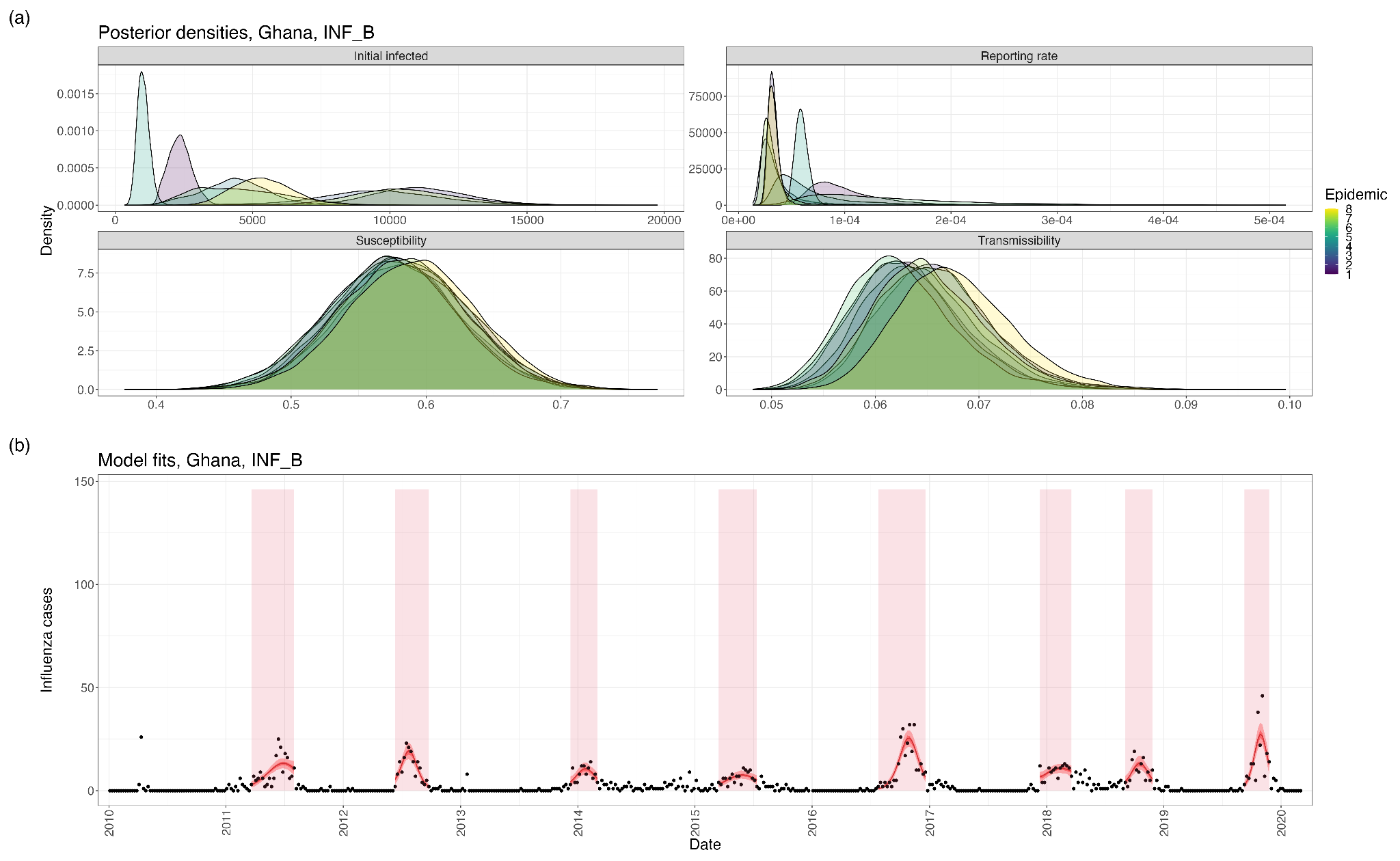

**Figure S15: (a)** Posterior distributions of the initial number of infections, reporting rate, population-level susceptibility, and influenza transmissibility in each epidemic used for inference (Ghana, Influenza B). **(b)** Model fits using parameter posteriors.

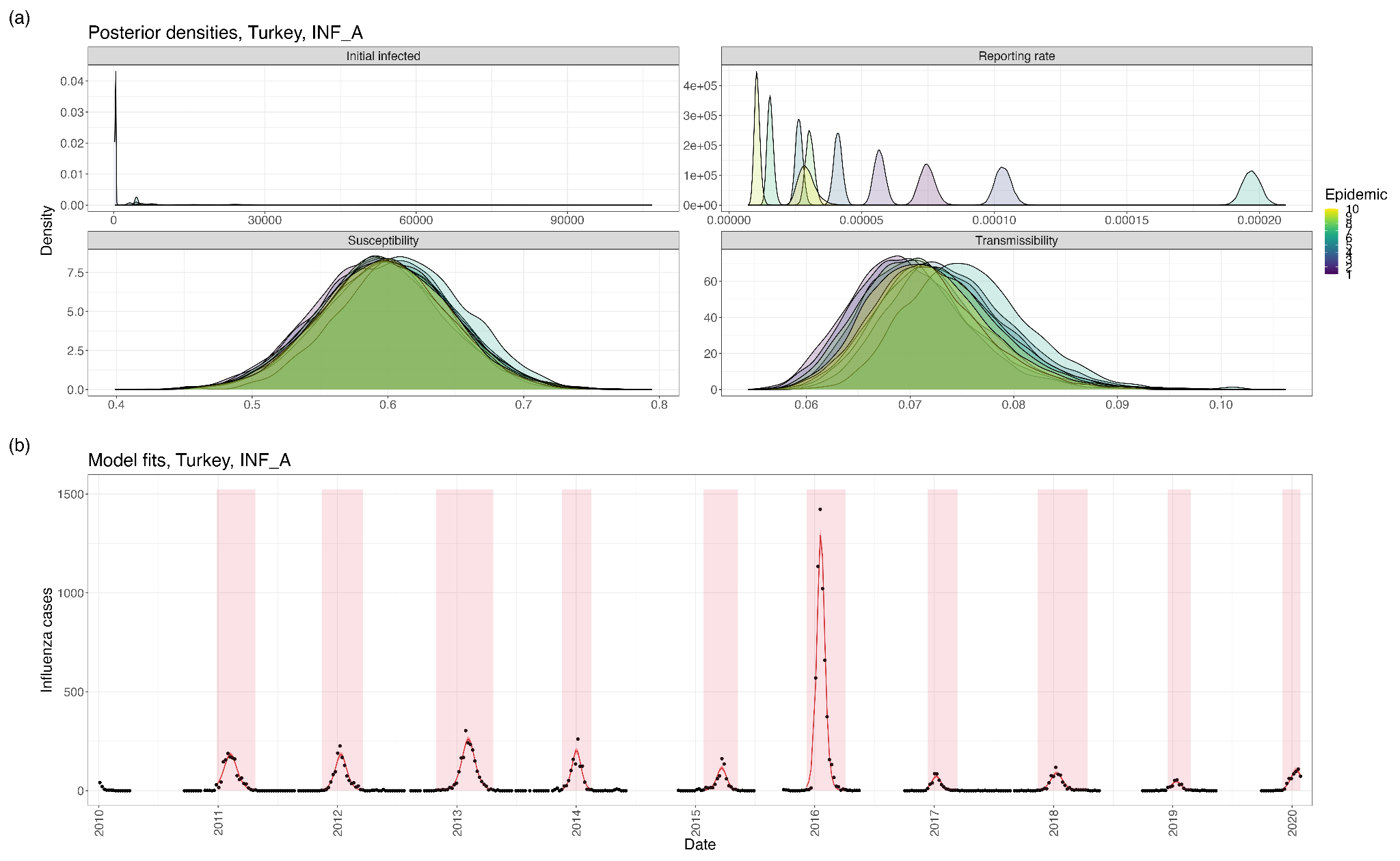

**Figure S16: (a)** Posterior distributions of the initial number of infections, reporting rate, population-level susceptibility, and influenza transmissibility in each epidemic used for inference (Turkey, Influenza A). **(b)** Model fits using parameter posteriors.

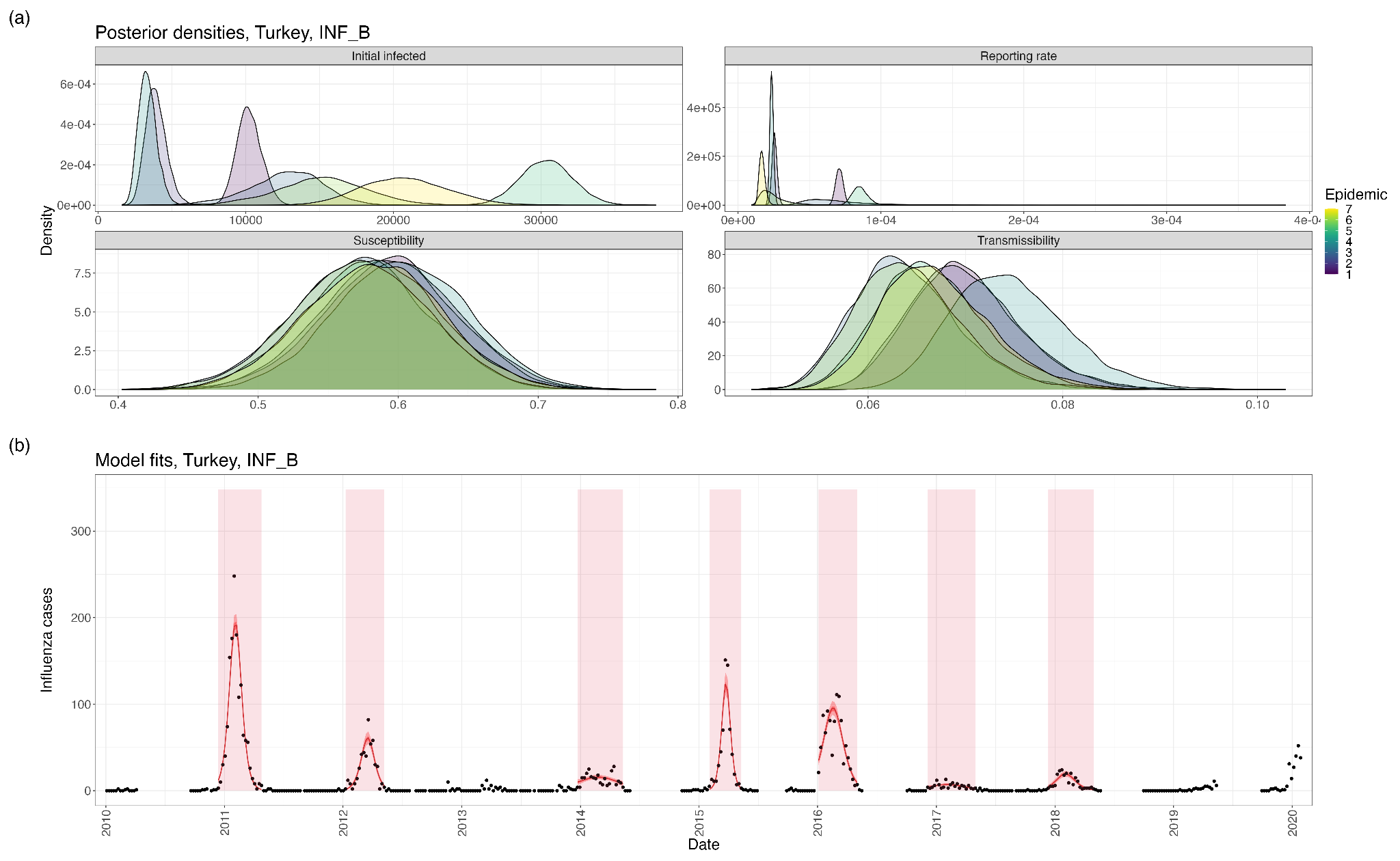

**Figure S17: (a)** Posterior distributions of the initial number of infections, reporting rate, population-level susceptibility, and influenza transmissibility in each epidemic used for inference (Turkey, Influenza B). **(b)** Model fits using parameter posteriors.

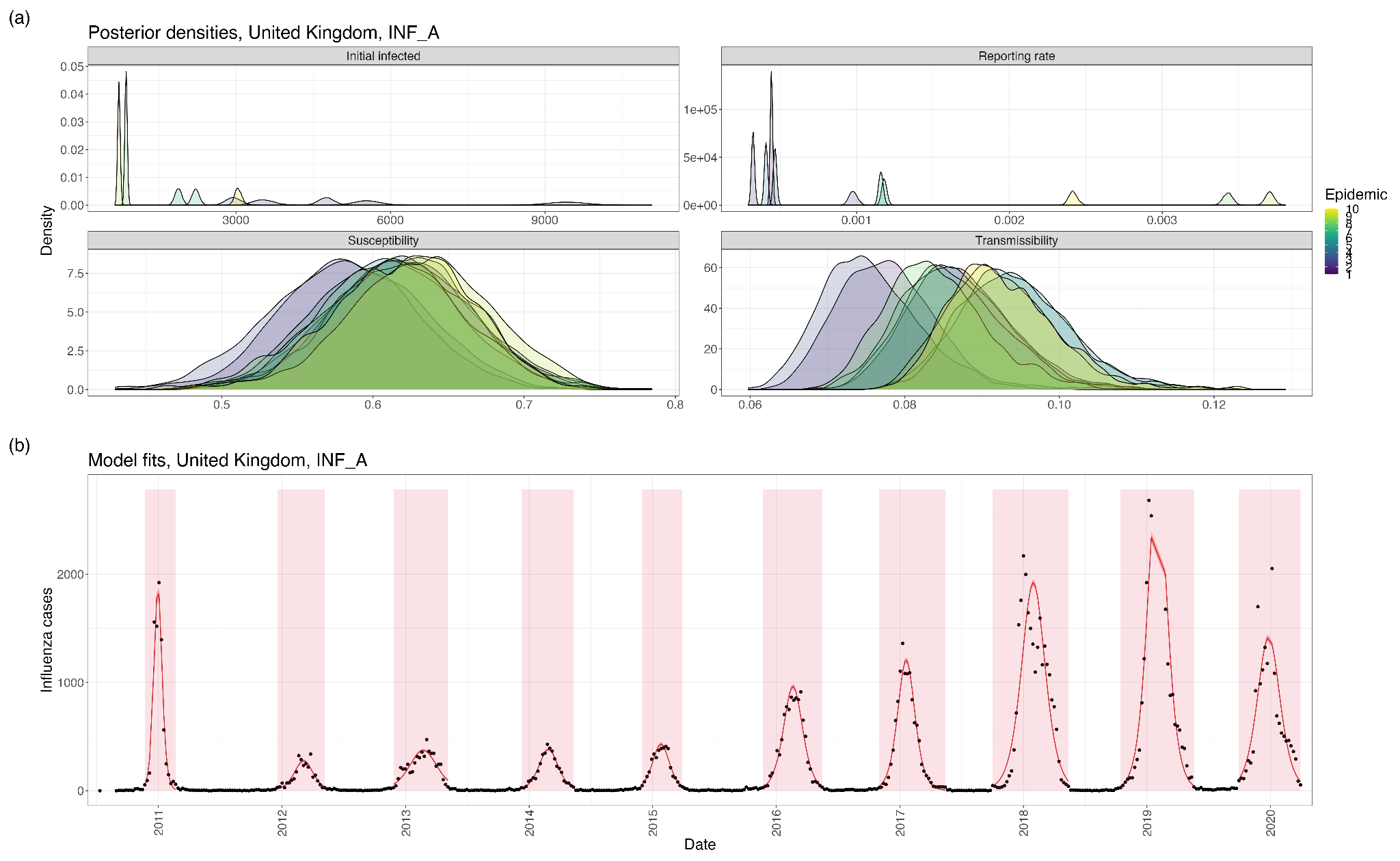

**Figure S18: (a)** Posterior distributions of the initial number of infections, reporting rate, population-level susceptibility, and influenza transmissibility in each epidemic used for inference (United Kingdom, Influenza A). **(b)** Model fits using parameter posteriors.

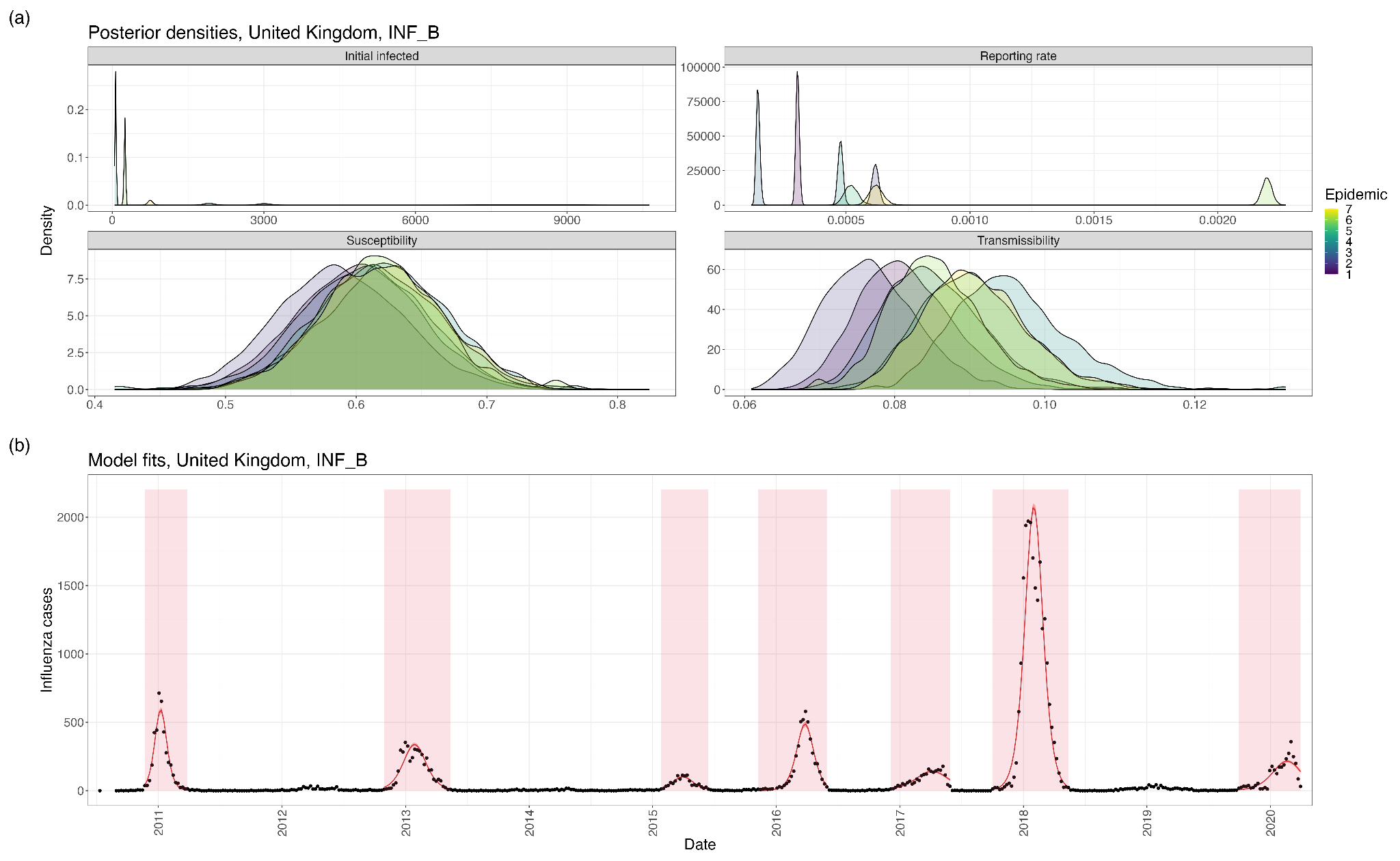

**Figure S19: (a)** Posterior distributions of the initial number of infections, reporting rate, population-level susceptibility, and influenza transmissibility in each epidemic used for inference (United Kingdom, Influenza B). **(b)** Model fits using parameter posteriors.

### 5. Simulating epidemics

#### a. Demographic changes

Each simulation used the 2025 age-specific population, extracted from the World Population Prospects 2022 [48]. We aged each country-specific population annually (for example 1/5 of the [0,4] age group moves into the [5,19] age group each year), and simulated any epidemics occurring in a given calendar year on the population defined at the start of that year. We removed a proportion of each age group from the model to reflect age-specific mortality rates, and assumed in the model that mortality occurred independently of vaccination states or influenza infection status. We also introduced a number of births into the youngest age group, at a rate proportional to the crude birth rate (CBR), and assumed all newborns to be susceptible. The country-specific rates at which the births and deaths occurred were based on data from World Population Prospects 2022 [48], and fixed at the projected 2025 values:

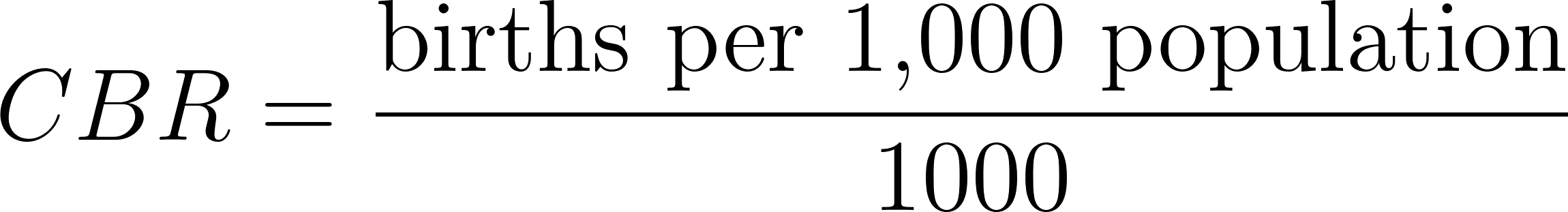
<https://www.codecogs.com/eqnedit.php?latex=CBR%20%3D%20%5Cfrac%7B%5Ctext%7Bbirths%20per%201%2C000%20population%7D%7D%7B1000%7D#0>

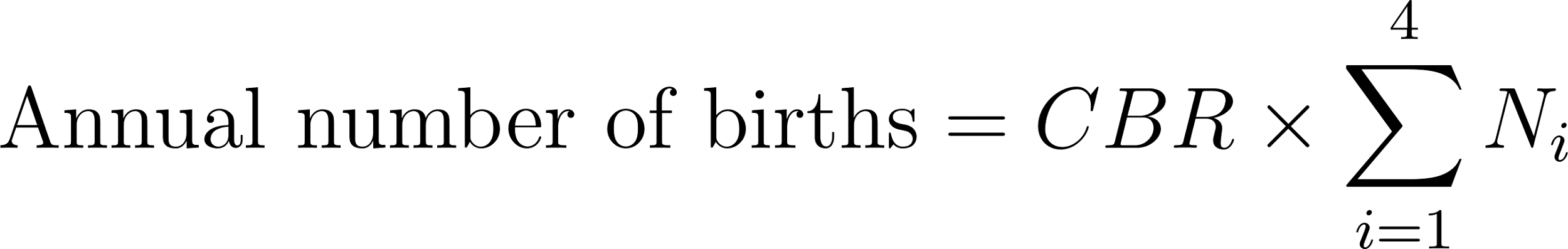
<https://www.codecogs.com/eqnedit.php?latex=%5Ctext%7BAnnual%20number%20of%20births%7D%20%3D%20CBR%20%5Ctimes%20%5Csum_%7Bi%3D1%7D%5E4%20N_i#0>

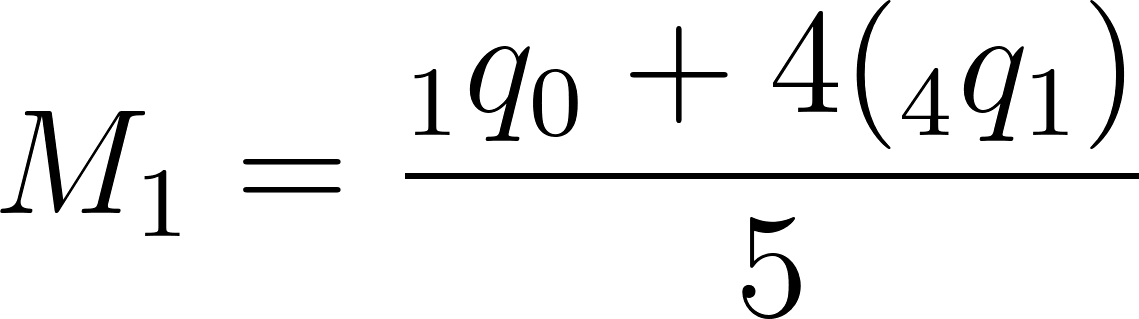
<https://www.codecogs.com/eqnedit.php?latex=M_1%20%3D%20%5Cfrac%7B%7B_1q_0%7D%20%2B%204(%7B_4q_1%7D)%7D%7B5%7D#0>

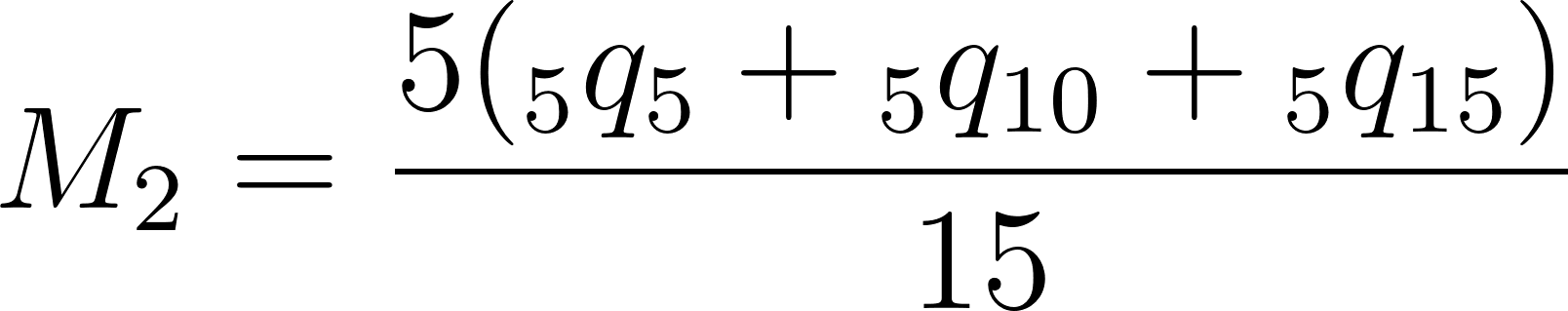
<https://www.codecogs.com/eqnedit.php?latex=M_2%20%3D%20%5Cfrac%7B5%20(%7B_5q_5%7D%20%2B%20%7B_5q_%7B10%7D%7D%20%2B%20%7B_5q_%7B15%7D%7D)%7D%7B15%7D%20#0>

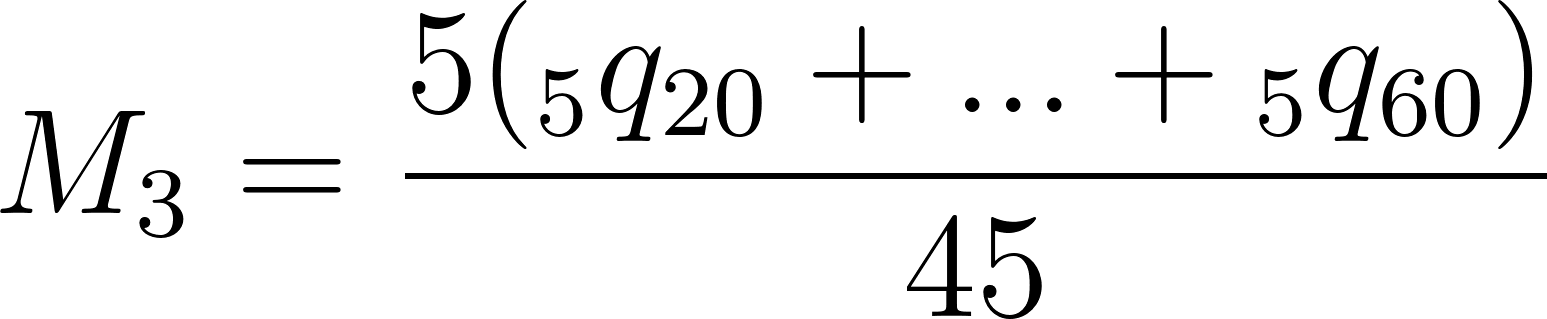
<https://www.codecogs.com/eqnedit.php?latex=M_3%20%3D%20%5Cfrac%7B5%20(%7B_5q_%7B20%7D%7D%20%2B%20.%20.%20.%20%2B%20%7B_5q_%7B60%7D%7D)%7D%7B45%7D%20#0>

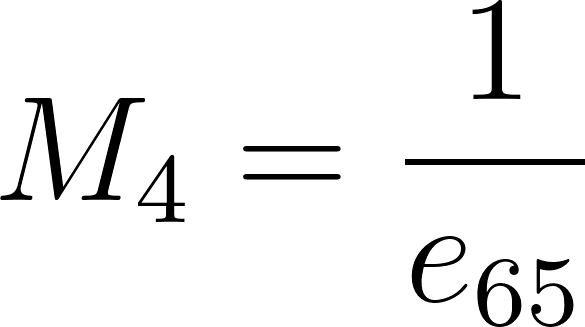
<https://www.codecogs.com/eqnedit.php?latex=M_4%20%3D%20%5Cfrac%7B1%7D%7Be_%7B65%7D%7D#0>

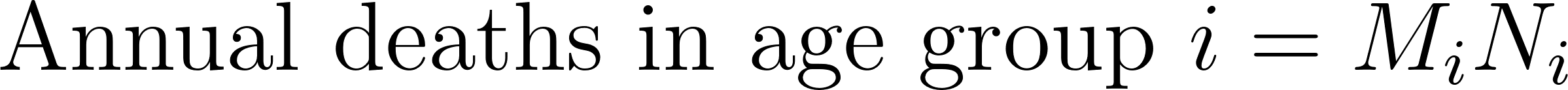
<https://www.codecogs.com/eqnedit.php?latex=%5Ctext%7BAnnual%20deaths%20in%20age%20group%20%7D%20i%20%3D%20M_i%20N_i%20#0>

Where
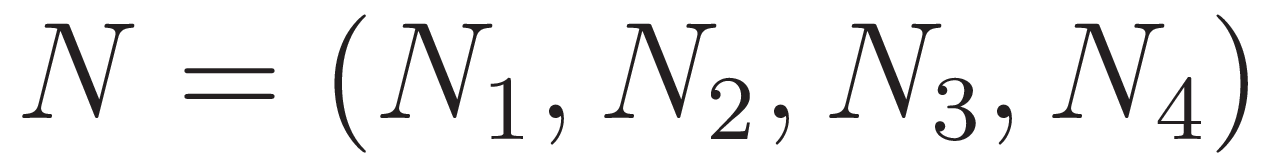
<http://www.sciweavers.org/tex2img.php?bc=Transparent&fc=Black&im=jpg&fs=100&ff=modern&edit=0&eq=N%20%3D%20(N_1%2C%20N_2%2C%20N_3%2C%20N_4)#0> are the population sizes of the four model age groups in a given year,
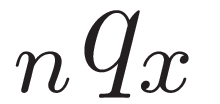
<http://www.sciweavers.org/tex2img.php?bc=Transparent&fc=Black&im=jpg&fs=100&ff=modern&edit=0&eq=%7B_nq_x%7D#0> is the probability of dying in the age interval [x, x+n),
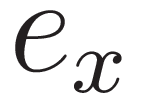
<http://www.sciweavers.org/tex2img.php?bc=Transparent&fc=Black&im=jpg&fs=100&ff=modern&edit=0&eq=e_x#0> is remaining life expectancy at age x, and
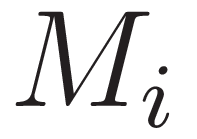
<http://www.sciweavers.org/tex2img.php?bc=Transparent&fc=Black&im=jpg&fs=100&ff=modern&edit=0&eq=M_i#0> is the fixed annual mortality rate in model age group i. We assumed a rectangular age structure within each model age group and hence equally weighted all mortality rates in each age group.

In the Northern Hemisphere, demographic changes occurred annually on 1st April, similarly 1st October in the Southern Hemisphere. We assumed that there was no in- or out-migration, for simplicity.

#### b. Contact matrices

We produced the contact matrices for the epidemic simulation by applying the same process as in the inference (Supplementary Section 3b): we reweighted the country-specific contact matrices derived in Prem et al. [1] to be reciprocal with respect to the annual projected demography, and divided by the age-specific populations to produce a per-capita probability of contact between two age-specific individuals.

For the 9 countries not included in Prem et al.’s output, we substituted the underlying contact matrices for their closest geographical neighbour (Australia to New Zealand, Somalia to Ethiopia, Lebanon to Syria, Japan to South Korea, Taiwan to China, Kosovo for Serbia, New Caledonia to Vanuatu, French Guiana to Suriname, and Haiti to the Dominican Republic).

Contact matrices were then rescaled in magnitude for each epidemic simulation to match the epidemic’s original R_0_ from inference in the exemplar country (see Supplementary Section 6f for discussion).

#### c. Epidemic timing

The epidemic identification from FluNet data only identified the period of each epidemic with the highest number of infections, and tended to exclude the start and end of epidemics as the reported number of infections became low. In order to capture whole epidemics and any effects of vaccination during the period in which the epidemic is growing, we calculated the date on which the epidemic had 10 infections in each age group in order to produce the epidemic peak inferred from our analysis. We did this by simulating the epidemic from 10 infections in each age group using the epidemic posterior parameter samples, calculating the number of weeks that the simulated epidemic took to reach its peak, and pushing back the start date of each epidemic by the appropriate number of weeks. If the new start date was before the start of the inference period (1st January 2025) or the most recent ‘ageing date’, the epidemic started on that date and the number of infected individuals on the start date was calculated, so that the epidemic peaks still match.

#### d. Epidemic simulation

For each country, the 100 sampled 30-year periods were simulated under each vaccination scenario (varying the vaccine type used, age-specific population coverage levels, and vaccination mechanisms). No background infections were assumed to occur in inter-epidemic periods.

In the first year, the intended vaccination coverage was achieved in each age group. In each following year, vaccine doses were distributed to each age group at the necessary rate to achieve intended coverage, based on the mean immunity duration of the vaccine type used. These vaccine doses were distributed randomly across the population, independent of previous vaccination history, as the immunity status of any recently vaccinated individual would be unknown. Therefore, some vaccine doses were given to individuals whose immunity had not waned; these vaccine doses were costed in the economic model but redundant in the vaccine impact model. Vaccinations were given over a twelve-week period, starting on October 1st (Northern Hemisphere) or April 1st (Southern Hemisphere). The ageing and vaccinations therefore occurred 6 months apart in each hemisphere. Where the targeted age groups for vaccination did not directly align with model age groups, we scaled coverage by the proportion of the age group being targeted by vaccination (i.e. using a rectangular age structure within each age group).

In each country, weekly infections were accumulated over each epidemic, stratified by age, vaccination status, and infecting strain. Annual total vaccine doses were also accumulated, stratified by age.

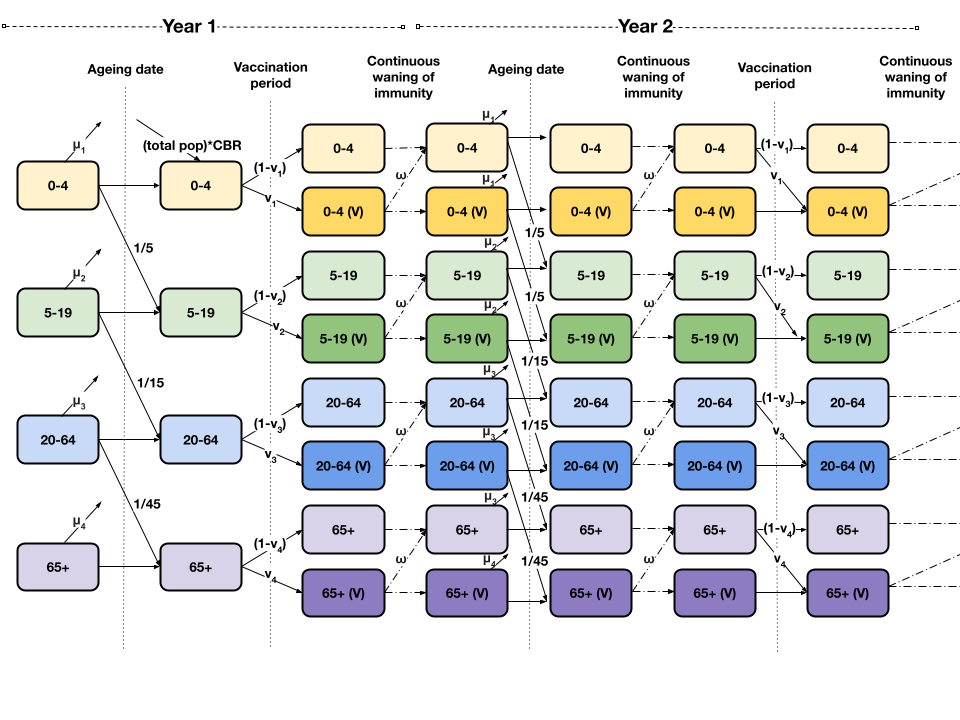

**Figure S20:** The vaccination model, example shown for the first two years of the simulation period. The whole population begins as unvaccinated. On the ageing date, individuals were removed from the model at age-specific mortality rates (μ_i_), and aged into the next age groups at rates proportional to their size. Susceptible newborns were introduced at a rate proportional to the crude birth rate (CBR). Over the vaccination period (12 weeks), individuals were moved into the vaccinated compartment at age-specific rates which depend on vaccination coverage and efficacy (v_i_). After the vaccination period, individuals lost their vaccine-induced immunity and moved back into the unvaccinated compartment at rate ω, which varies by vaccine type. The ageing, waning, and vaccination occurs again annually.

### 6. Simulation outputs

#### a. Population-level immunity

The annual vaccination program occurs for 12 weeks, beginning on 1st October in the Northern Hemisphere, or 1st April in the Southern Hemisphere. Annual ageing occurs on 1st April in the Northern Hemisphere (resp. 1st October in Southern Hemisphere), at which point 20% of individuals previously in the 0-4 age into the 5-19 age group (resp. for older age groups), non-influenza deaths occur, and susceptible newborns enter the 0-4 age group.

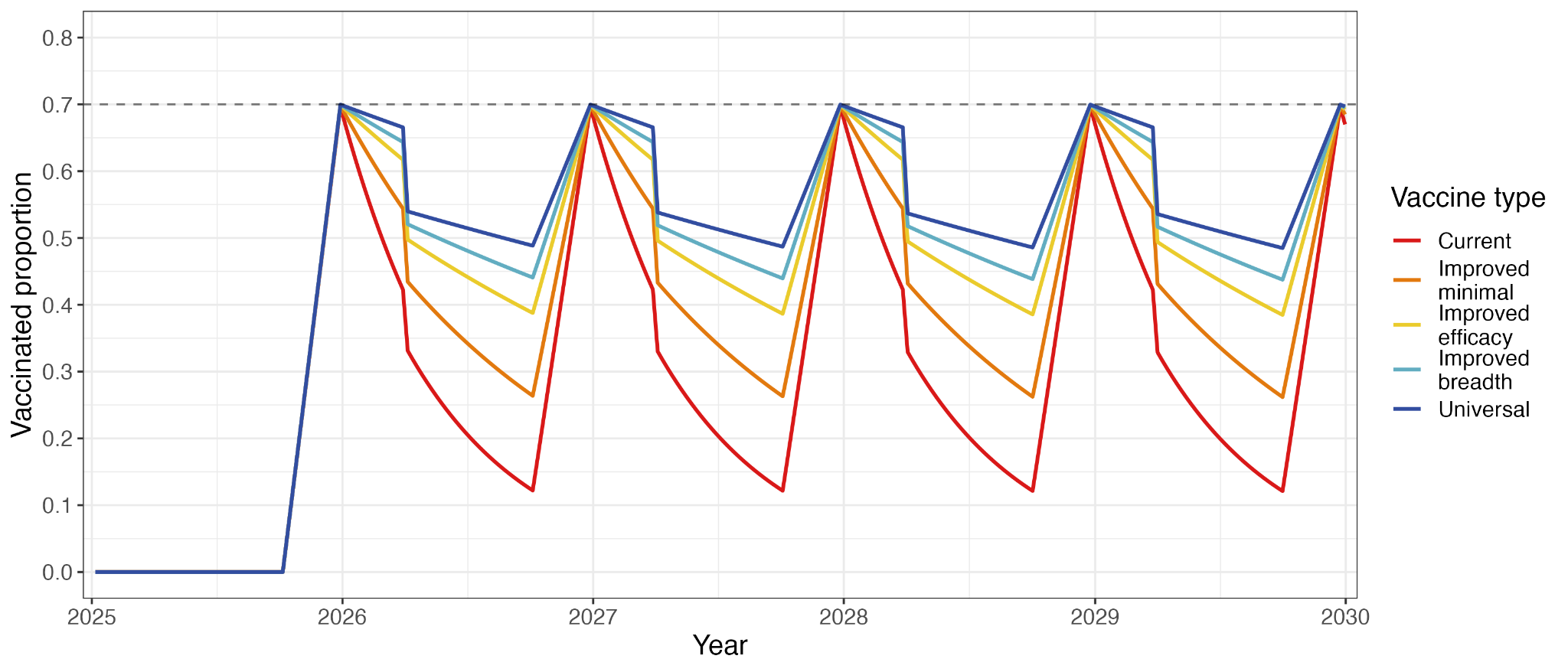

**Figure S21:** Example vaccination coverage in the 0-4 age group in a Northern Hemisphere country, under 70% vaccination coverage in the 0-4 age group.

#### b. Vaccine doses

As vaccine doses were distributed independently of previous vaccination and infection status, some vaccine doses were given to individuals who were still protected by previous vaccination, and were assumed to be ‘null’ as we did not assume any increased protection upon multiple doses (Figure S23).

| **Age-targeting strategy** | **Vaccine type** | **Vaccine doses (millions)** |
| --- | --- | --- |
| 0-4 | Current | 411 |
| 0-4 | Improved (minimal) | 329 |
| 0-4 | Improved (efficacy) | 255 |
| 0-4 | Improved (breadth) | 220 |
| 0-4 | Universal | 186 |
| 0-10 | Current | 888 |
| 0-10 | Improved (minimal) | 645 |
| 0-10 | Improved (efficacy) | 434 |
| 0-10 | Improved (breadth) | 339 |
| 0-10 | Universal | 249 |
| 0-17 | Current | 1550 |
| 0-17 | Improved (minimal) | 1150 |
| 0-17 | Improved (efficacy) | 795 |
| 0-17 | Improved (breadth) | 624 |
| 0-17 | Universal | 457 |
| 65+ | Current | 826 |
| 65+ | Improved (minimal) | 652 |
| 65+ | Improved (efficacy) | 476 |
| 65+ | Improved (breadth) | 387 |
| 65+ | Universal | 296 |
| 0-17, 65+ | Current | 2440 |
| 0-17, 65+ | Improved (minimal) | 1840 |
| 0-17, 65+ | Improved (efficacy) | 1290 |
| 0-17, 65+ | Improved (breadth) | 1020 |
| 0-17, 65+ | Universal | 755 |

**Table S6:** Global average annual vaccine doses given over the 30-year projection period under each age-targeting strategy and vaccine type, under 50% vaccination coverage.

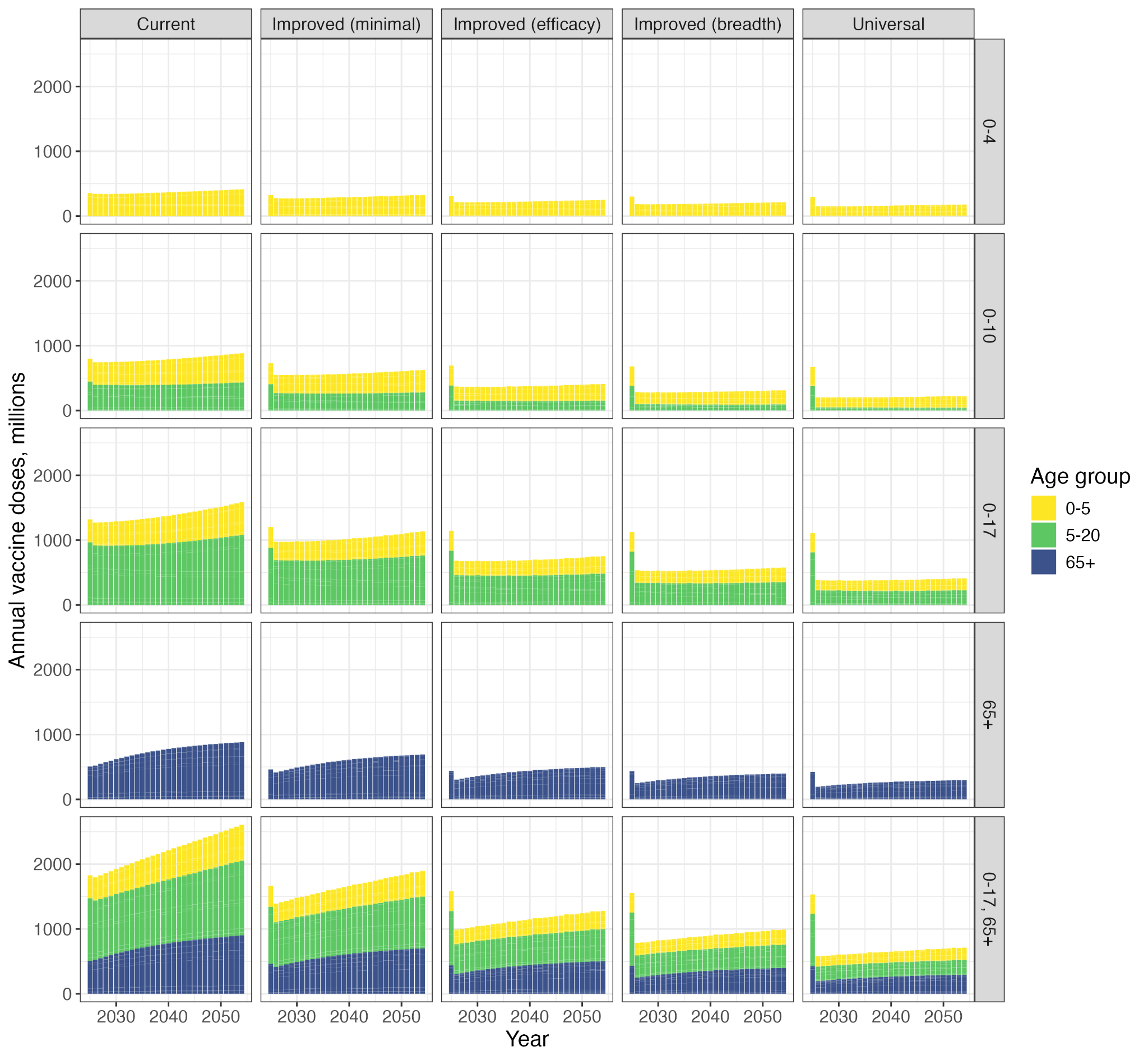

**Figure S22:** Annual age-specific vaccine doses given under each age-targeting strategy and vaccine type, assuming 50% vaccination coverage.

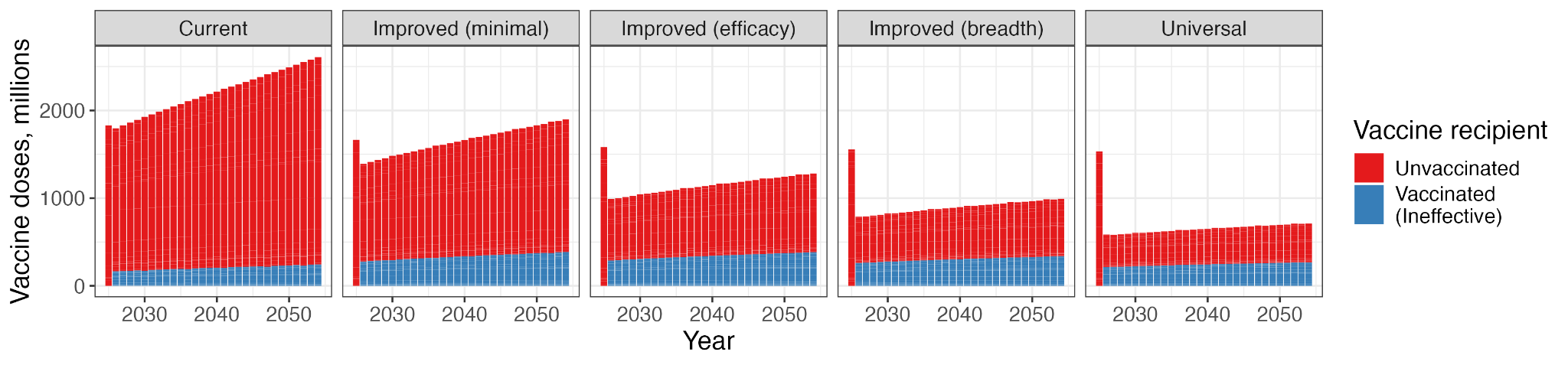

**Figure S23:** Annual vaccine doses given worldwide, stratified by vaccination status of the recipient, under 50% vaccination coverage of 0-17 and 65+ age groups.

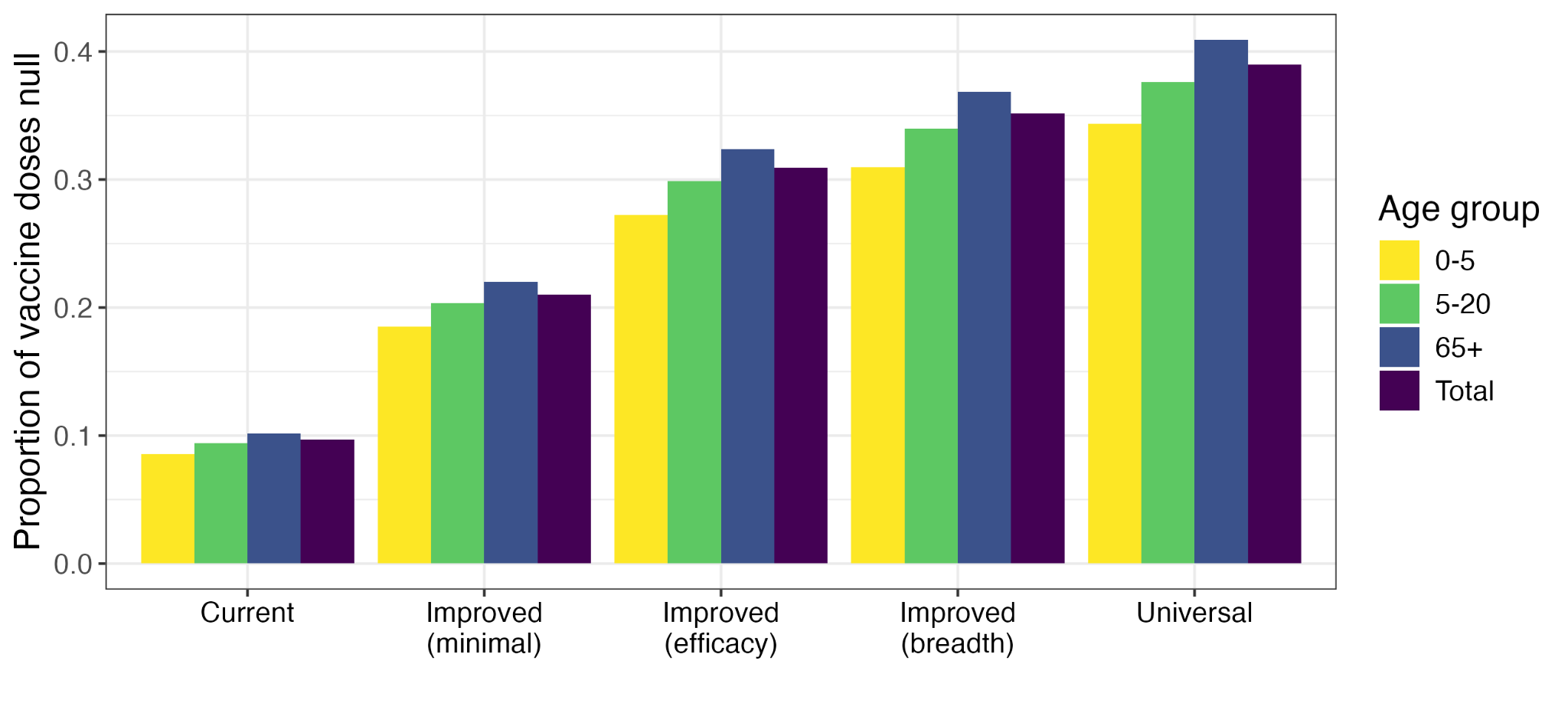

**Figure S24:** Proportion of annual age-specific vaccine doses given to already-vaccinated individuals (‘null’), assuming 50% vaccination coverage of 0-17 and 65+ age groups.

#### c. Influenza incidence

Figure S25 shows an overlay of ten simulated time series of influenza A and B infections in the exemplar countries under no vaccinations. We simulated 100 time series with randomly sampled epidemic years and parameters in each country, under each age-targeting strategy and vaccine type.

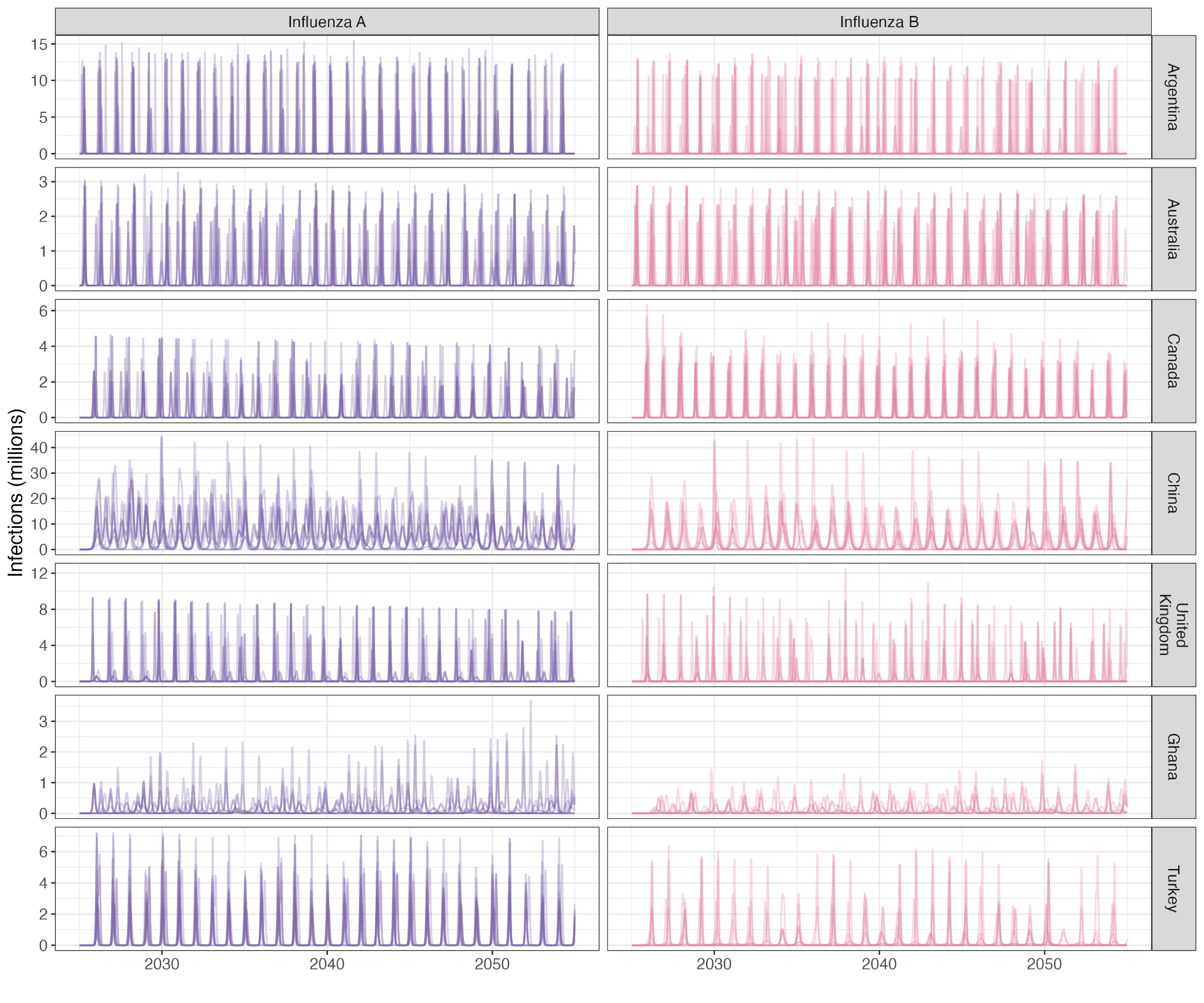

**Figure S25:** Overlay of ten simulations of influenza incidence in each exemplar country with no vaccination coverage, stratified by strain.

| **Age-targeting strategy** | **Vaccine type** | **Annual infections (millions)** |
| --- | --- | --- |
| No vaccination | | 3590 (3290, 3910) |
| 0-4 | Current | 3300 (3020, 3590) |
| 0-4 | Improved (minimal) | 3160 (2900, 3440) |
| 0-4 | Improved (efficacy) | 2710 (2490, 2960) |
| 0-4 | Improved (breadth) | 2610 (2400, 2860) |
| 0-4 | Universal | 1970 (1790, 2140) |
| 0-10 | Current | 2730 (2520, 3000) |
| 0-10 | Improved (minimal) | 2380 (2200, 2620) |
| 0-10 | Improved (efficacy) | 1610 (1480, 1800) |
| 0-10 | Improved (breadth) | 1660 (1520, 1830) |
| 0-10 | Universal | 1210 (1100, 1370) |
| 0-17 | Current | 2270 (2070, 2460) |
| 0-17 | Improved (minimal) | 1680 (1500, 1880) |
| 0-17 | Improved (efficacy) | 938 (847, 1080) |
| 0-17 | Improved (breadth) | 942 (857, 1070) |
| 0-17 | Universal | 614 (556, 681) |
| 65+ | Current | 3480 (3180, 3780) |
| 65+ | Improved (minimal) | 3450 (3160, 3750) |
| 65+ | Improved (efficacy) | 3350 (3070, 3650) |
| 65+ | Improved (breadth) | 3390 (3100, 3690) |
| 65+ | Universal | 3280 (2990, 3570) |
| 0-17, 65+ | Current | 2210 (2000, 2400) |
| 0-17, 65+ | Improved (minimal) | 1610 (1440, 1810) |
| 0-17, 65+ | Improved (efficacy) | 868 (787, 1000) |
| 0-17, 65+ | Improved (breadth) | 877 (798, 998) |
| 0-17, 65+ | Universal | 545 (492, 604) |

**Table S7:** Annual influenza infections under each vaccine type and age-targeting strategy, assuming 50% vaccination coverage (median, 95% uncertainty intervals).

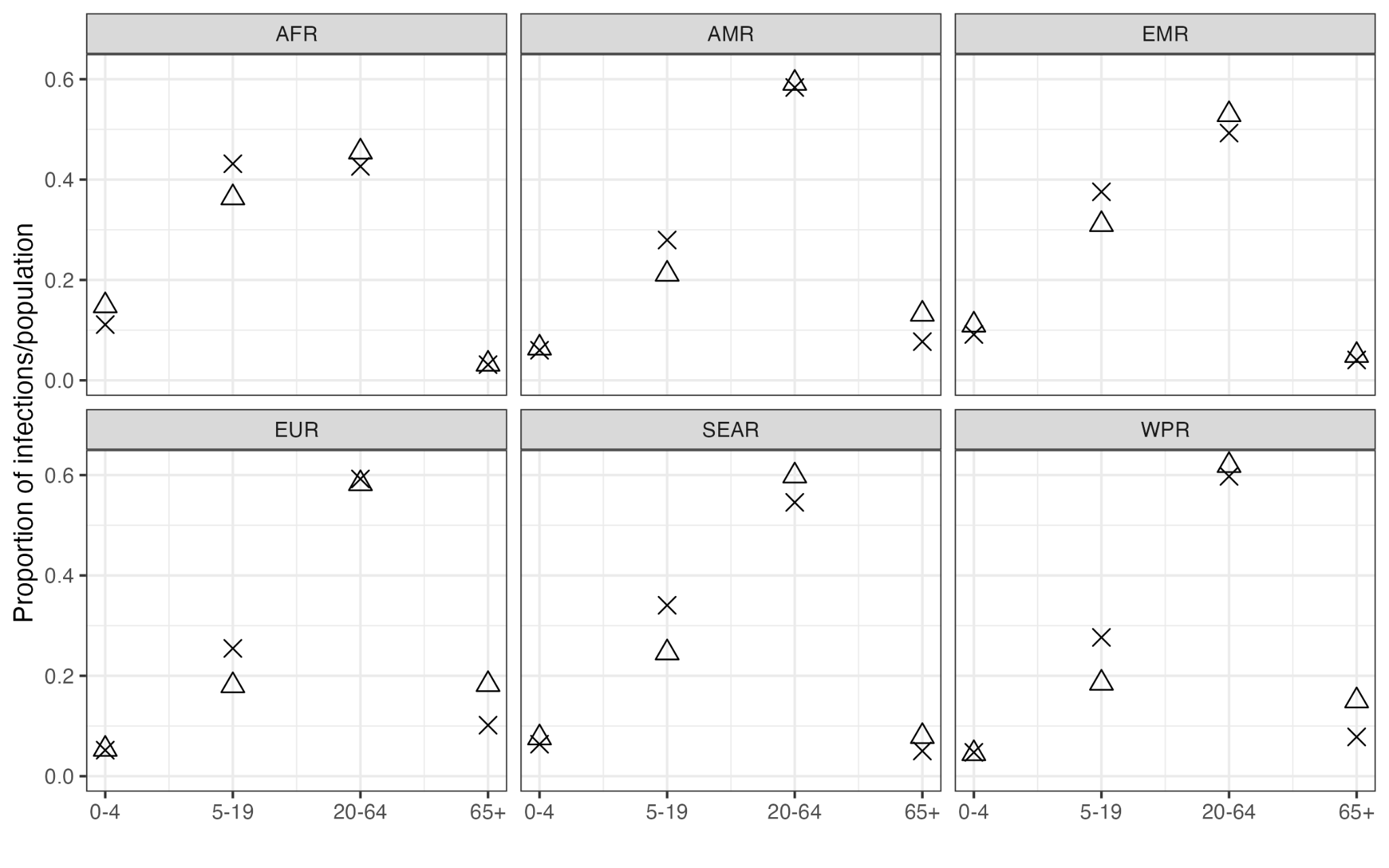

**Figure S26:** Median distribution of influenza infections across age groups under no vaccinations in each WHO region (shown as crosses), compared to distribution of the 2025 population (shown as triangles).

#### d. Infections averted

| **Age-targeting strategy** | **Vaccine type** | **Annual infections averted (millions)** | **Median proportion of infections averted** |
| --- | --- | --- | --- |
| 0-4 | Current | 292 (261, 321) | 8% |
| 0-4 | Improved (minimal) | 436 (390, 482) | 12% |
| 0-4 | Improved (efficacy) | 881 (803, 970) | 25% |
| 0-4 | Improved (breadth) | 972 (897, 1070) | 27% |
| 0-4 | Universal | 1630 (1490, 1800) | 45% |
| 0-10 | Current | 849 (771, 941) | 24% |
| 0-10 | Improved (minimal) | 1200 (1100, 1330) | 33% |
| 0-10 | Improved (efficacy) | 1990 (1790, 2170) | 55% |
| 0-10 | Improved (breadth) | 1930 (1760, 2130) | 54% |
| 0-10 | Universal | 2360 (2150, 2600) | 66% |
| 0-17 | Current | 1330 (1200, 1480) | 37% |
| 0-17 | Improved (minimal) | 1930 (1720, 2110) | 53% |
| 0-17 | Improved (efficacy) | 2650 (2390, 2930) | 74% |
| 0-17 | Improved (breadth) | 2640 (2390, 2930) | 74% |
| 0-17 | Universal | 2960 (2700, 3270) | 83% |
| 65+ | Current | 117 (105, 129) | 3% |
| 65+ | Improved (minimal) | 144 (129, 159) | 4% |
| 65+ | Improved (efficacy) | 241 (215, 266) | 7% |
| 65+ | Improved (breadth) | 203 (188, 223) | 6% |
| 65+ | Universal | 315 (292, 346) | 9% |
| 0-17, 65+ | Current | 1400 (1260, 1550) | 39% |
| 0-17, 65+ | Improved (minimal) | 2000 (1790, 2180) | 55% |
| 0-17, 65+ | Improved (efficacy) | 2720 (2450, 3020) | 76% |
| 0-17, 65+ | Improved (breadth) | 2700 (2450, 3000) | 75% |
| 0-17, 65+ | Universal | 3030 (2770, 3350) | 85% |

**Table S8:** Annual influenza infections averted under each vaccine type and age-targeting strategy, assuming 50% vaccination coverage (median, 95% uncertainty intervals), and median percentage of influenza infections averted, compared to under no vaccinations.

#### e. Number needed to vaccinate (NNV)

We calculated the number needed to vaccinate (NNV) in order to prevent one influenza infection under each age-targeting strategy and vaccine type, including wasted vaccines. Figure S27 shows the distributions of median NNV within each WHO region.

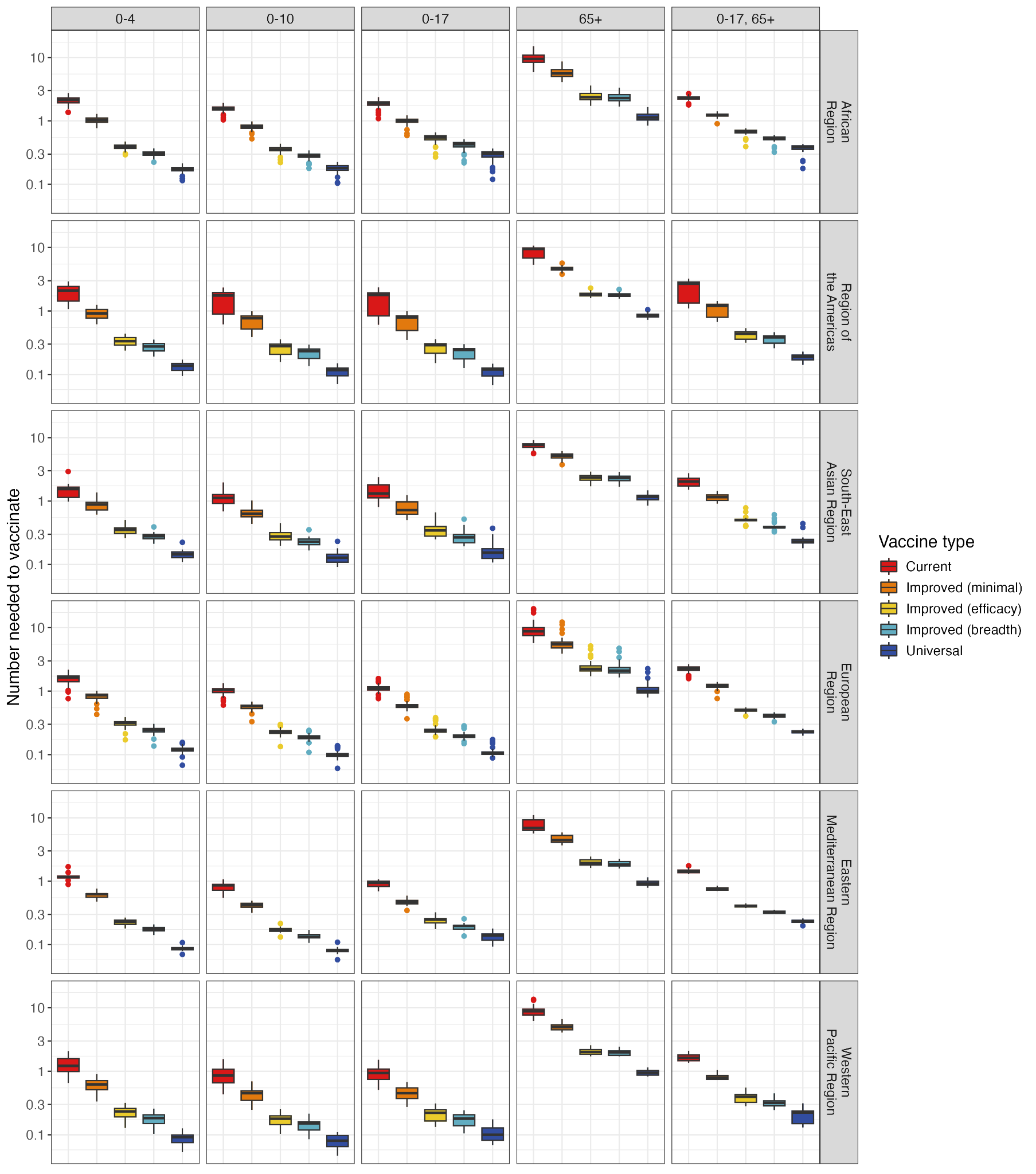

**Figure S27:** Number needed to vaccinate, stratified by WHO region, under each age-targeting strategy and vaccine type.

#### f. Scaling of contact matrices

We scaled the synthetic contact matrices in each epidemic to match the R_0_ value of the original epidemic in the exemplar country. As a sensitivity analysis, we also conducted the epidemic projections without scaling to R_0_. In this analysis, there were slightly more influenza infections over the 30-year period (for example 3600-4300 annual infections under no vaccinations, compared to 3300-3900 when scaling contact matrices to match R_0_), but the relative effects of improved vaccines remained similar.

However, in some countries the synthetic contact matrices were not of a magnitude to produce influenza epidemics, even in the ‘no vaccination’ scenario, leading to minimal influenza infections over the 30-year period. The countries with minimal projected influenza infections reported annual influenza epidemics in FluNet. This phenomenon in this sensitivity analysis therefore justified the decision to scale contact matrices to match original R_0_ values.

### 7. Economic modelling

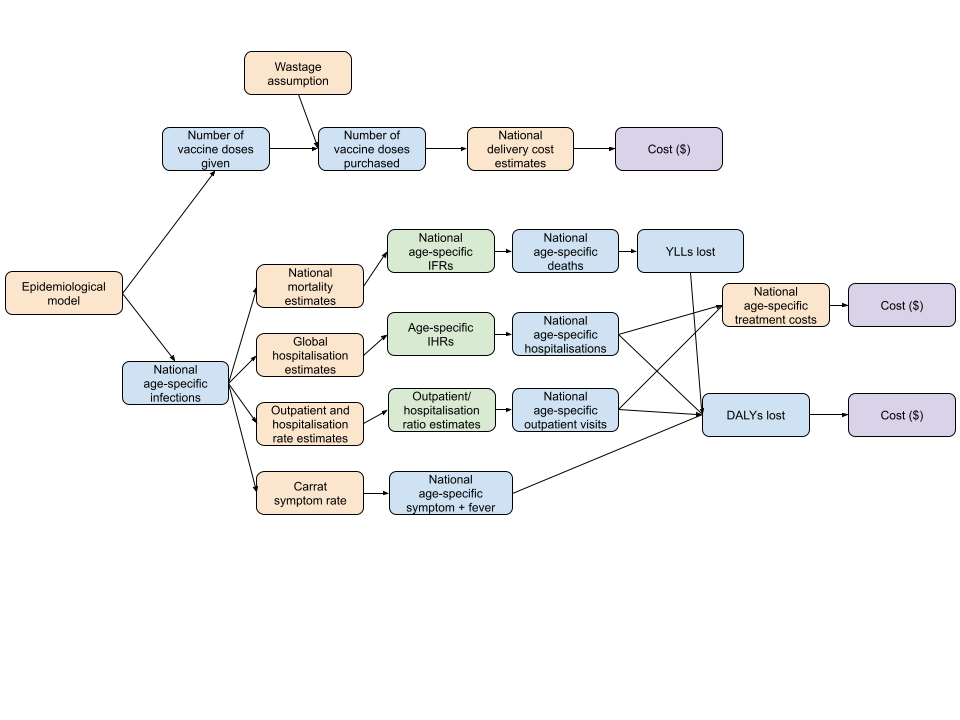

**Figure S28:** Overview of the economic decision tree model.

#### a. Health outcomes data

Non-death DALYs incurred were calculated using globally-consistent probabilities and weights for symptomatic (non-fever) infections, symptomatic (fever) infections, and hospitalisations (Tables S9, S10, S11).

##### Symptomatic and fever probabilities

We used probabilities of symptomatic influenza and fever upon infection from the literature [49]. We assumed that these values are consistent regionally and at all ages, and sampled from their distribution. We then calculated the number of symptomatic non-fever infections.

| **Outcome** | **Probability of occurrence** | **95% uncertainty interval** |
| --- | --- | --- |
| **Symptoms** | 0.669 | (0.583, 0.745) |
| **Fever** | 0.349 | (0.267, 0.442) |

**Table S9:** Probabilities of symptomatic influenza and fever upon infection.

##### Infection-fatality ratios

National age-specific infection fatality ratios (IFRs) were calculated using a combination of our assigned ITZs, estimated age-specific national attack rates in 2010-2015 from the above analysis, estimates of national age-specific seasonal influenza-associated respiratory mortality [50] (up to 2015), and national 2015 population age structures and all-cause mortality rates from World Population Prospects 2022 [48].

We applied three strategies for extrapolating IFRs, depending on current seasonal influenza vaccination coverage and the representativeness of exemplar countries’ mortality rates.

In ITZs where current seasonal vaccination coverage is low/near-zero (Africa, Asia-Europe, Southern and Eastern Asia), we calculated each country’s age-specific IFRs as follows:

1. Fit a gamma distribution to each age-specific mortality rate (given as a median and 95% uncertainty interval in [50]), which are stratified into <65, 65-74 and 75+ age groups
2. Randomly sample 100 mortality rates from each distribution
3. Obtain mortality rate samples for 65+ age group by weighting the 65-74 and 75+ samples by the 2015 sizes of each age group
4. Run 100 simulations of the estimated 2010-2015 age-specific influenza burden in the target country using the inferred timing and transmission parameters of the ITZ’s exemplar country, but the demography of the target country
5. Calculate 100 samples of mean 2010-2015 age-specific attack rates in the target country using 2015 population
6. For each of the 100 samples of 65+ mortality rate and 65+ attack rate, calculate the 65+ IFR as (mortality rate/attack rate)
7. Calculate the all-cause mortality rates in 0-4, 5-19, 20-64 age groups using the target country’s UN 2015 life table and the population age distribution
8. For each of the 100 samples of age-specific attack rates and influenza mortality rates, calculate the three age-specific IFRs by scaling down the all-cause mortality rates such that when applied to the target country’s 2015 population and attack rates, the corresponding influenza mortality is achieved

In ITZs where there is significant seasonal influenza coverage, we calculated the age-specific IFRs as above in the exemplar country only, and then applied these IFRs to each country in the ITZ due to their flu transmission similarities.

In Southern America, where the exemplar country of Argentina was found to have atypically high seasonal influenza mortality rates [50], but significant vaccination coverage, we applied the above analysis to both Argentina and Brazil, as Brazil was found to have more representative influenza mortality rates for the rest of the ITZ. We used age-specific 2010-2015 vaccination coverage in Brazil as reported in [51]. We then applied Argentina’s age-specific IFRs to itself only, and Brazil’s to the rest of the Southern America ITZ. In this way, we have attempted to obtain the most representative global IFRs for our analysis.

We sampled from the obtained distribution for each country/ITZ and age group.

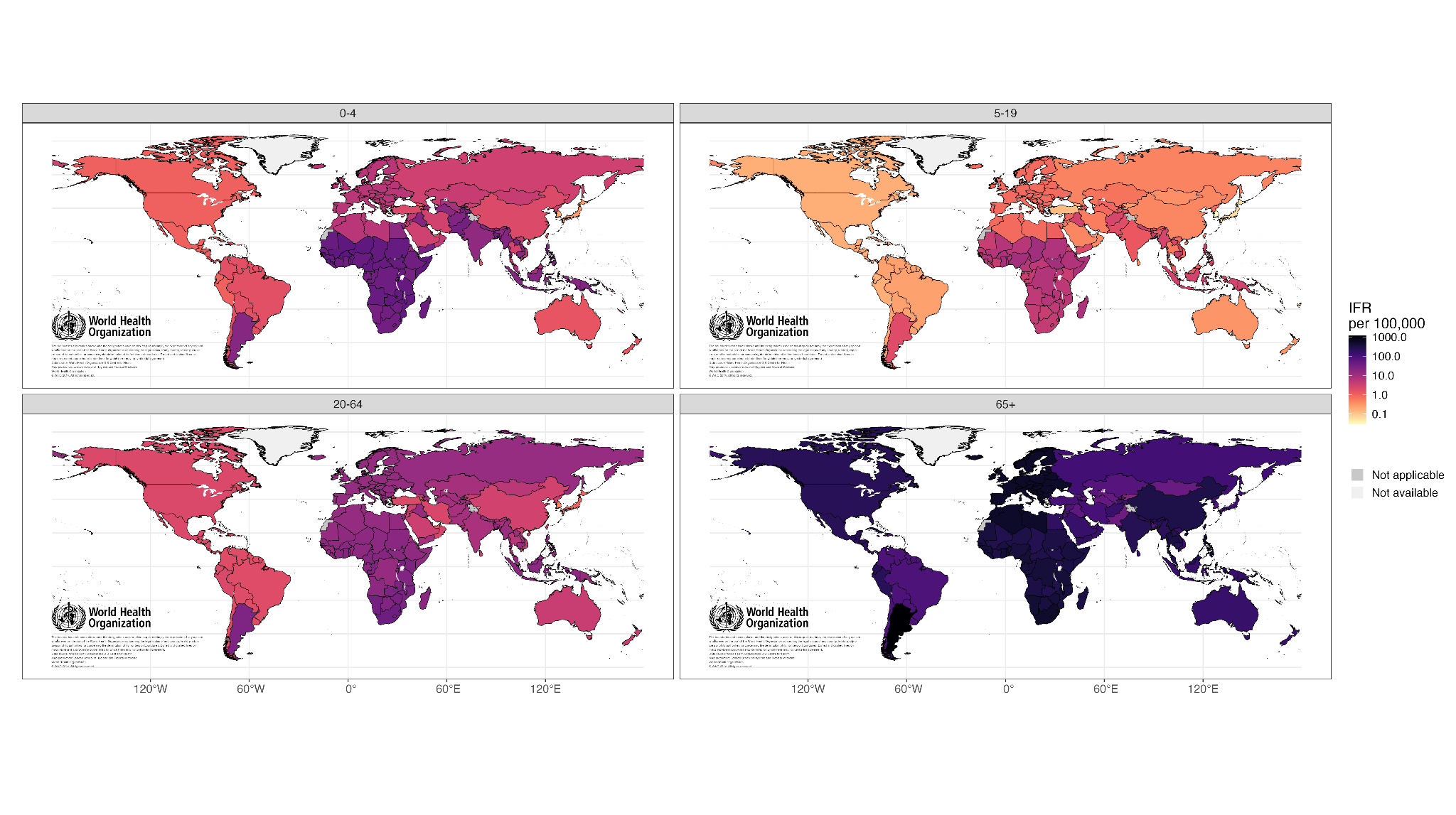

**Figure S29:** Age-specific national IFRs, per 100,000 infections. Map base layer from <https://gis-who.hub.arcgis.com/pages/detailedboundary>.

##### Infection-hospitalisation ratios

To determine age-specific infection hospitality ratios (IHRs), we combined our distribution of the global 2010-2015 age-specific attack rates as calculated above and samples of global age-specific hospitalisation rates as reported in Paget et al. [52]. We weighted the IHRs for the 5-19 and 20-64 age groups according to the age distribution of hospitalisation rates found by Cromer et al. [53]. The resulting age-specific IHRs are shown in Table S10. We sampled from their distribution.

| **Age group** | **Median IHR per 100,000 infections** | **95% Uncertainty interval** |
| --- | --- | --- |
| **0-4** | 855.29 | (436.25, 1434.13) |
| **5-19** | 31.55 | (22.91, 40.32) |
| **20-64** | 43.04 | (31.26, 55.02) |
| **65+** | 572.02 | (328.40, 884.16) |

**Table S10:** Calculated age-specific infection hospitalisation ratios.

##### Outpatient visit rates

We did not account for the cost of outpatient visits in our base analysis, due to a lack of data on their frequency and drastically varying characteristics of an outpatient visit depending on setting, but included them as a sensitivity analysis. We sampled from estimates of hospitalisation and outpatient visit incidence using data from the Global Burden of Disease (GBD) Study 2017 [49], and calculated the estimated ratio between hospitalisation and outpatient visit incidence in each GBD super-region. We applied these ratios to the hospitalisation estimates in each vaccination scenario to obtain estimated outpatient visits.

##### Disability-adjusted life years

YLLs were discounted at 3% in the base case and 0% in a sensitivity analysis, and calculated using life tables from the World Population Prospects 2022 [48]. We used influenza disability weights as reported by the Global Burden of Disease study (GBD) [54]:

| **Outcome** | **Median** | **95% Uncertainty Interval** |
| --- | --- | --- |
| **Non-fever** | 0.006 | (0.002, 0.012) |
| **Fever** | 0.051 | (0.032, 0.074) |
| **Hospitalisation** | 0.133 | (0.088, 0.190) |

**Table S11:** Influenza disability weights for each health outcome [54].

We sampled from the distribution of each disability weight and applied these to our estimates of the frequency of each outcome. We assumed that cases of non-fever, fever, and hospitalisation stays each had a duration of 4 days at all ages [49,55].

#### b. Economic inputs

##### Cost of hospitalisation/outpatient

To estimate the national age-specific costs of hospitalisation and outpatient care, we took estimates from existing systematic reviews and meta-analyses and used GDP per capita as a predictor for costs [56–60]. Cost estimates from the literature were split into the broad study populations of children, adults, and elderly, which we then applied to our model age groups of [0,5), [5-64), and [65+) respectively. We supplemented the sparse data on the cost of adult outpatient care with a further search of the literature [55,61,62].

We used GDP per capita, inflation rates, and local currency unit exchange rates from World Bank data. Costs were predicted using a joint log-linear model with interactions between outcomes (hospitalisation, outpatient) and study populations (children, adults, elderly):

*lm(log(cost_usd_2022) ~ log(log(gdp_per_capita))*factor(outcome)*factor(study_pop)*

In the case that we had multiple estimates of costs for a given study population and outcome within the same country (n_obs_ such that n_obs_ > 1), we weighted each observation by 1/sqrt(n_obs_), so as to not over-rely on countries with more cost estimates.

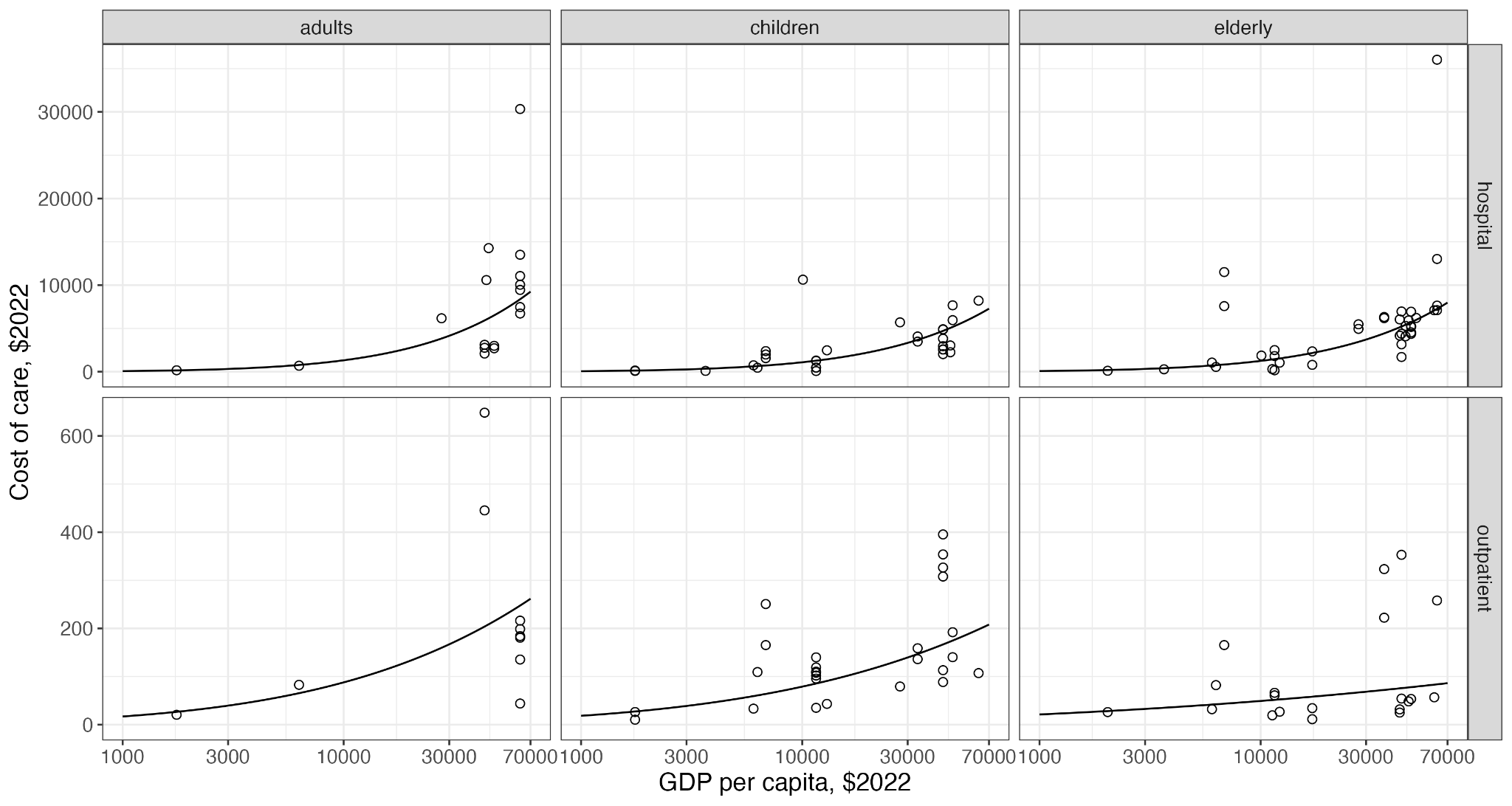

**Figure S30:** Mean estimated costs of care for adult, children, and elderly hospitalisations and outpatient visits, with GDP per capita shown on a log scale. GDP per capita and costs of care in 2022 USD. Data points shown are estimates from the literature.

We then sampled costs for each country and age group from the normal distribution of the log-linear model, to capture the uncertainty of the model.

##### Willingness-to-pay

We used national willingness to pay thresholds estimated in Pichon-Riviere et al. [63]. These were reported as a proportion of GDP per capita; we recalculated the WTP thresholds using 2022 GDP per capita for each country. For countries without thresholds in [63], we estimated the proportion of GDP per capita used as a willingness to pay threshold using a log-linear model regressing on GDP per capita. We also conducted a sensitivity analysis using 50% of 2022 GDP per capita in each country. Figure S31 shows the comparison between the two WTP thresholds for each country, colour-coded by World Bank income group. Low-income and lower-middle income countries tended to have WTP thresholds lower than 50% of GDP per capita, while high-income countries generally had WTP thresholds higher than 50% of GDP per capita.

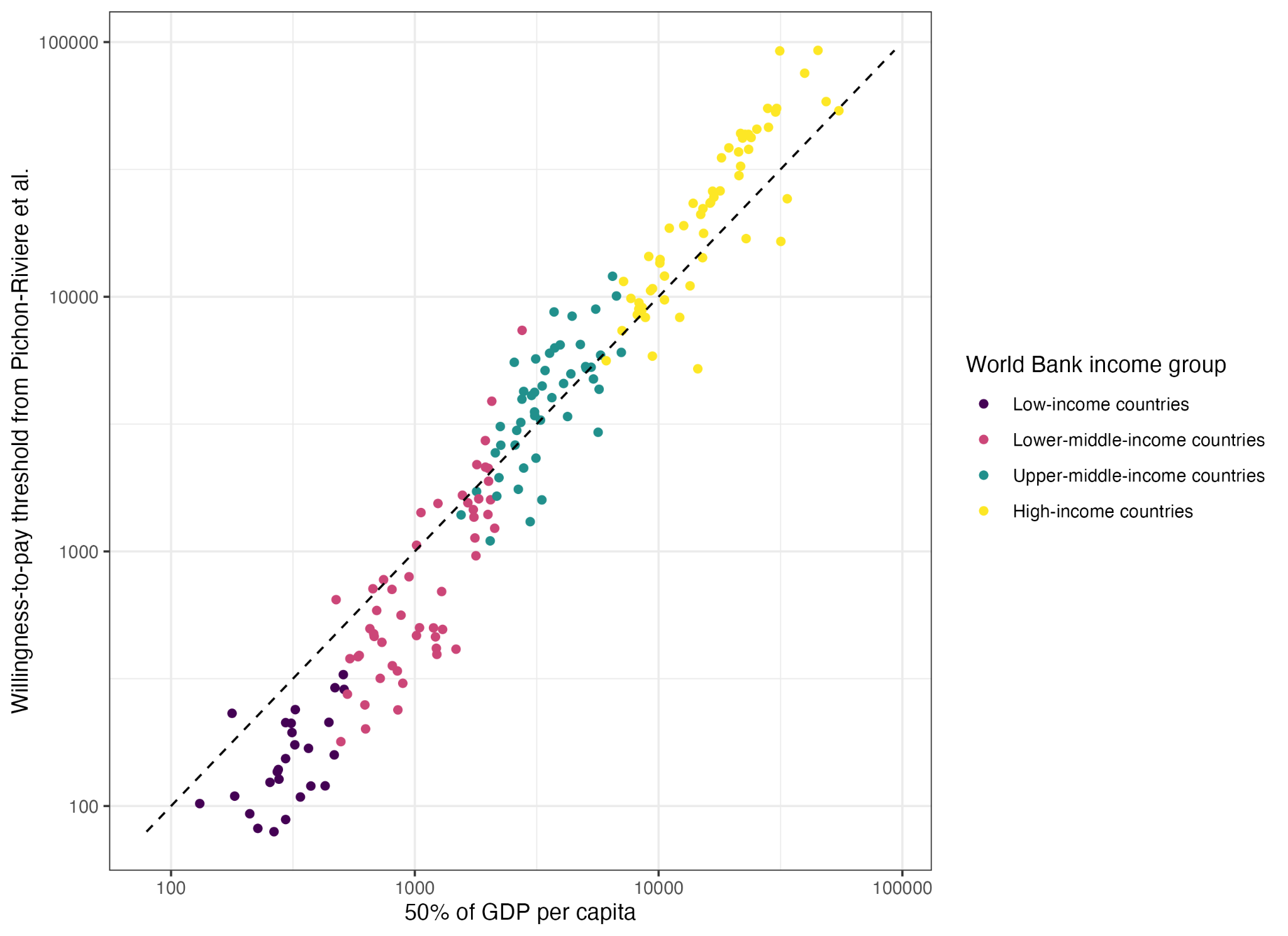

**Figure S31**: National willingness-to-pay thresholds [63] and 50% of 2022 GDP per capita. Dotted line indicates y=x.

##### Vaccine delivery cost

We estimated the country-level costs of vaccine dose delivery using data from Portnoy et al. [64], which used Bayesian meta-regression to predict delivery costs against country- and study-level factors. The model was informed by data from 29 studies, and produced estimates for 129 countries used in this study. To extrapolate to HICs, we included additional estimates of vaccine delivery cost from the USA, United Kingdom, and Spain [65–67]. Figure S32 shows the original and additional data, against healthcare expenditure per capita.

Where cost estimates were available from the Portnoy et al. paper, we sampled 100 delivery costs from a log-normal distribution using the median cost and uncertainty intervals provided. For those countries not included in the existing dataset, we predicted vaccine dose delivery costs using a linear regression against healthcare expenditure per capita, and sampled delivery costs from the associated prediction intervals, assuming a normal distribution. The regression model weighted the cost estimates from Portnoy et al. with weight 29/129, and the additional data with weight 1, so that each study informing the model was represented with weight 1.

When calculating the related costs of vaccination programs, we also assumed a constant rate of 10% vaccine wastage (the proportion of procured vaccine doses which were not administered).

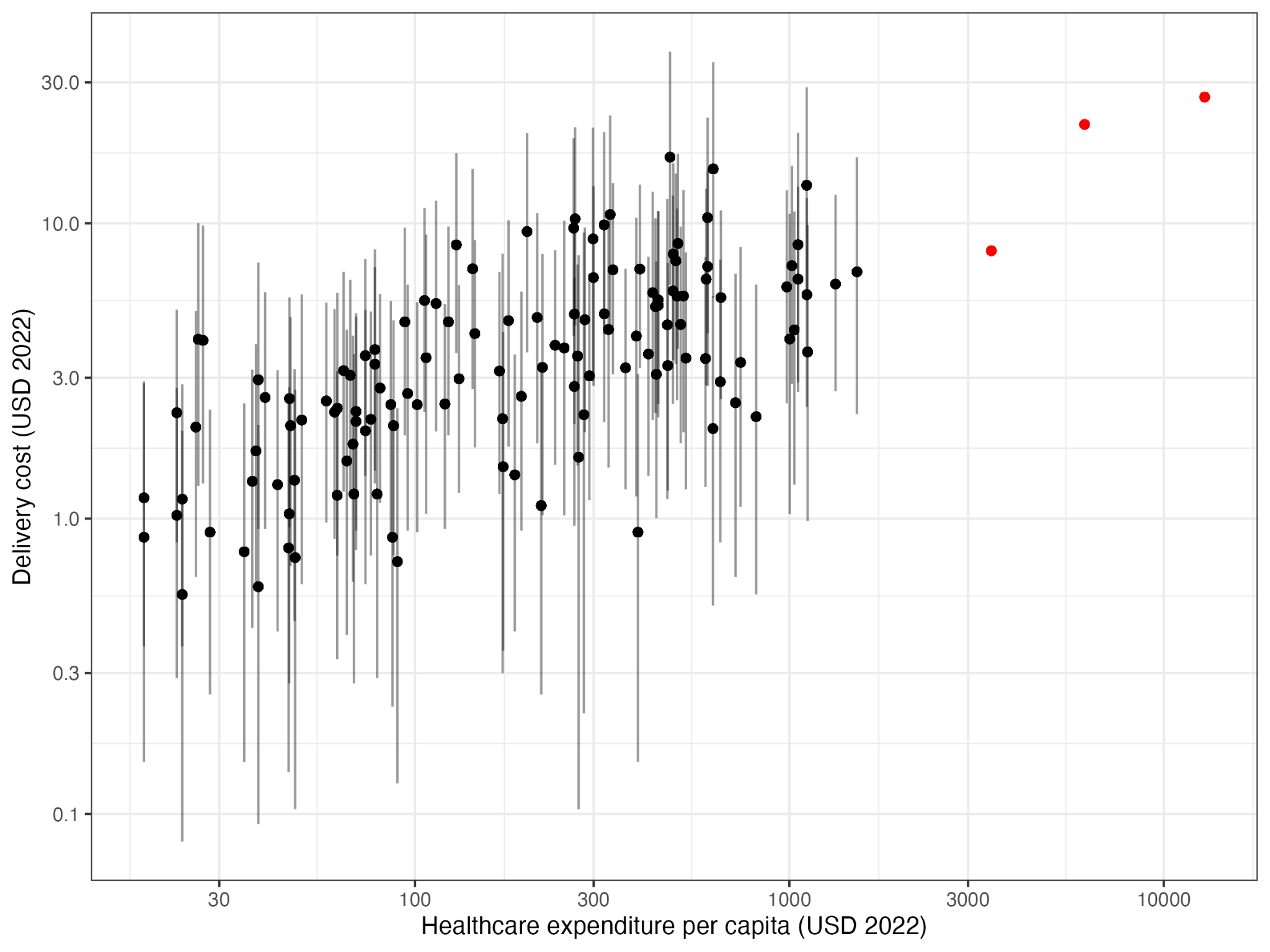

**Figure S32:** Costs of vaccine dose delivery in LMICs from Portnoy et al. [64] with 95% uncertainty intervals (black), and additional HIC data for regression (red), against healthcare expenditure per capita, on a log-log scale.

#### c.  Health outcomes

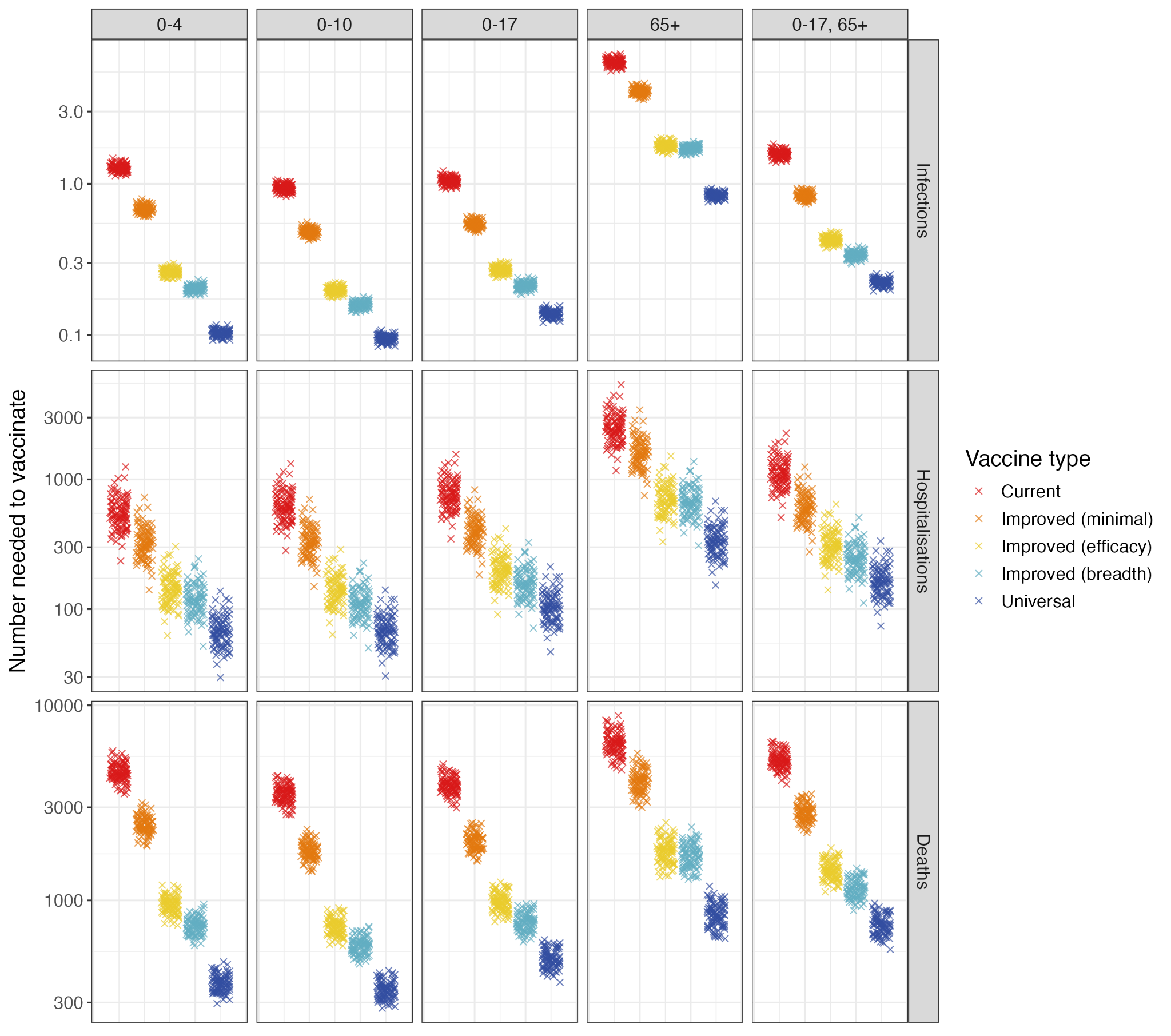

**Figure S33:** Global number needed to vaccinate to prevent one influenza-associated infection, hospitalisation, or death, for each vaccine type and age-targeting strategy, on a log scale.

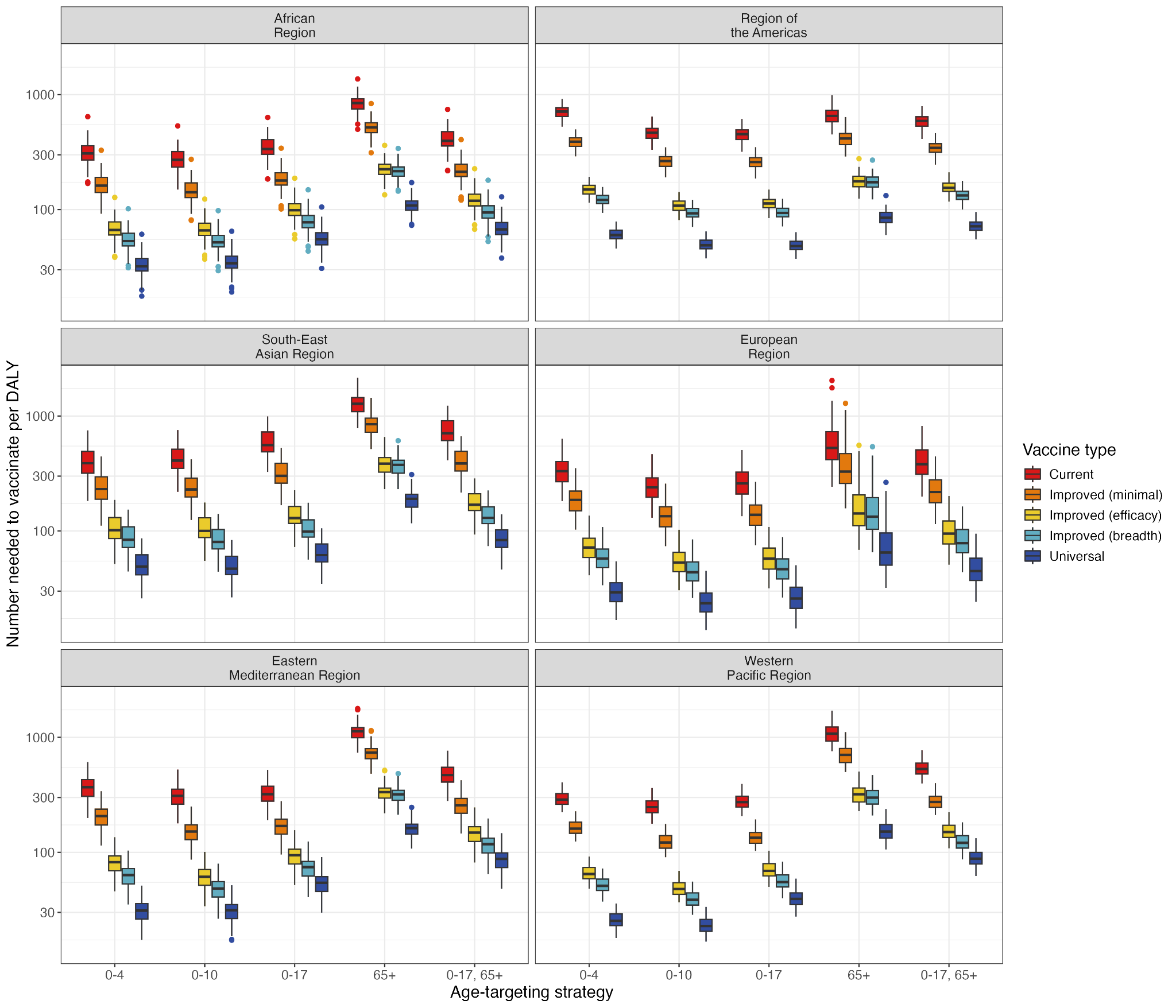

**Figure S34:** Number needed to vaccinate to avert one DALY in each WHO region, for each vaccine type and age-targeting strategy, on a log scale.

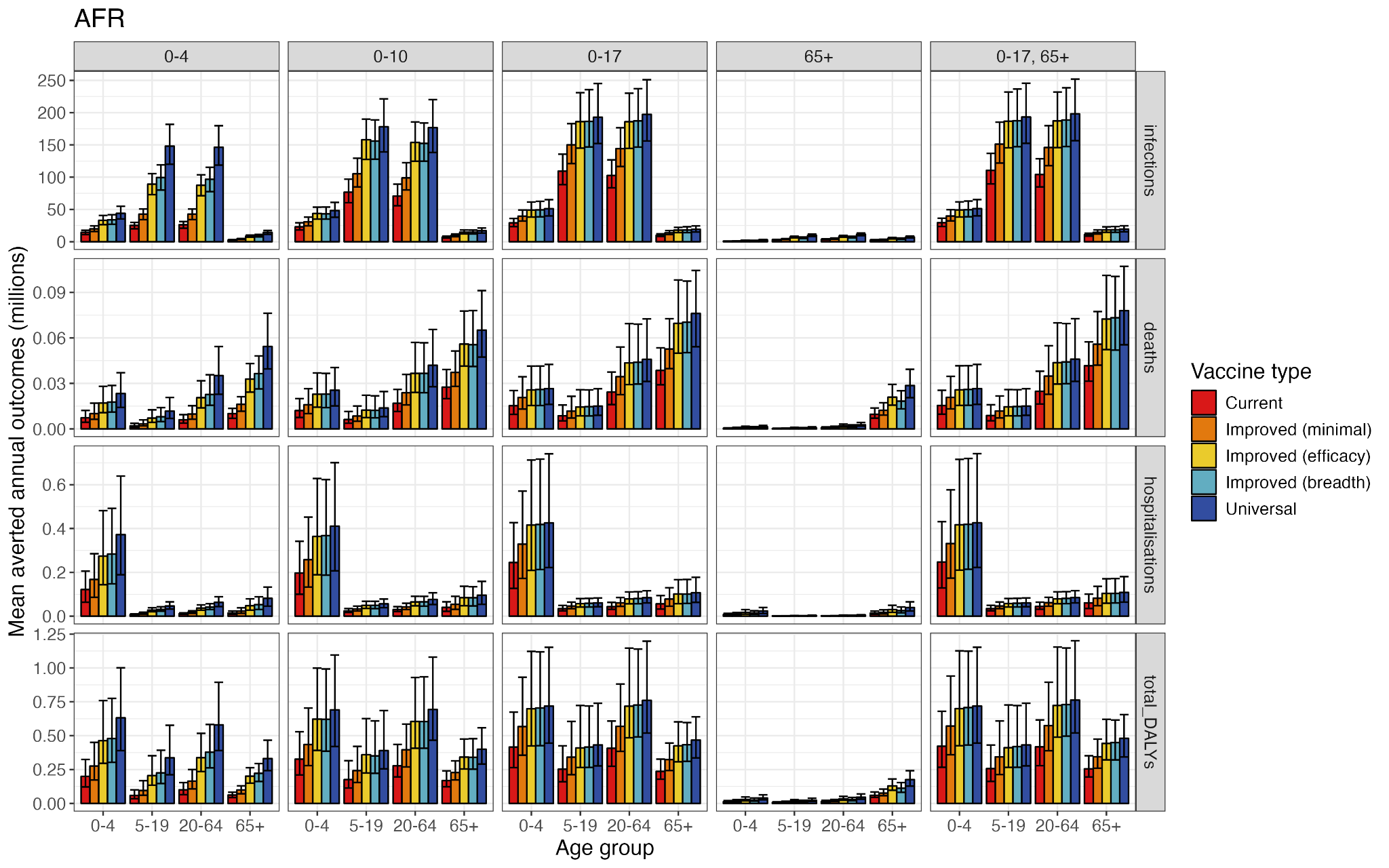

**Figure S35:** Averted annual age-specific health outcomes under each age-targeting strategy and vaccine type, in the African Region.

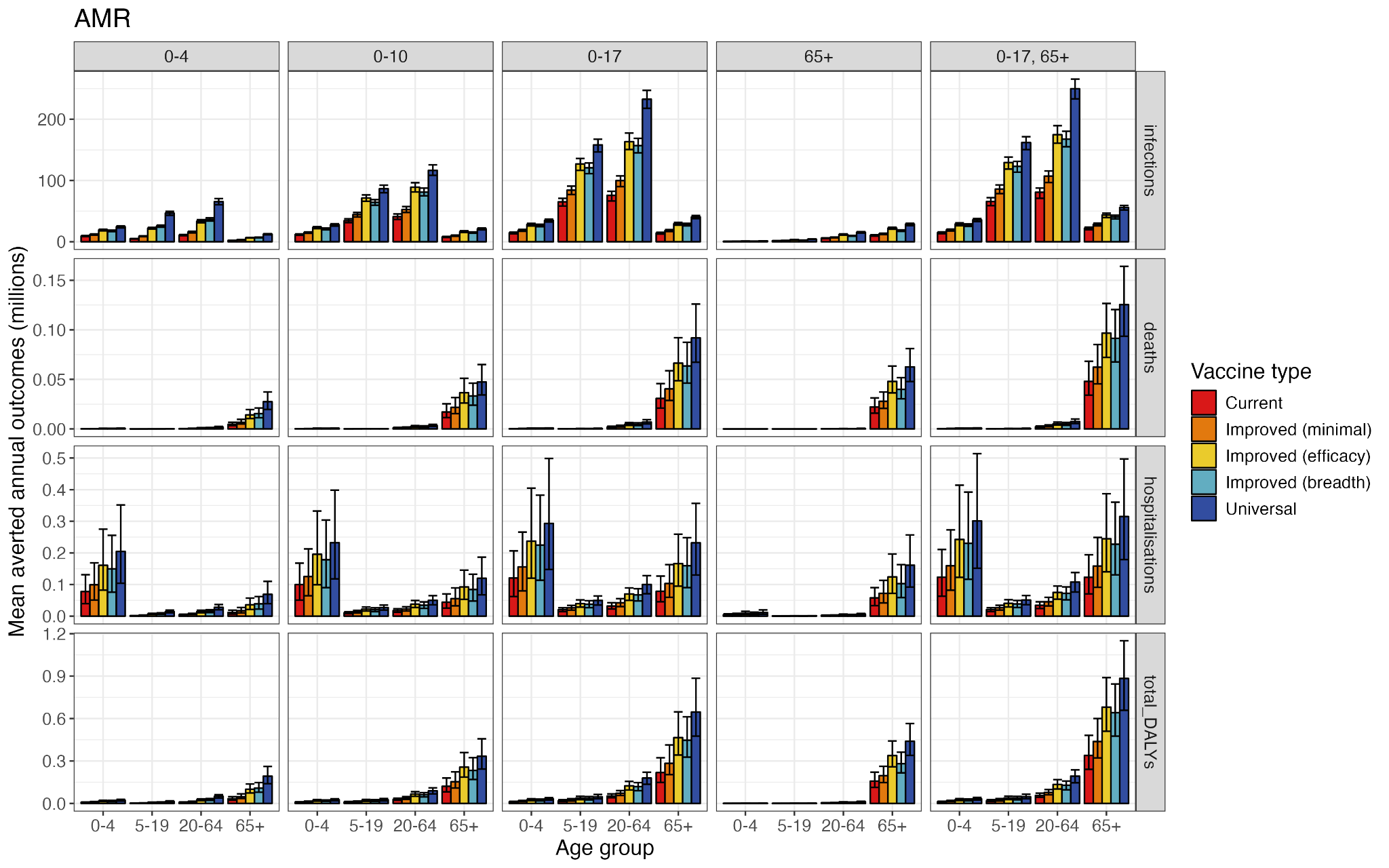

**Figure S36:** Averted annual age-specific health outcomes under each age-targeting strategy and vaccine type, in the Region of the Americas.

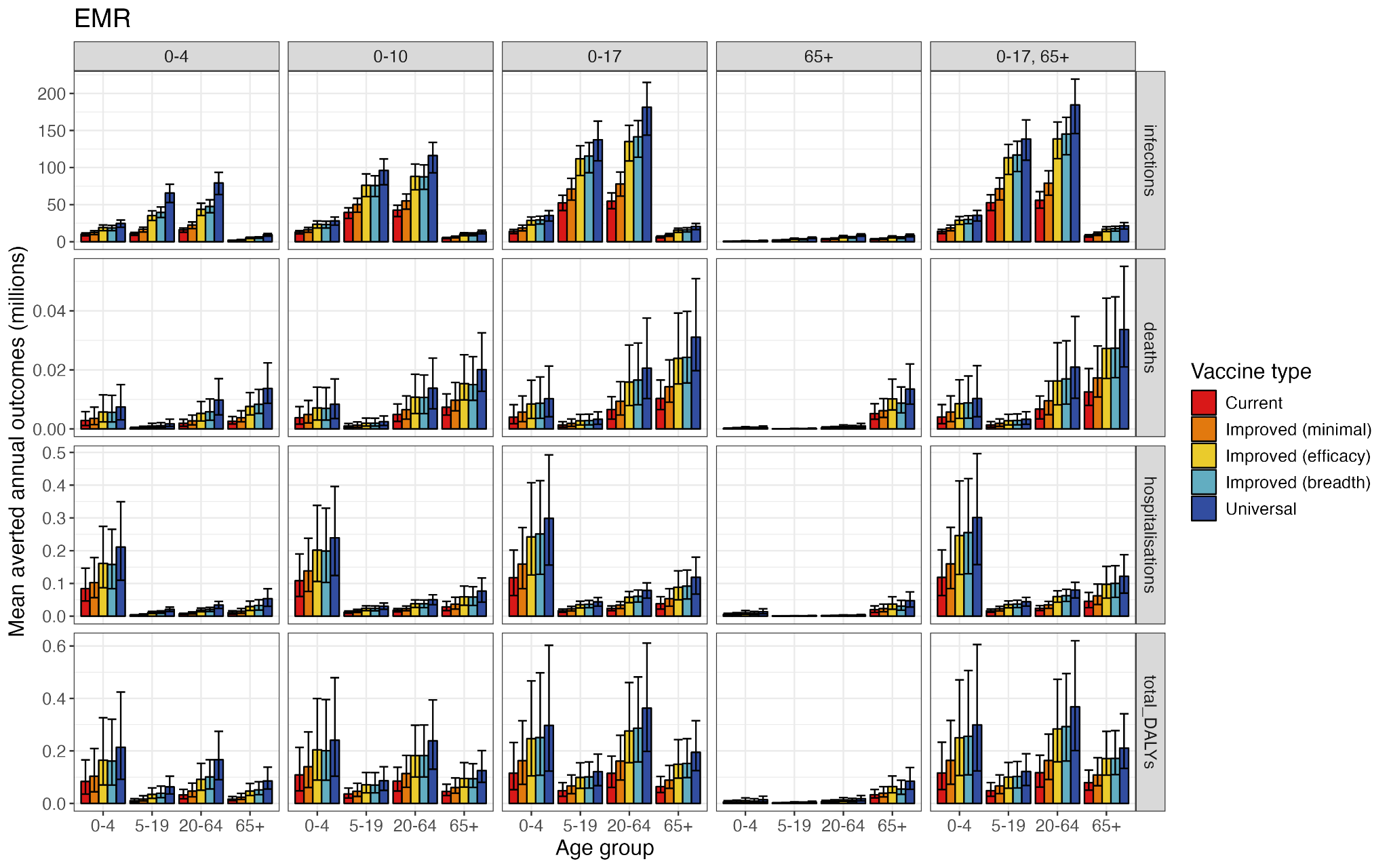

**Figure S37:** Averted annual age-specific health outcomes under each age-targeting strategy and vaccine type, in the Eastern Mediterranean Region.

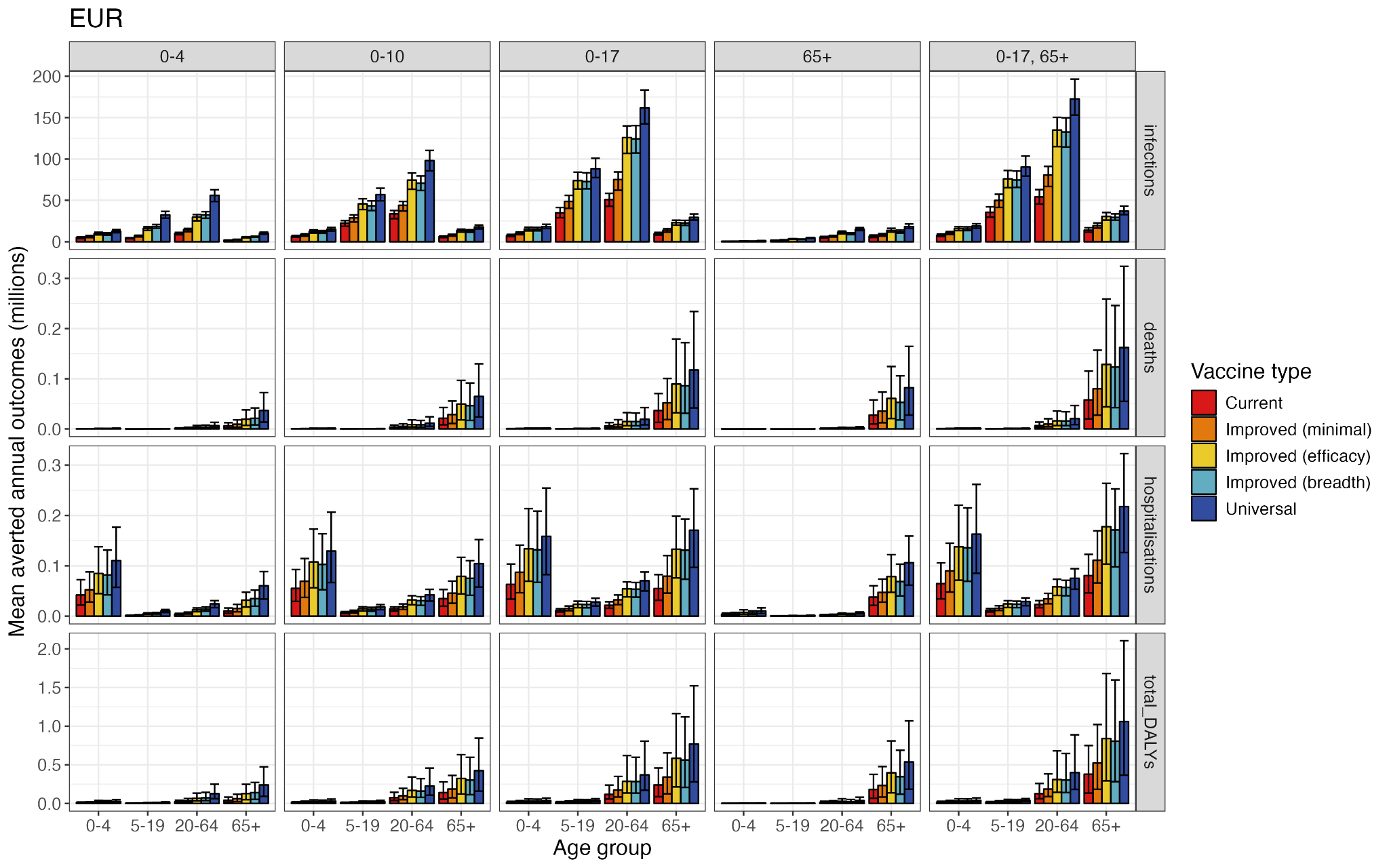

**Figure S38:** Averted annual age-specific health outcomes under each age-targeting strategy and vaccine type, in the European Region.

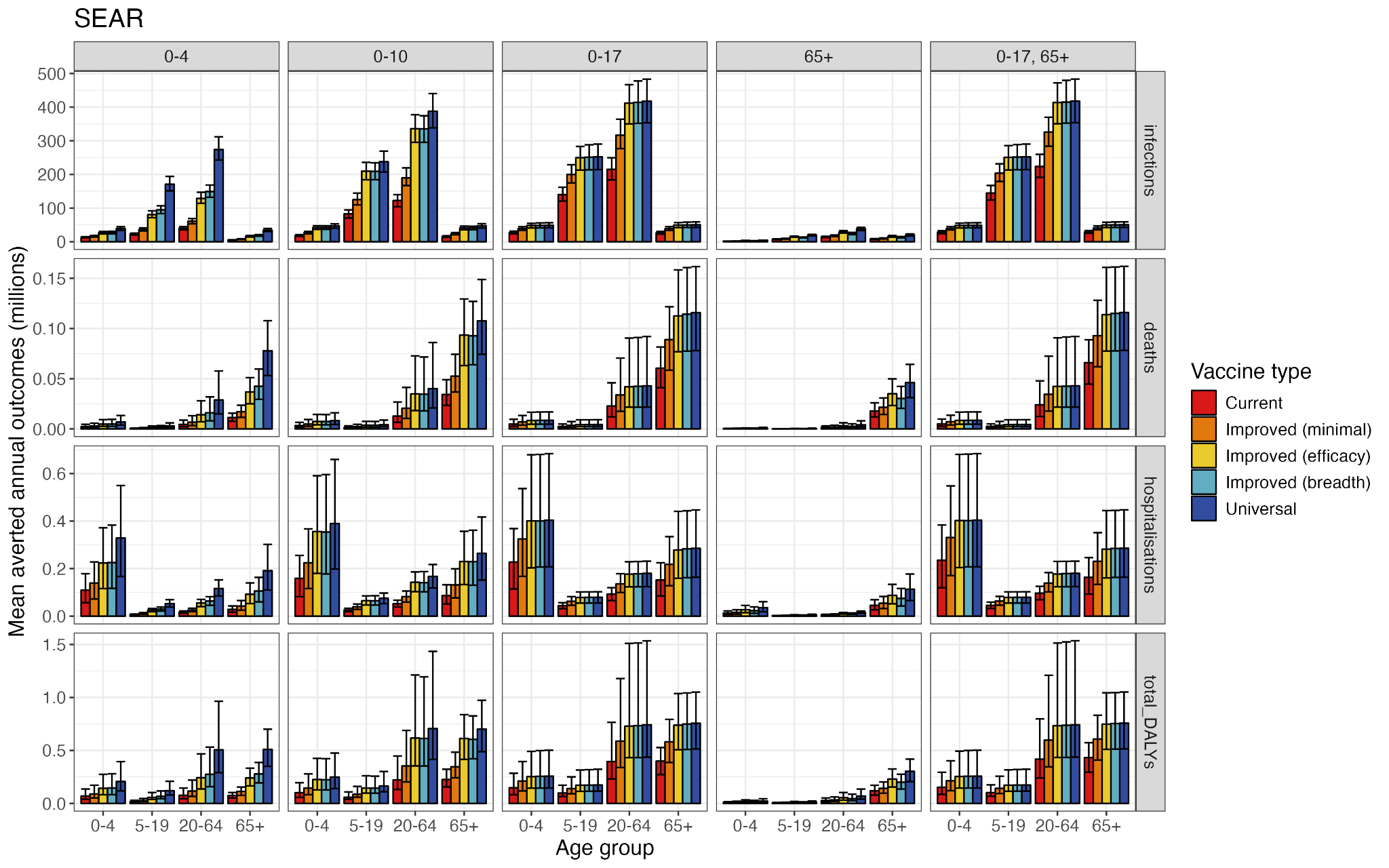

**Figure S39:** Averted annual age-specific health outcomes under each age-targeting strategy and vaccine type, in the South-East Asian Region.

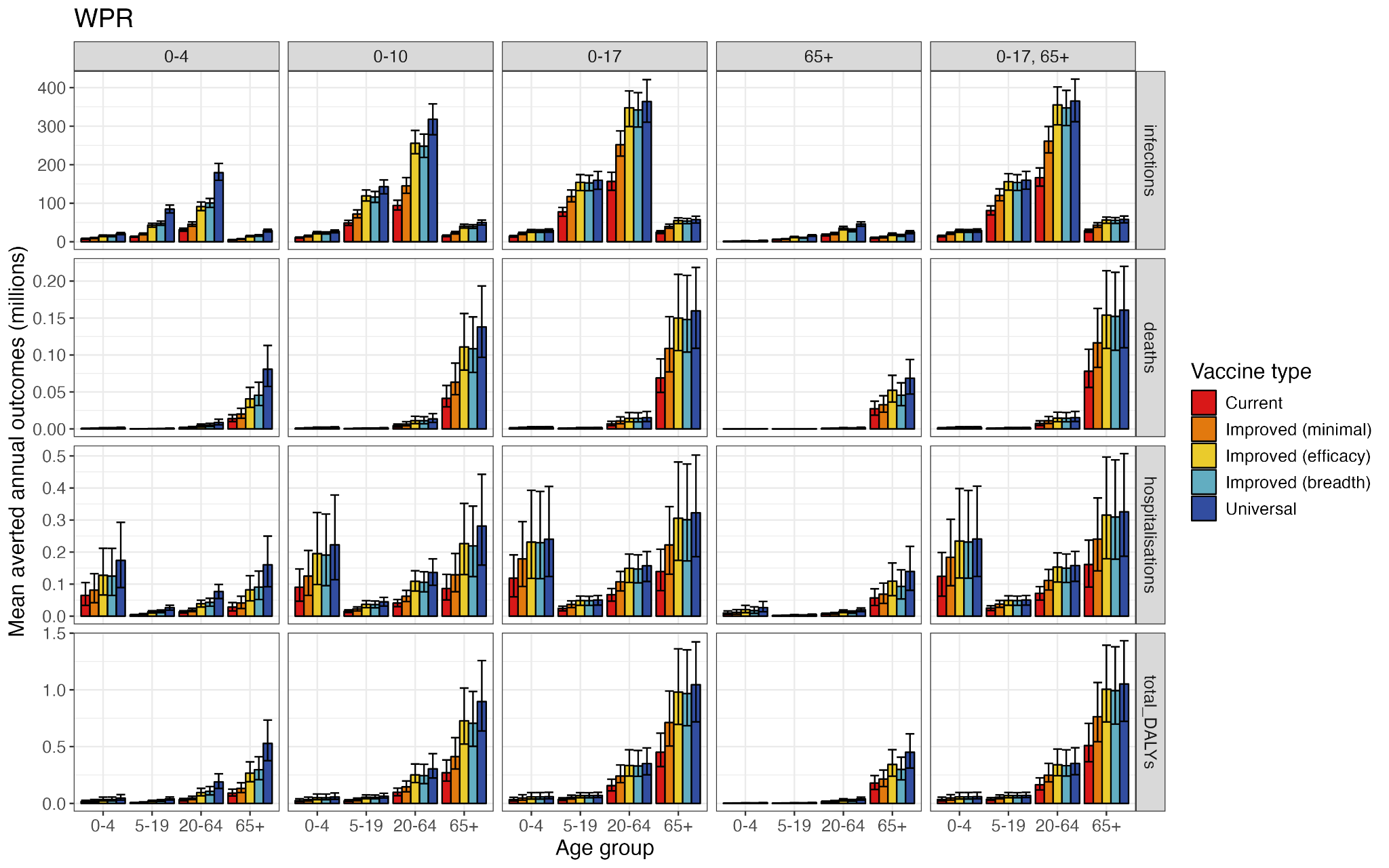

**Figure S40:** Averted annual age-specific health outcomes under each age-targeting strategy and vaccine type, in the Western Pacific Region.

### 8. Threshold prices

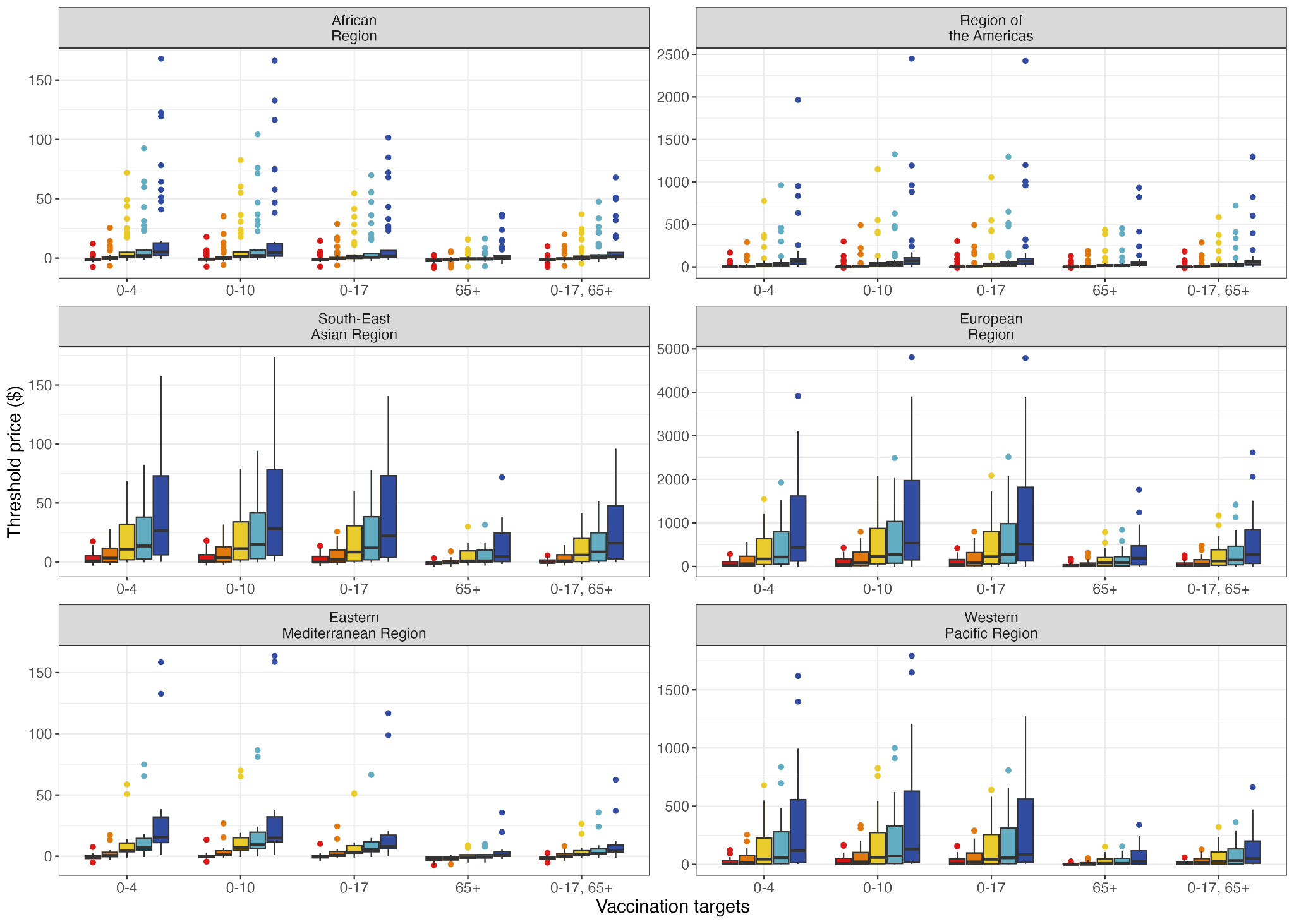

**Figure S41:** Median national threshold vaccine prices in each WHO Region, for each vaccine type and age-targeting strategy.

|  | | **Annual outcomes averted per 100,000 population**  **(median and 95% uncertainty interval)** | | | |
| --- | --- | --- | --- | --- | --- |
| **WHO region** | **Vaccine type** | **Infections** | **Cases (symptomatic)** | **Hospitalisations** | **Deaths** |
| AFR | Current | 9880 (4030, 15100), 21600 (13200-31100) | 6530 (2790, 10700), 14700 (8690-21400) | 11.7 (4.65, 22.7), 31.6 (12-69.3) | 2.36 (0.782, 5.41),  6.2 (2.88-11.4) |
| AFR | Improved (minimal) | 13700 (5470, 21500), 25600 (15200-37900) | 9370 (3700, 14400), 17500 (10400-26100) | 16 (7.41, 31.9), 39.4 (17.4-87.3) | 3.12 (1.01, 8.88),  8.37 (3.7-15.3) |
| AFR | Improved (efficacy) | 18300 (5560, 29200), 37300 (22000-58900) | 12300 (3770, 20700), 25100 (14600-40800) | 20.5 (7.94, 43.4), 52.7 (25.7-103) | 4.55 (1.84, 11),  11.8 (4.58-25.4) |
| AFR | Improved (breadth) | 17900 (5500, 28800), 36800 (20500-52400) | 12400 (3760, 20400), 25300 (13700-35900) | 20.4 (7.82, 42.5), 52.1 (24.6-96.7) | 4.56 (1.84, 11),  11.2 (4.32-24.3) |
| AFR | Universal | 20000 (5560, 31300), 45300 (27300-72000) | 13400 (3770, 21600), 30500 (18100-47900) | 22.2 (7.96, 48.2), 64.1 (31.5-120) | 5.02 (1.86, 12),  15 (5.57-32.8) |
| AMR | Current | 5870 (3810, 8990), 30000 (18400-40500) | 3930 (2600, 6090), 20300 (12300-29700) | 8.36 (4.59, 16.7), 39.8 (20.3-71.7) | 0.357 (0.163, 0.813), 7.76 (4.63-13.1) |
| AMR | Improved (minimal) | 9910 (7310, 14700), 36600 (24100-48200) | 6690 (4690, 10300), 24600 (16400-35600) | 14.7 (7.93, 26.1), 47.7 (24.9-86.7) | 0.637 (0.284, 1.3), 12.5 (7.58-21.1) |
| AMR | Improved (efficacy) | 18400 (14200, 25100), 52600 (40000-65600) | 12500 (8980, 17500), 35300 (26600-48900) | 27.2 (15.5, 45.8), 68.4 (36.4-124) | 1.21 (0.575, 2.36), 24.2 (14-39.3) |
| AMR | Improved (breadth) | 17200 (13100, 22400), 50500 (37800-63600) | 11600 (8330, 15600), 33600 (25200-45700) | 25.3 (14.4, 41.6), 66.1 (35-115) | 1.09 (0.525, 2.11), 21.4 (13.1-32.5) |
| AMR | Universal | 24200 (18500, 31100), 62500 (49600-76600) | 16200 (11800, 21400), 41900 (32400-56200) | 35.2 (20.1, 57.1), 81 (42.8-140) | 1.58 (0.758, 3.01), 32.2 (19.1-51.2) |
| EMR | Current | 11900 (5490, 17400), 20500 (11300-27900) | 7850 (3910, 12300), 13600 (7560-19100) | 12.2 (6.43, 20.2), 26.8 (14.1-45.2) | 0.176 (0.0627, 0.55), 5.29 (1.48-13) |
| EMR | Improved (minimal) | 14000 (7810, 19600), 24400 (13500-33400) | 9440 (5410, 13500), 16200 (8920-23700) | 13.9 (7.26, 23.2), 33.9 (15.7-59.5) | 0.202 (0.0722, 0.644), 7.42 (2.58-17.6) |
| EMR | Improved (efficacy) | 19900 (11900, 27700), 35200 (20000-51400) | 13500 (8120, 18900), 23800 (13300-35800) | 19.9 (9.99, 33.3), 50.6 (25.1-94.6) | 0.297 (0.106, 0.934), 12.6 (4.62-26.3) |
| EMR | Improved (breadth) | 18400 (11100, 26100), 35600 (19800-51500) | 12700 (7520, 17800), 24000 (13400-35400) | 18.2 (8.78, 32.4), 51.5 (25.8-99) | 0.277 (0.098, 0.862), 11.9 (4.36-24.4) |
| EMR | Universal | 22800 (13800, 32500), 42000 (23900-63800) | 15600 (9440, 22000), 28000 (16600-44100) | 22.3 (10.9, 40.1), 57.3 (26.5-115) | 0.345 (0.121, 1.07), 15.8 (5.59-33.3) |
| EUR | Current | 7710 (3310, 12700), 19500 (11300-27900) | 5110 (2100, 7940), 13100 (7900-19400) | 10.5 (4.82, 23.4), 26 (14.5-45.9) | 0.595 (0.186, 1.38), 6.54 (1.66-15.7) |
| EUR | Improved (minimal) | 11300 (5610, 17500), 23500 (12600-34200) | 7580 (3550, 11400), 15800 (8990-23500) | 15.2 (7.89, 30.8), 31.8 (16.9-54.3) | 0.716 (0.223, 1.74), 9.65 (2.79-23.7) |
| EUR | Improved (efficacy) | 19600 (10800, 29200), 33800 (17500-47500) | 13300 (7040, 19100), 22700 (12200-33100) | 26.9 (13.9, 49.8), 45 (23.9-83.7) | 1.02 (0.301, 2.47), 15.7 (4.8-33.5) |
| EUR | Improved (breadth) | 18600 (11000, 26300), 35000 (18600-49600) | 12200 (7230, 18000), 23600 (12500-34400) | 25.8 (13.5, 44), 46.2 (23.3-87.6) | 1.03 (0.325, 2.46), 15.4 (4.93-34.9) |
| EUR | Universal | 24400 (14400, 35700), 39100 (21000-58700) | 16600 (9740, 23600), 26300 (13500-41900) | 33.6 (16.9, 61.7), 51.7 (25.4-98.7) | 1.25 (0.395, 2.9),  19.8 (6.4-46) |
| SEAR | Current | 11800 (6650, 15800), 22500 (12500-30800) | 7930 (4330, 10300), 14900 (8120-21000) | 12.4 (5.68, 23.6), 26.3 (12.5-49) | 1.13 (0.333, 3.3),  4.93 (1.92-11) |
| SEAR | Improved (minimal) | 18700 (10300, 24400), 28400 (16400-38200) | 12400 (6720, 16500), 19000 (10500-26200) | 19.8 (9.38, 37), 32.8 (16.5-61.6) | 1.86 (0.575, 5.25),  6.8 (2.74-14.8) |
| SEAR | Improved (efficacy) | 28000 (17500, 36600), 35000 (21100-46600) | 18800 (11200, 25000), 23400 (13400-31600) | 29.8 (14.3, 52.9), 40.9 (21.6-74.3) | 2.92 (0.974, 8.26), 8.96 (3.58-20.6) |
| SEAR | Improved (breadth) | 27500 (17600, 36700), 34400 (21100-44900) | 18600 (11200, 25200), 22700 (13400-30800) | 29.3 (14.3, 53), 40 (20.1-72.4) | 2.73 (0.983, 8.04), 8.89 (3.57-20.6) |
| SEAR | Universal | 29800 (17900, 39500), 35800 (21500-47200) | 20100 (11400, 26700), 24200 (13600-32100) | 31.5 (15.4, 56.8), 42 (21.9-76.6) | 3.28 (1.14, 9.33),  9.54 (3.63-21) |
| WPR | Current | 10800 (6040, 14100), 23500 (13800-32200) | 7140 (3890, 9410), 15700 (8650-21800) | 12.8 (6.04, 23.2), 29.5 (14.7-55.8) | 0.999 (0.418, 1.98), 6.63 (3.25-12.9) |
| WPR | Improved (minimal) | 16600 (9250, 21400), 30000 (16700-40100) | 11000 (5980, 14600), 20300 (11000-27000) | 19.7 (9.09, 35.3), 44.9 (25.2-79.7) | 1.29 (0.536, 2.72), 8.45 (4.05-15.7) |
| WPR | Improved (efficacy) | 23700 (14800, 31000), 52200 (45700-61800) | 15900 (9430, 21100), 35700 (29400-43500) | 27.8 (15.2, 50.6), 75.8 (43-133) | 1.6 (0.688, 3.14),  10.4 (4.86-17.9) |
| WPR | Improved (breadth) | 23400 (14800, 30900), 47800 (43600-59000) | 15700 (9430, 21200), 32600 (27100-41300) | 27.5 (12.3, 52.3), 69.8 (41.1-120) | 1.61 (0.688, 3.12), 10.4 (4.86-17.9) |
| WPR | Universal | 25100 (15200, 33500), 63200 (56600-74900) | 17000 (9660, 22600), 42600 (35600-52000) | 29.4 (13.4, 56.3), 92.3 (53.6-157) | 1.66 (0.712, 3.19), 10.5 (4.92-18.1) |

**Table S12:** Regional minimum and maximum annual averted outcomes per 100,000 population between 2025-2029 (inclusive), under 50% vaccination coverage in under 18-year-olds (median and 95% uncertainty ranges), for each vaccine type. Range of years chosen for increased comparability with current population sizes.

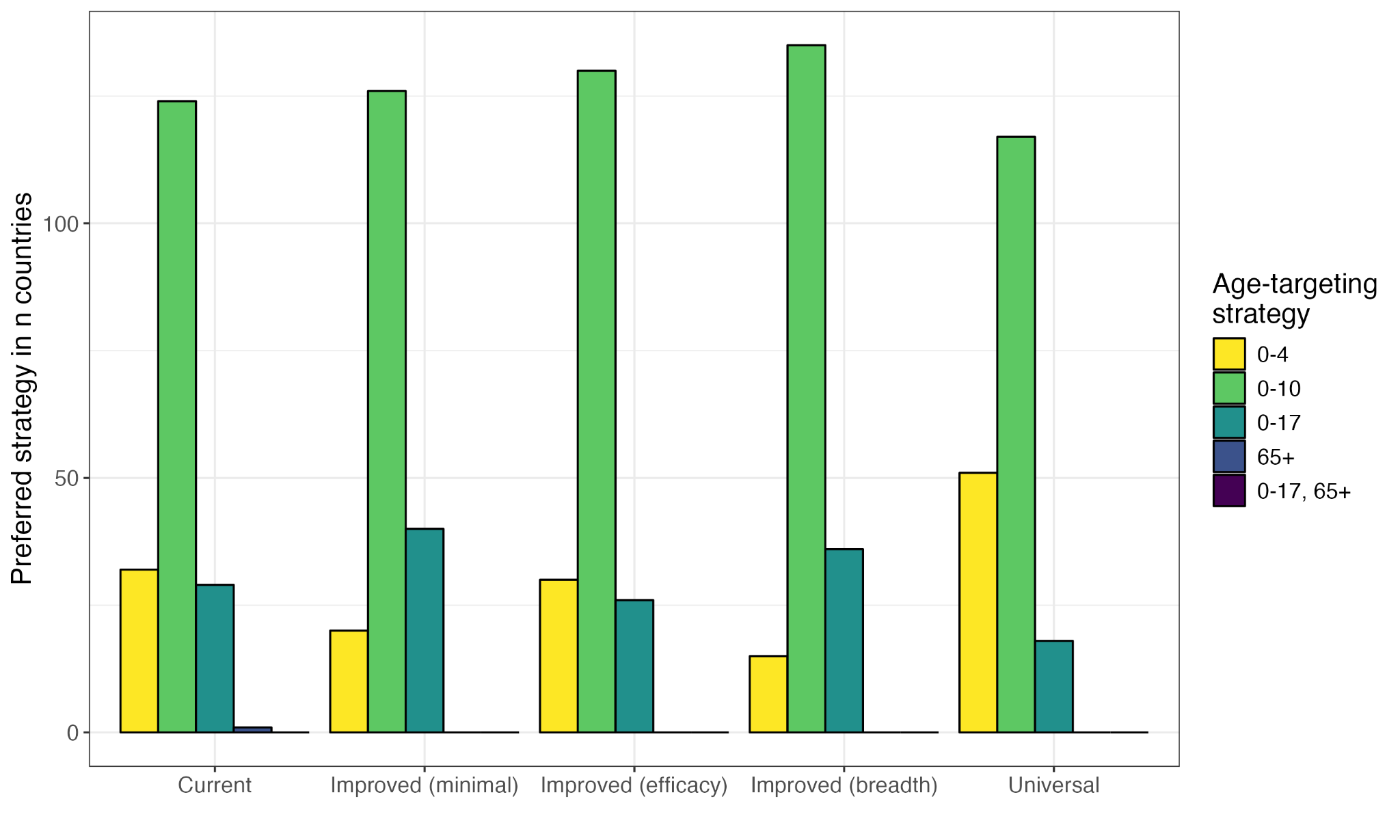

**Figure S42:** Number of countries in which each age-targeting strategy has the highest median threshold price, under each vaccine type.

| **Income group** | **Age-targeting strategy** | **Vaccine type** | **Minimum price ($)** | **Maximum price ($)** | **Countries cost-effective** |
| --- | --- | --- | --- | --- | --- |
| Low-income countries | 0-4 | Current | -3.1 (-9.8, -0.52) | -0.028 (-1.9, 1.4) | 0% |
| Low-income countries | 0-4 | Improved (minimal) | -2.9 (-9.7, -0.12) | 0.37 (-1.6, 2.8) | 12% |
| Low-income countries | 0-4 | Improved (efficacy) | -2.2 (-9, 1.5) | 2 (-2.4, 12) | 62% |
| Low-income countries | 0-4 | Improved (breadth) | -1.9 (-8.8, 2.1) | 2.9 (-1.9, 15) | 77% |
| Low-income countries | 0-4 | Universal | -0.57 (-7.9, 5.1) | 6 (-0.091, 27) | 92% |
| Low-income countries | 0-10 | Current | -3.1 (-9.8, -0.51) | -0.085 (-2, 1.1) | 0% |
| Low-income countries | 0-10 | Improved (minimal) | -2.9 (-9.6, -0.05) | 0.38 (-1.6, 2.6) | 15% |
| Low-income countries | 0-10 | Improved (efficacy) | -2.2 (-9, 1.6) | 1.9 (-2.3, 11) | 62% |
| Low-income countries | 0-10 | Improved (breadth) | -1.8 (-8.7, 2.4) | 3 (-1.8, 15) | 77% |
| Low-income countries | 0-10 | Universal | -0.55 (-7.2, 4.7) | 5.5 (-0.38, 23) | 92% |
| Low-income countries | 0-17 | Current | -3.2 (-9.9, -0.77) | -0.32 (-3.1, 0.21) | 0% |
| Low-income countries | 0-17 | Improved (minimal) | -3 (-9.8, -0.33) | 0.062 (-1.9, 1.5) | 4% |
| Low-income countries | 0-17 | Improved (efficacy) | -2.5 (-9.3, 0.87) | 1.2 (-0.96, 4.8) | 35% |
| Low-income countries | 0-17 | Improved (breadth) | -2.2 (-9, 1.7) | 1.9 (-0.64, 6.9) | 54% |
| Low-income countries | 0-17 | Universal | -1.6 (-8, 2) | 3.6 (0.24, 11) | 81% |
| Low-income countries | 65+ | Current | -3.4 (-10, -0.98) | -0.41 (-3.2, -0.048) | 0% |
| Low-income countries | 65+ | Improved (minimal) | -3.3 (-10, -0.92) | -0.36 (-3.1, 0.023) | 0% |
| Low-income countries | 65+ | Improved (efficacy) | -3 (-8.5, -0.91) | 0.07 (-2.2, 1.2) | 4% |
| Low-income countries | 65+ | Improved (breadth) | -3 (-8.5, -0.88) | 0.087 (-2.2, 1.2) | 4% |
| Low-income countries | 65+ | Universal | -2.7 (-8.2, -0.28) | 0.94 (-1.4, 2.8) | 38% |
| Low-income countries | 0-17, 65+ | Current | -3.3 (-10, -0.85) | -0.33 (-3.1, 0.16) | 0% |
| Low-income countries | 0-17, 65+ | Improved (minimal) | -3.1 (-9.8, -0.53) | -0.12 (-1.6, 0.89) | 0% |
| Low-income countries | 0-17, 65+ | Improved (efficacy) | -2.7 (-9.5, 0.32) | 0.8 (-1.2, 3.8) | 31% |
| Low-income countries | 0-17, 65+ | Improved (breadth) | -2.4 (-9.3, 1.1) | 1.3 (-0.87, 5.3) | 35% |
| Low-income countries | 0-17, 65+ | Universal | -1.9 (-8.2, 1.3) | 2.6 (-0.32, 8.7) | 58% |
| Lower-middle-income countries | 0-4 | Current | -7.5 (-17, -1.9) | 5.1 (-0.39, 10) | 25% |
| Lower-middle-income countries | 0-4 | Improved (minimal) | -6.5 (-16, -0.65) | 10 (3.8, 19) | 56% |
| Lower-middle-income countries | 0-4 | Improved (efficacy) | -2.1 (-13, 6.6) | 28 (17, 47) | 92% |
| Lower-middle-income countries | 0-4 | Improved (breadth) | -1.3 (-5.8, 0.88) | 35 (22, 55) | 92% |
| Lower-middle-income countries | 0-4 | Universal | -0.047 (-4.3, 3) | 68 (45, 100) | 98% |
| Lower-middle-income countries | 0-10 | Current | -7.2 (-17, -1.7) | 5.6 (0.2, 11) | 31% |
| Lower-middle-income countries | 0-10 | Improved (minimal) | -5.6 (-15, 0.52) | 12 (5.6, 22) | 67% |
| Lower-middle-income countries | 0-10 | Improved (efficacy) | -1.5 (-6, 0.57) | 32 (20, 53) | 92% |
| Lower-middle-income countries | 0-10 | Improved (breadth) | -1.2 (-5.7, 1.1) | 41 (26, 65) | 96% |
| Lower-middle-income countries | 0-10 | Universal | 0.028 (-4.2, 3.1) | 78 (38, 140) | 100% |
| Lower-middle-income countries | 0-17 | Current | -7.3 (-17, -1.8) | 5 (1.3, 11) | 25% |
| Lower-middle-income countries | 0-17 | Improved (minimal) | -6.2 (-15, -0.63) | 9.8 (3.6, 20) | 58% |
| Lower-middle-income countries | 0-17 | Improved (efficacy) | -3.5 (-14, 2.3) | 32 (13, 59) | 87% |
| Lower-middle-income countries | 0-17 | Improved (breadth) | -2.5 (-13, 4.9) | 40 (17, 71) | 94% |
| Lower-middle-income countries | 0-17 | Universal | -0.41 (-4.9, 2.1) | 76 (35, 130) | 98% |
| Lower-middle-income countries | 65+ | Current | -8.4 (-18, -3.3) | 1.8 (-4, 6.8) | 10% |
| Lower-middle-income countries | 65+ | Improved (minimal) | -8 (-18, -3) | 4.9 (-2.1, 12) | 17% |
| Lower-middle-income countries | 65+ | Improved (efficacy) | -7.1 (-17, -1.7) | 16 (1.1, 34) | 37% |
| Lower-middle-income countries | 65+ | Improved (breadth) | -6.9 (-16, -1.7) | 17 (1.4, 37) | 37% |
| Lower-middle-income countries | 65+ | Universal | -4.8 (-15, 0.25) | 38 (7.9, 79) | 81% |
| Lower-middle-income countries | 0-17, 65+ | Current | -7.4 (-17, -2) | 2.7 (0.51, 4.9) | 19% |
| Lower-middle-income countries | 0-17, 65+ | Improved (minimal) | -6.7 (-16, -1.2) | 7.5 (2.2, 16) | 38% |
| Lower-middle-income countries | 0-17, 65+ | Improved (efficacy) | -4.5 (-14, 0.92) | 22 (7.6, 42) | 77% |
| Lower-middle-income countries | 0-17, 65+ | Improved (breadth) | -3.4 (-14, 2.5) | 27 (10, 51) | 87% |
| Lower-middle-income countries | 0-17, 65+ | Universal | -1.9 (-13, 6.2) | 52 (22, 92) | 96% |
| Upper-middle-income countries | 0-4 | Current | -11 (-30, -3.6) | 56 (38, 72) | 73% |
| Upper-middle-income countries | 0-4 | Improved (minimal) | -7.6 (-27, 0.94) | 130 (90, 160) | 84% |
| Upper-middle-income countries | 0-4 | Improved (efficacy) | 1.5 (-1.4, 5.3) | 350 (260, 430) | 100% |
| Upper-middle-income countries | 0-4 | Improved (breadth) | 2.6 (-0.54, 6.9) | 420 (310, 520) | 100% |
| Upper-middle-income countries | 0-4 | Universal | 7.3 (2.8, 14) | 830 (620, 1000) | 100% |
| Upper-middle-income countries | 0-10 | Current | -13 (-31, -4.7) | 63 (43, 81) | 75% |
| Upper-middle-income countries | 0-10 | Improved (minimal) | -8.7 (-27, -0.33) | 150 (100, 180) | 84% |
| Upper-middle-income countries | 0-10 | Improved (efficacy) | 1.2 (-18, 13) | 410 (300, 510) | 100% |
| Upper-middle-income countries | 0-10 | Improved (breadth) | 3.3 (-0.17, 7.6) | 480 (360, 600) | 100% |
| Upper-middle-income countries | 0-10 | Universal | 7.9 (3.2, 15) | 960 (710, 1200) | 100% |
| Upper-middle-income countries | 0-17 | Current | -13 (-31, -5) | 64 (44, 81) | 67% |
| Upper-middle-income countries | 0-17 | Improved (minimal) | -9.6 (-28, -1.2) | 150 (100, 180) | 86% |
| Upper-middle-income countries | 0-17 | Improved (efficacy) | -0.18 (-19, 11) | 420 (300, 520) | 98% |
| Upper-middle-income countries | 0-17 | Improved (breadth) | 1.9 (-1.2, 5.5) | 480 (350, 600) | 100% |
| Upper-middle-income countries | 0-17 | Universal | 4.9 (1, 10) | 1000 (740, 1200) | 100% |
| Upper-middle-income countries | 65+ | Current | -13 (-32, -5.3) | 71 (42, 100) | 35% |
| Upper-middle-income countries | 65+ | Improved (minimal) | -11 (-31, -2.1) | 140 (89, 190) | 59% |
| Upper-middle-income countries | 65+ | Improved (efficacy) | -3.4 (-25, 7.7) | 380 (240, 510) | 86% |
| Upper-middle-income countries | 65+ | Improved (breadth) | -3.1 (-25, 7.9) | 390 (260, 520) | 86% |
| Upper-middle-income countries | 65+ | Universal | -0.57 (-2.7, 2.3) | 820 (540, 1100) | 98% |
| Upper-middle-income countries | 0-17, 65+ | Current | -13 (-32, -5.3) | 63 (41, 84) | 53% |
| Upper-middle-income countries | 0-17, 65+ | Improved (minimal) | -10 (-29, -2.2) | 140 (93, 180) | 80% |
| Upper-middle-income countries | 0-17, 65+ | Improved (efficacy) | -2.3 (-22, 7.6) | 370 (260, 470) | 96% |
| Upper-middle-income countries | 0-17, 65+ | Improved (breadth) | -0.78 (-21, 9.9) | 410 (290, 520) | 98% |
| Upper-middle-income countries | 0-17, 65+ | Universal | 2.9 (-0.27, 7.3) | 820 (580, 1000) | 100% |
| High-income countries | 0-4 | Current | -0.79 (-10, 3.8) | 280 (130, 550) | 98% |
| High-income countries | 0-4 | Improved (minimal) | 6.2 (-2.7, 13) | 560 (270, 1000) | 100% |
| High-income countries | 0-4 | Improved (efficacy) | 27 (12, 42) | 1500 (750, 2800) | 100% |
| High-income countries | 0-4 | Improved (breadth) | 33 (15, 50) | 1900 (950, 3500) | 100% |
| High-income countries | 0-4 | Universal | 64 (38, 110) | 3900 (1900, 7100) | 100% |
| High-income countries | 0-10 | Current | -2 (-11, 2.4) | 430 (200, 810) | 98% |
| High-income countries | 0-10 | Improved (minimal) | 5.2 (-3.8, 11) | 800 (380, 1500) | 100% |
| High-income countries | 0-10 | Improved (efficacy) | 26 (12, 41) | 2100 (1000, 3700) | 100% |
| High-income countries | 0-10 | Improved (breadth) | 32 (16, 49) | 2500 (1200, 4500) | 100% |
| High-income countries | 0-10 | Universal | 65 (39, 120) | 4800 (2300, 8700) | 100% |
| High-income countries | 0-17 | Current | -2.5 (-12, 1.8) | 420 (200, 790) | 98% |
| High-income countries | 0-17 | Improved (minimal) | 3.7 (-5.4, 9.9) | 800 (380, 1500) | 100% |
| High-income countries | 0-17 | Improved (efficacy) | 22 (10, 40) | 2100 (1000, 3700) | 100% |
| High-income countries | 0-17 | Improved (breadth) | 28 (14, 52) | 2500 (1200, 4600) | 100% |
| High-income countries | 0-17 | Universal | 51 (29, 94) | 4800 (2200, 8300) | 100% |
| High-income countries | 65+ | Current | -3 (-13, 3.3) | 180 (54, 380) | 77% |
| High-income countries | 65+ | Improved (minimal) | -0.64 (-4.2, 3.5) | 310 (100, 640) | 96% |
| High-income countries | 65+ | Improved (efficacy) | 4.3 (-1.2, 13) | 790 (270, 1600) | 100% |
| High-income countries | 65+ | Improved (breadth) | 4.7 (-0.67, 14) | 840 (290, 1700) | 100% |
| High-income countries | 65+ | Universal | 14 (5, 31) | 1800 (620, 3500) | 100% |
| High-income countries | 0-17, 65+ | Current | -2.8 (-12, 1.7) | 260 (96, 500) | 95% |
| High-income countries | 0-17, 65+ | Improved (minimal) | 2.8 (-6.9, 9.3) | 480 (200, 900) | 100% |
| High-income countries | 0-17, 65+ | Improved (efficacy) | 15 (8.6, 25) | 1200 (520, 2200) | 100% |
| High-income countries | 0-17, 65+ | Improved (breadth) | 19 (12, 31) | 1400 (610, 2500) | 100% |
| High-income countries | 0-17, 65+ | Universal | 35 (18, 64) | 2600 (1100, 4600) | 100% |

**Table S13:** Minimum and maximum national threshold prices in each World Bank income group, assuming 50% vaccination coverage, under each age-targeting strategy and vaccine type, and proportion of countries in which the median threshold cost is above $0.

### 9. Sensitivity analyses

#### a. Coverage levels

Increased vaccination coverage in the targeted age groups is associated with increased numbers of averted infections, hospitalisations, deaths, and DALYs (Figure S43). However, there is less marginal benefit when increasing vaccination coverage, likely because less benefit of indirect protection is seen under higher vaccination coverage. For example, while vaccinating 20% of under 18-year-olds with *current* vaccines results in 1.41 prevented infections per dose, this decreases to 0.96 and 0.78 infections under 50% and 70% vaccination coverage, respectively. The corresponding numbers of prevented infections per dose using *universal* vaccines are 16.3, 7.20, and 4.33.

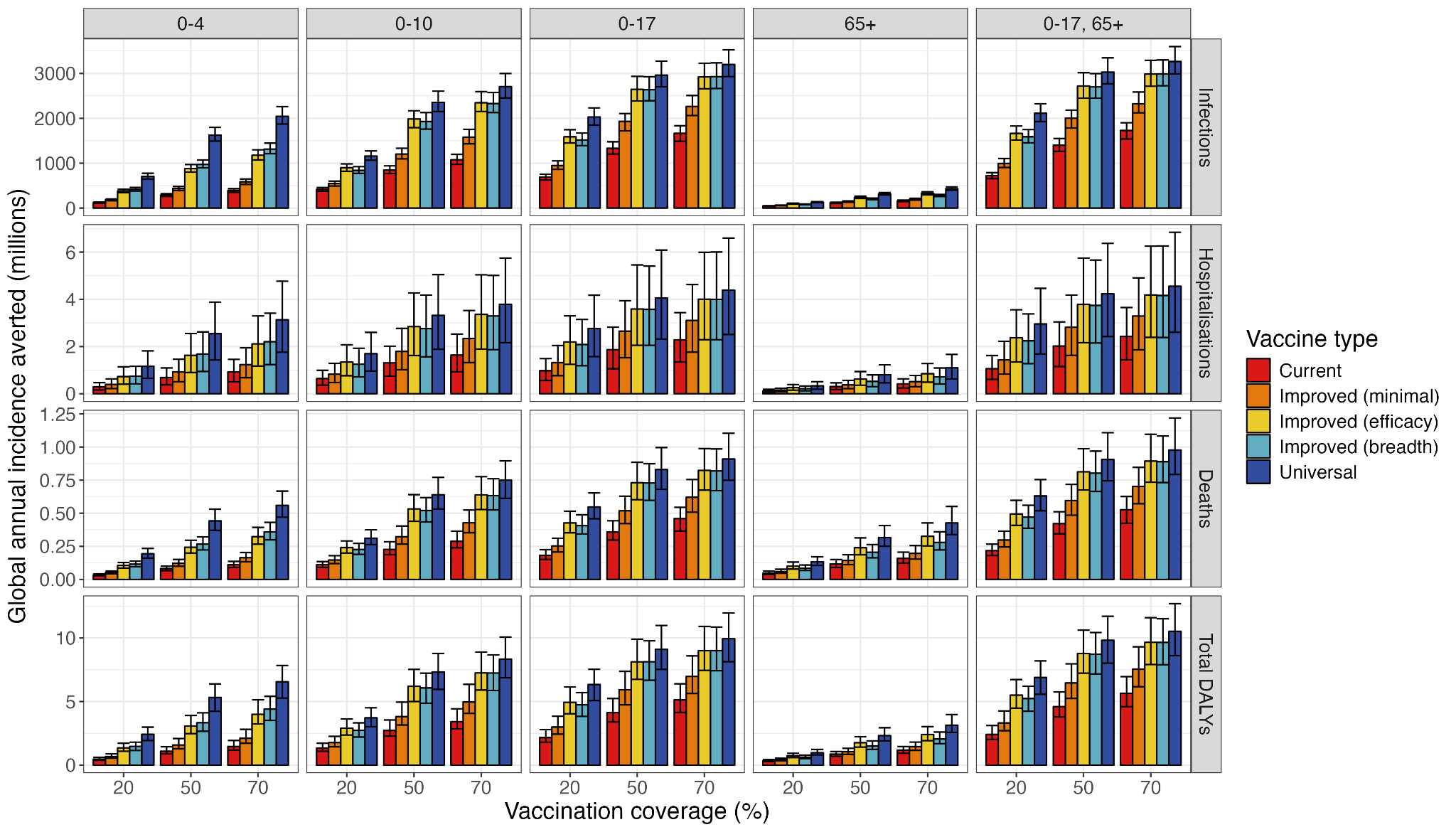

**Figure S43:** Global annual averted age-specific health outcomes under each age-targeting strategy and vaccine type, under 20%, 50%, and 70% vaccination coverage.

|  | | **Current** | **Improved (minimal)** | **Improved (efficacy)** | **Improved (breadth)** | **Universal** |
| --- | --- | --- | --- | --- | --- | --- |
| **Infections (billions)** | **20%** | 0·417 (0·376, 0·459) | 0·547 (0·492, 0·601) | 0·9 (0·82, 0·984) | 0·84 (0·774, 0·924) | 1·16 (1·07, 1·27) |
|  | **50%** | 0·849 (0·771, 0·941) | 1·20 (1·10, 1·33) | 1·99 (1·79, 2·17) | 1·93 (1·76, 2·13) | 2·36 (2·15, 2·60) |
|  | **70%** | 1·08 (0·976, 1·20) | 1·58 (1·43, 1·75) | 2·35 (2·15, 2·6) | 2·33 (2·13, 2·57) | 2·71 (2·46, 3·00) |
| **Hospitalisations (millions)** | **20%** | 0·643 (0·366, 0·995) | 0·827 (0·474, 1·29) | 1·35 (0·763, 2·07) | 1·25 (0·713, 1·93) | 1·70 (0·964, 2·60) |
|  | **50%** | 1·31 (0·744, 2·02) | 1·80 (1·02, 2·77) | 2·84 (1·62, 4·27) | 2·76 (1·56, 4·18) | 3·32 (1·88, 5·05) |
|  | **70%** | 1·64 (0·931, 2·53) | 2·34 (1·32, 3·53) | 3·36 (1·89, 5·04) | 3·30 (1·87, 5·01) | 3·79 (2·17, 5·75) |
| **Deaths (thousands)** | **20%** | 111 (90·6, 135) | 146 (120, 179) | 241 (199, 291) | 226 (189, 272) | 312 (262, 375) |
|  | **50%** | 227 (187, 285) | 323 (267, 404) | 532 (439, 641) | 520 (435, 618) | 640 (532, 771) |
|  | **70%** | 288 (239, 364) | 428 (352, 524) | 639 (528, 776) | 634 (525, 761) | 750 (613, 895) |

**Table S14:** Annual global averted infections, hospitalisations, and deaths under 20%, 50%, and 70% coverage, under the 0-10 age-targeting strategy (median and 95% uncertainty ranges).

#### b. Vaccine mechanisms

The inclusion of either reduced relative infectiousness of vaccinated individuals or disease modification for infections in vaccinated individuals only slightly increased vaccine threshold prices. This is likely due to the low number of predicted breakthrough cases; less than 10% of infections were in vaccinated individuals in all scenarios. This provides evidence that the majority of benefits of vaccination is driven by prevention of infections and the majority of costs driven by infection of unvaccinated individuals; breakthrough infections of vaccinated individuals do not significantly alter the cost-effectiveness of current or next-generation vaccines.

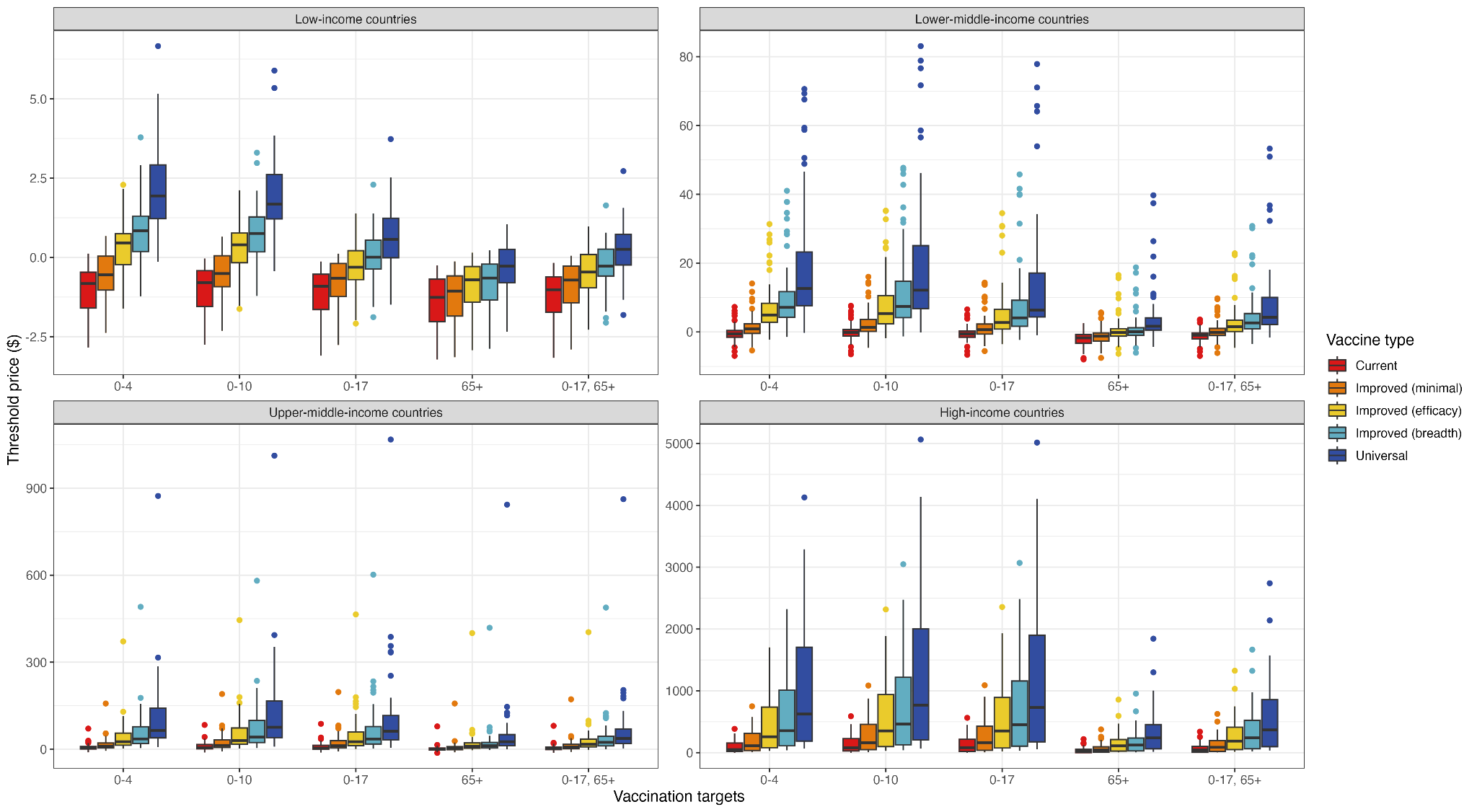

**Figure S44:** Median national threshold vaccine prices in each World Bank income group, for each vaccine type and age-targeting strategy, with reduced relative infectiousness in vaccinated individuals.

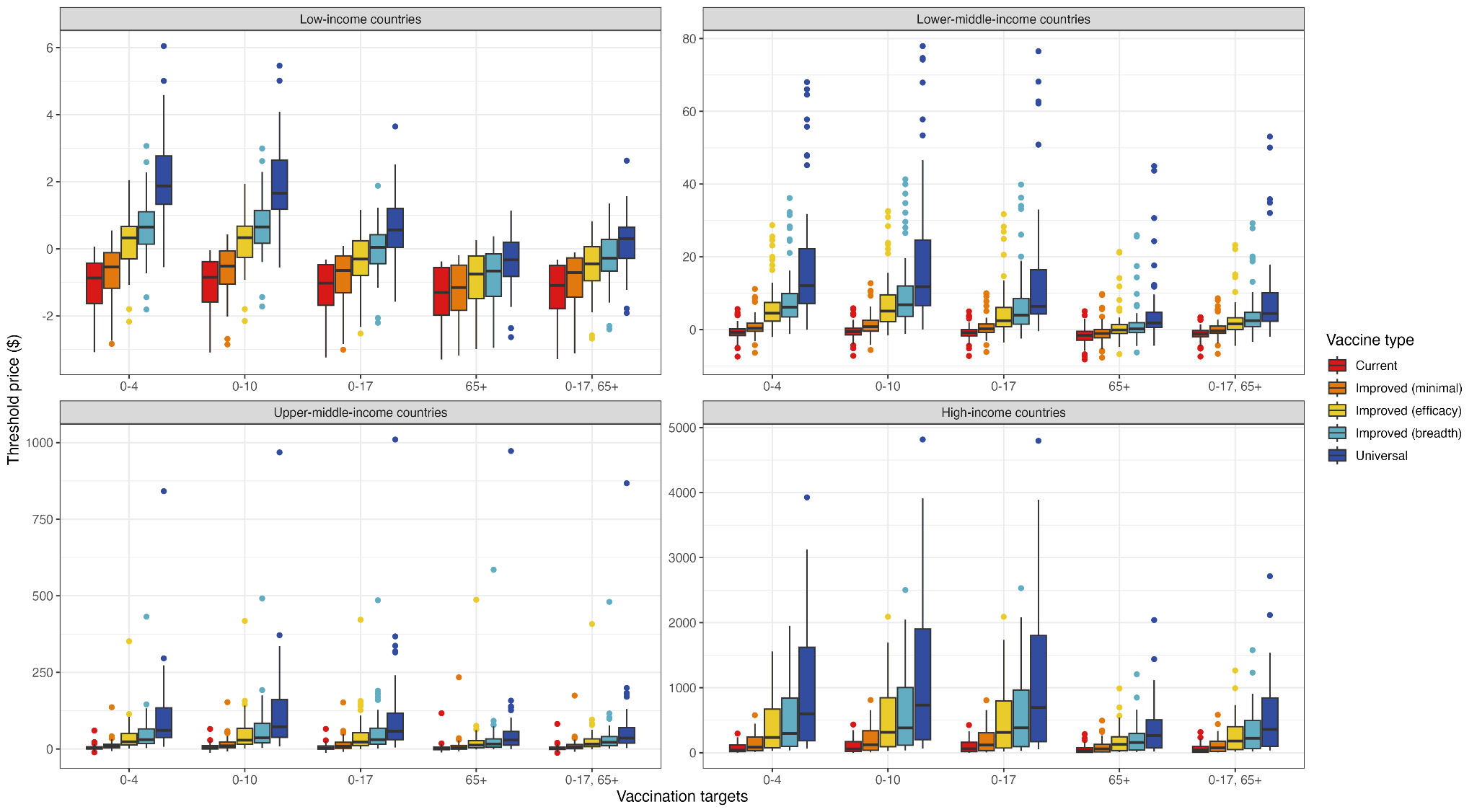

**Figure S45:** Median national threshold vaccine prices in each World Bank income group, for each vaccine type and age-targeting strategy, with disease modification in vaccinated individuals.

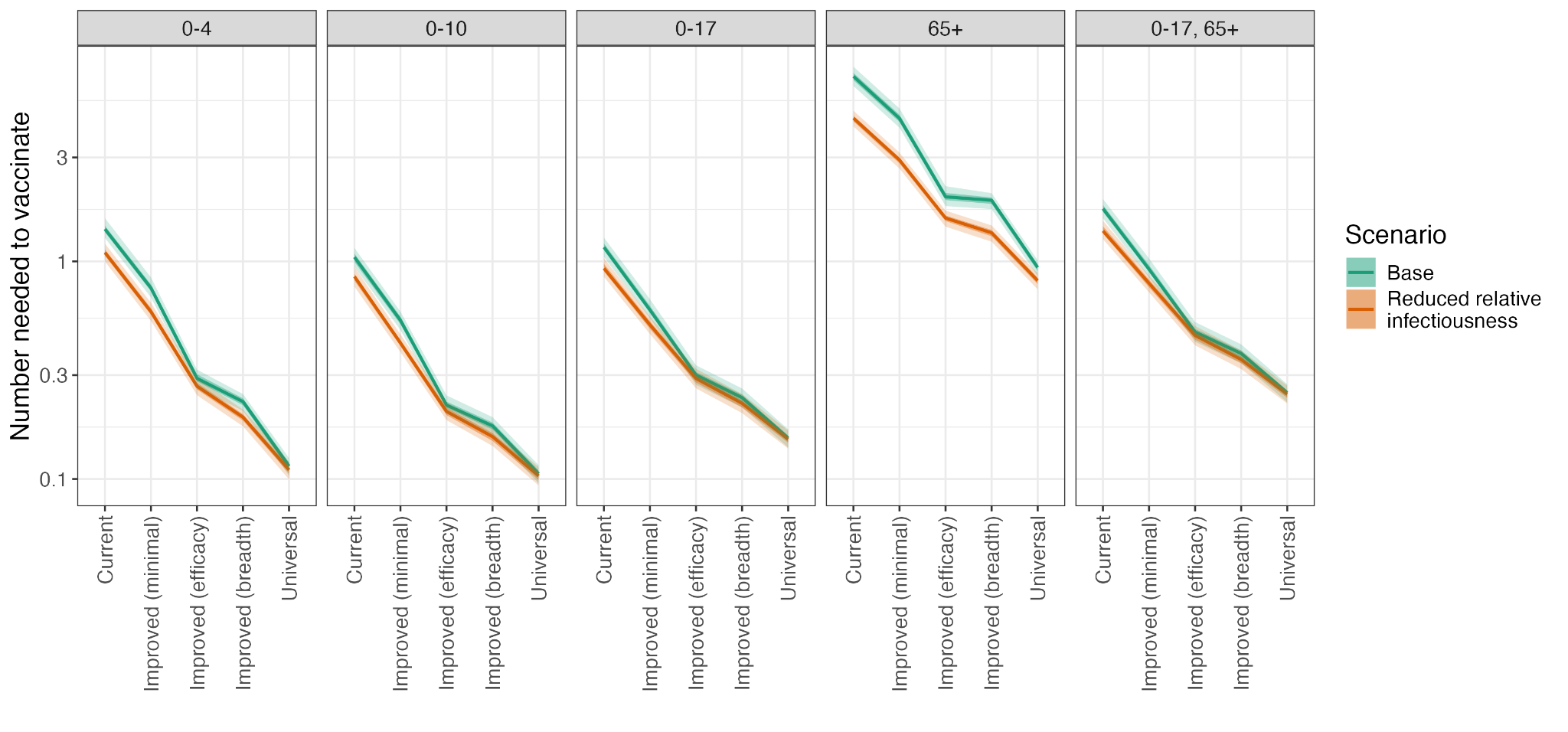

**Figure S46:** Number needed to vaccinate associated with original vaccine mechanisms and with reduced relative infectiousness of vaccinated individuals, under each age-targeting strategy and vaccine type, with 50% and 95% uncertainty intervals.

#### c. Breadth and depth

To disentangle the effects of increased vaccine efficacy and matching ability (breadth) and the length of immunity provided (depth), we ran an analysis using vaccines with the vaccine characteristics described in Table S15. This included the base case, breadth scenario (vaccines with the same mean duration of protection as *current* vaccines but improved VE), and depth scenario (vaccines with the same VE as *current* vaccines but improved duration of protection).

| **Vaccine type** | **Base** | | **Breadth** | | **Depth** | |
| --- | --- | --- | --- | --- | --- | --- |
|  | **Mean duration of protection (years)** | **Efficacy (Matched 0-64, 65+/ Mismatched 0-64, 65+)** | **Mean duration of protection (years)** | **Efficacy (Matched 0-64, 65+/ Mismatched 0-64, 65+)** | **Mean duration of protection (years)** | **Efficacy (Matched 0-64, 65+/ Mismatched 0-64, 65+)** |
| **Current seasonal vaccines** | 0.5 | 0.70, 0.46/  0.42, 0.28 | 0.5 | 0.70, 0.46/  0.42, 0.28 | 0.5 | 0.70, 0.46/  0.42, 0.28 |
| **Improved (minimal)** | 1 | 0.70, 0.46/  0.42, 0.28 | 0.5 | 0.70, 0.46/  0.42, 0.28 | 1 | 0.70, 0.46/  0.42, 0.28 |
| **Improved (efficacy)** | 2 | 0.90, 0.70/ 0.70, 0.46 | 0.5 | 0.90, 0.70/ 0.70, 0.46 | 2 | 0.70, 0.46/  0.42, 0.28 |
| **Improved (breadth)** | 3 | 0.70, 0.46/ 0.70, 0.46 | 0.5 | 0.70, 0.46/ 0.70, 0.46 | 3 | 0.70, 0.46/  0.42, 0.28 |
| **Universal vaccines** | 5 | 0.90, 0.70/ 0.90, 0.70 | 0.5 | 0.90, 0.70/ 0.90, 0.70 | 5 | 0.70, 0.46/  0.42, 0.28 |

**Table S15:** Vaccine characteristics under the base case, breath, and depth scenarios.

In order to compare these vaccine features, we compared the number needed to vaccinate to prevent one infection between the base case, breadth scenario, and depth scenario (Figure S47). By definition, current seasonal vaccines performed identically in all scenarios. As the base case vaccines benefitted from combined breadth and depth effects, they outperformed each of the breadth and depth analyses (resulting in lower NNVs).

Figure S47 shows that vaccines with increased duration of immunity (depth) outperform vaccines with increased vaccine efficacy/matching ability. These results show that duration of immunity is a key driver of the benefits of NGIVs, and that every increase in length of duration, from 6 months to 1 year or from 3 to 5 years, is associated with reduced NNV.

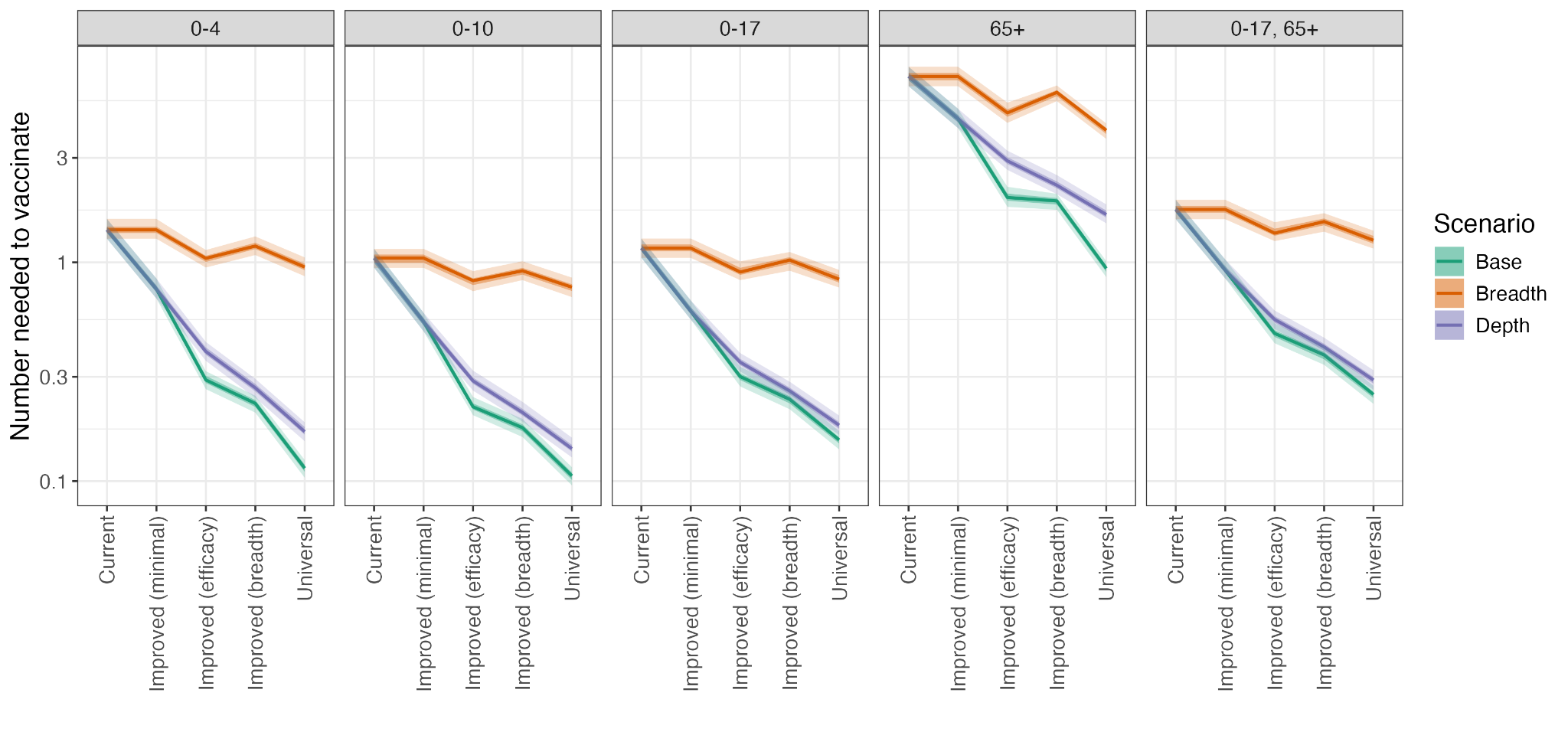

**Figure S47:** Number needed to vaccinate for each original and modified vaccine type, under each age-targeting strategy and vaccine type, with 50% and 95% uncertainty intervals.

#### d. Willingness-to-pay at 50% of GDP per capita

When the willingness-to-pay thresholds were changed to 50% of GDP per capita, we observed increased threshold prices in LICs and LMICs, and decreased threshold prices in UMICs and HICs (see Figure S48 compared to Figure 4b in the Main Text). For example, threshold costs for *universal* vaccines under the 0-10 age-targeting strategy now ranged from $0.89-$11 in LICs, but from $76-$2400 in HICs. These changes reflect the propensity for WTP thresholds in HICs to be above and LICs to be below 50% of GDP per capita (Figure S31) [63].

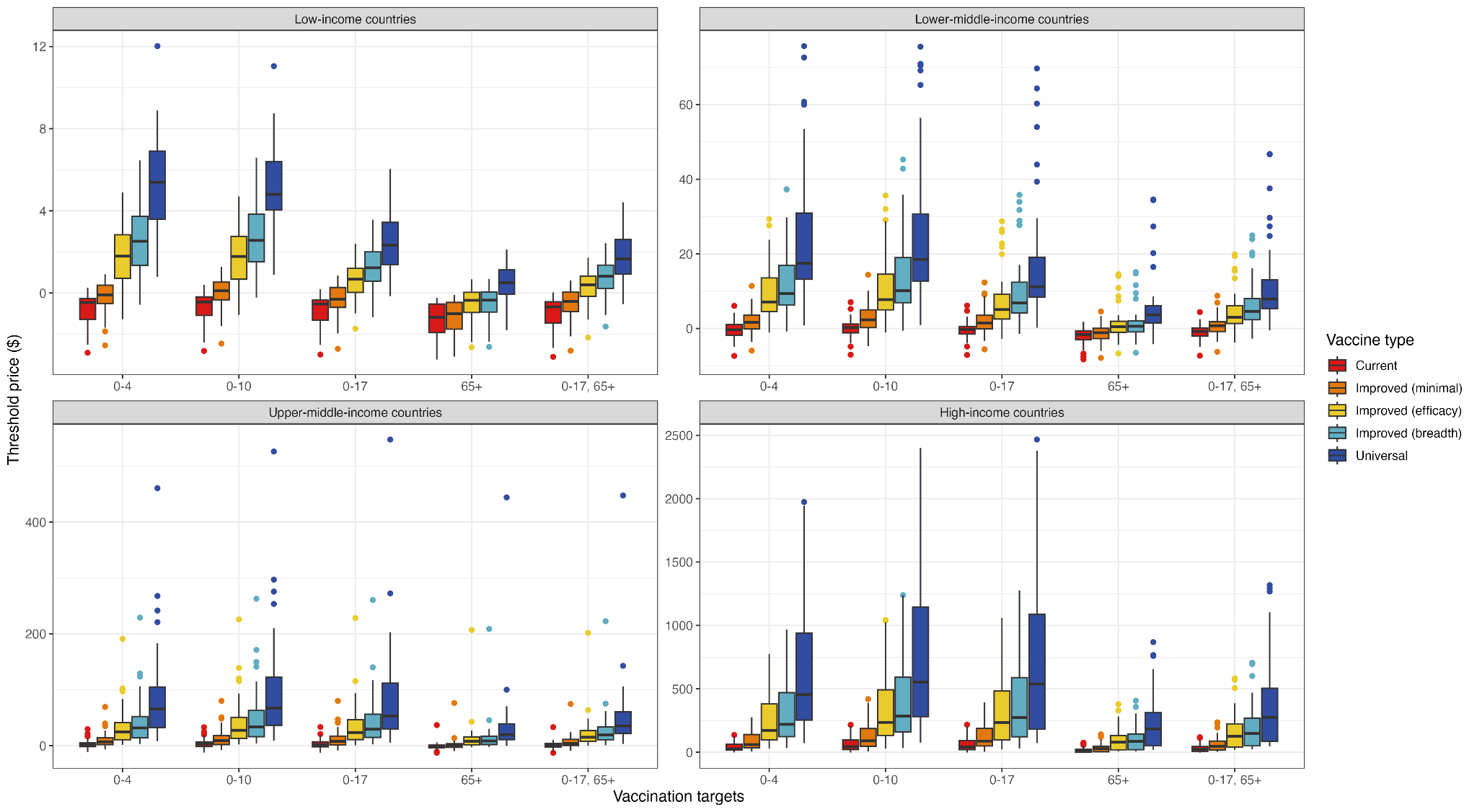

**Figure S48:** Median national threshold vaccine prices in each World Bank income group, for each vaccine type and age-targeting strategy, with willingness-to-pay thresholds set as 50% of GDP per capita.

#### e. DALY discount rate at 0%

When the discount rate of DALYs was changed from 3% to 0%, but costs still discounted at 3%, we found greatly increased threshold prices; under the 0-10 age-targeting strategy, threshold prices of universal vaccines ranged from $0.87-$11 in LICs, and $110-$9200 in HICs (see Figure S49 compared to Figure 4b in the Main Text).

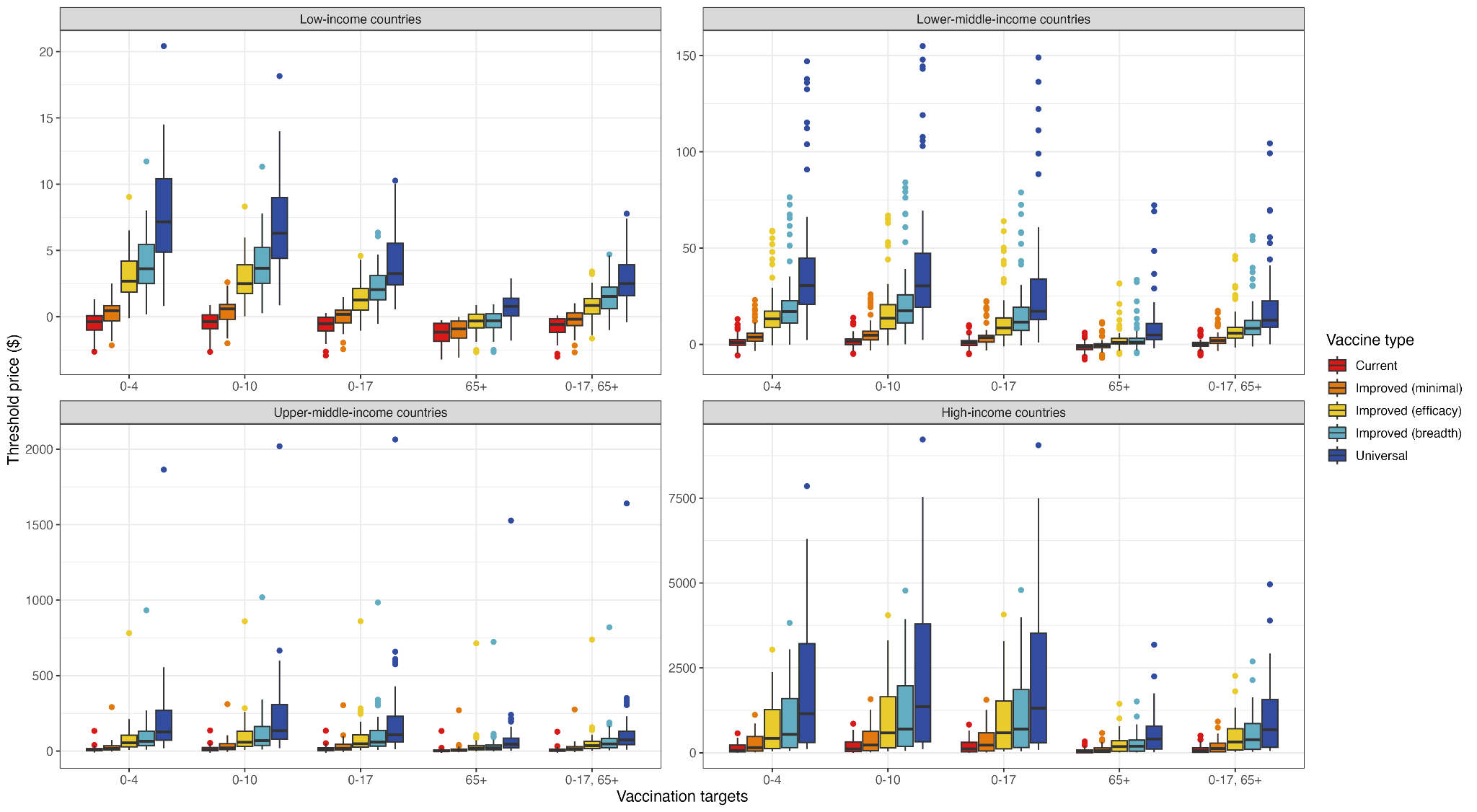

**Figure S49:** Median national threshold vaccine prices in each World Bank income group, for each vaccine type and age-targeting strategy, with discount rates for DALYs set at 0%.

#### f. Outpatient inclusion

When outpatient visits were included, we observed marginally increased threshold prices in all income groups (see Figure S50 compared to Figure 4b in the Main Text). For example, the highest threshold price of *universal* vaccines in LICs under 0-10 age-targeting increased to $6.90 (95% CI: $2.20-$19).

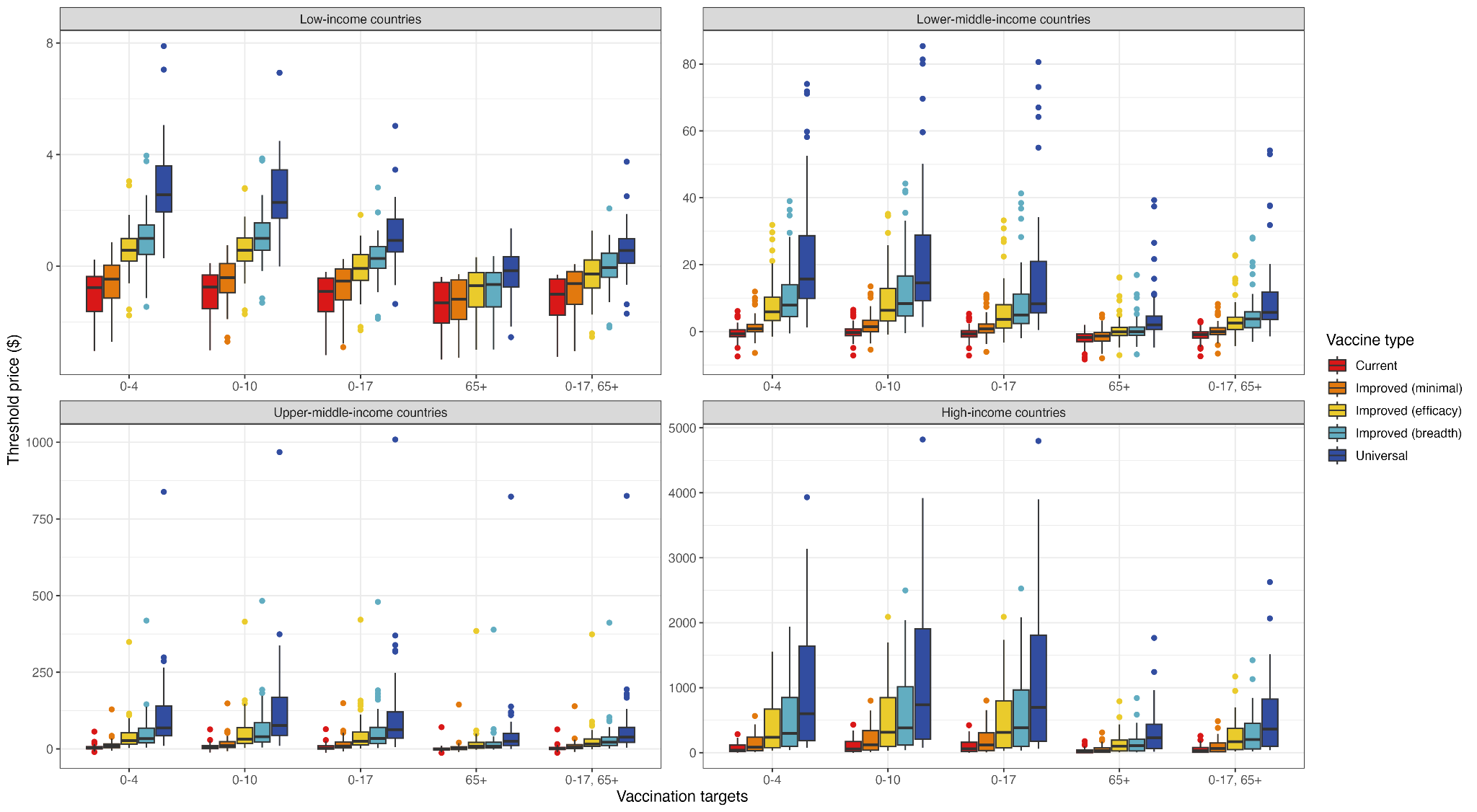

**Figure S50:** Median national threshold vaccine prices in each World Bank income group, for each vaccine type and age-targeting strategy, with the inclusion of outpatient visits and their associated costs.

#### g. Productivity costs

As a sensitivity analysis, we estimated additional economic benefit due to averted productivity losses using the human capital approach: YLLs averted (discounted over the 30-year period at a rate of 3%) multiplied by GDP per capita.

| **WHO Region** | **Vaccine type** | **Productivity costs saved ($2022, millions)** |
| --- | --- | --- |
| SEAR | Current | 46.66 (31.45, 83.96) |
| SEAR | Improved (minimal) | 68.96 (45.28, 126.63) |
| SEAR | Improved (efficacy) | 87.2 (55, 165.7) |
| SEAR | Improved (breadth) | 87.74 (55.36, 169.04) |
| SEAR | Universal | 89.44 (56, 171.63) |
| WPR | Current | 144.07 (104.13, 182.55) |
| WPR | Improved (minimal) | 228.19 (165.34, 279.33) |
| WPR | Improved (efficacy) | 316.22 (230.21, 404.45) |
| WPR | Improved (breadth) | 312.66 (225.25, 397.64) |
| WPR | Universal | 334.79 (241.07, 435.04) |
| AMR | Current | 196.58 (137.03, 291.81) |
| AMR | Improved (minimal) | 242.14 (170.93, 360.52) |
| AMR | Improved (efficacy) | 371.81 (271.95, 541.52) |
| AMR | Improved (breadth) | 356.53 (264.7, 510.93) |
| AMR | Universal | 499.97 (374.96, 725.75) |
| AFR | Current | 38.1 (25.85, 61.98) |
| AFR | Improved (minimal) | 51.52 (34.92, 83.79) |
| AFR | Improved (efficacy) | 65.48 (45.46, 106.99) |
| AFR | Improved (breadth) | 66 (45.08, 106.9) |
| AFR | Universal | 70.62 (47.96, 110.76) |
| EUR | Current | 216.06 (99.19, 401.64) |
| EUR | Improved (minimal) | 308.53 (143.08, 562.77) |
| EUR | Improved (efficacy) | 543.73 (241.57, 962.8) |
| EUR | Improved (breadth) | 524.89 (230.04, 916.53) |
| EUR | Universal | 710.54 (302.19, 1233.11) |
| EMR | Current | 16.69 (11.95, 25.55) |
| EMR | Improved (minimal) | 22.9 (16.68, 35.29) |
| EMR | Improved (efficacy) | 38.22 (27.67, 59.96) |
| EMR | Improved (breadth) | 38.94 (28.18, 61.95) |
| EMR | Universal | 48.26 (36.05, 79.81) |

**Table S16:** Productivity costs saved in 2025-2050, inclusive, by 50% vaccination coverage in individuals aged 0-17, for various influenza vaccines, in each WHO region. Costs presented in $2022, discounted at a rate of 3%.

### 10. Systematised review of estimates of seasonal influenza infection-fatality risk

#### a. Objective

In our model, we estimated seasonal influenza IFRs in each country. To provide an external source of comparison to the model results, we compared model IFR estimates against those from published influenza studies worldwide. To obtain these estimates, we conducted a systematised review of published literature reporting seasonal influenza IFR by following search and reporting procedures in the Preferred Reporting Items for Systematic Reviews and Meta-Analyses (PRISMA) statement [68,69]. Given the anticipated limited number of studies available, we extended the search to include studies of pandemic influenza A(H1N1) 2009.

As a systematised review, we applied a pre-defined and comprehensive search strategy to identify and select relevant literature but did not critically evaluate the selection strategy and assess the quality of the studies included [68].

#### b. Methods

##### Eligibility criteria

We included all full research articles that reported an IFR of seasonal influenza or pandemic influenza A(H1N1) 2009, or that contained data from which we could estimate an IFR. We excluded articles that did not report specifically on seasonal or pandemic A(H1N1) 2009 influenza, or that focused on subgroups of the population (e.g. specific age or risk groups).

##### Information sources

We searched published articles in PubMed and consulted other relevant sources identified and retrieved by the authors (e.g. citations of or references within articles we found in PubMed). We used a previously published systematic review of case-fatality risk of pandemic A (H1N1) influenza [70] as a source of relevant articles not identified by our searches. All searches were conducted on 27 June 2024.

##### Search strategy

We used the following combined search terms (applied to all fields) in the PubMed database:

(

"infection fatality ratio" OR "infection fatality rate" OR "infection fatality risk"

OR ("infection fatality" AND "proportion") OR ("fatality proportion" AND "infection")

)

OR (

("season*") AND ("asymptom*" OR "symptom*" OR "serolog*" OR "suspect*" OR "cases") AND ("infection*" OR "cases*") AND ("case fatality" OR "death*" OR "fatal*" OR "mortality") AND ("rate*" OR "risk*" OR "ratio*" OR "proportion*")

)

AND ("influenza" or "flu")

The search was limited to articles with titles and abstracts in English language (but without restrictions on the language of the main article) published between 2009 and 2024, as it was felt that the influenza A (H1N1) strain prior to 2009 may have a different IFR from the current strain.

##### Selection process

The articles identified by the search strategy were screened and evaluated by author JF to assess their eligibility against the predefined inclusion criteria. CW independently evaluated a 10% random sample of the articles identified and screened them to ensure consistency with JF's evaluations. Discrepancies between the reviewers' selected articles and evaluations were resolved through discussion and joint evaluation of the full-texts in question. No automated tools were used to search or select articles.

##### Data collection and analysis

For each article included, a PDF format file was downloaded from the publisher website, and data were extracted manually from tables, main text or figures by one of the reviewers (JF). No automation tools were used. Data were managed and analysed in R version 4.3.3 [71].

The primary data extracted were IFR point estimates and 95% confidence intervals (95% CI) for the population; as well as counts or estimates of the numbers of influenza-associated deaths (numerator) and cases (denominator), where provided. If no IFR estimates were presented, we estimated the IFR from the numerator and denominator data.

#### c. Results

##### Studies selected

Among 945 articles identified by the search terms, we shortlisted 18 after title and abstract review, of which 3 were found to be eligible studies after reviewing their full text (Figure S51). We included 7 additional studies based on references within a systematic review of case-fatality risk of pandemic influenza A (H1N1) 2009 [70], which we identified from the initial search. We found no other relevant systematic reviews in that search.

##### Study characteristics

A list of the 10 studies selected reporting IFR, including country, period, and IFR value, is presented in Table S17. One study reported IFR for seasonal influenza and 9 studies did so for the original A (H1N1) pandemic influenza strain present during 2009-2010.

##### Results of individuals studies

Forest plots of IFR estimates from the 10 studies selected are shown in Figures S51 and S52. Figure S52 compares different measurements of IFR for seasonal influenza in Hong Kong; the measurements are from successive periods of observation and two influenza strains. Figure S53 compares measurements of IFR for A (H1N1) pandemic influenza in 2009 across different countries.

##### Comparison with the model

The forest plots in Figures S50 and S51 also display the IFR estimates of the model (point estimate and 95% credible intervals) for the same countries for which an empirical IFR value was reported.

#### d. Conclusion

##### Range of the studies found

The ten studies reporting IFR that were identified through the current literature search are an uneven representation of countries across the world. The studies contain solely high-income countries (n=7) and contain no middle-income and no low-income countries; there is also unevenness within the high-income countries, with some, e.g. Hong Kong, featuring 4 times. No studies were found reporting IFR for four of the seven ITZs: Africa, Asia-Europe, North America, and Southern America.

##### Comparison with the model

We found a single study, from Hong Kong, reporting IFR for seasonal influenza [72]. For each period of observation (between early 2009 and December 2011) and for either one or both of the strains reported in this study, the confidence intervals reported contain the IFR estimate of the model for Hong Kong (Figure S52), therefore, indicating consistency with the estimate of the model. Note that in the two periods where the empirical IFR of the two strains differ, the model seasonal-influenza IFR agrees with the empirical IFR of a seasonal strain, i.e. either A (H3N2) during Jan-09-Nov-09, or A (H1N1) during Dec-09-Nov-10 after seasonal adaptation following the 2009 pandemic emergence. In Hong Kong, the reported IFR values for the original pandemic influenza A (H1N1) during 2009 [73,74] (and the earlier value in [72]) are much lower than those for seasonal influenza. For the other countries, the IFR values reported for the pandemic strain are of a similar order of magnitude between them and compared to the Hong Kong value, which suggests these pandemic strain IFR values would also be below the seasonal influenza IFR ranges estimated by the model for these countries.

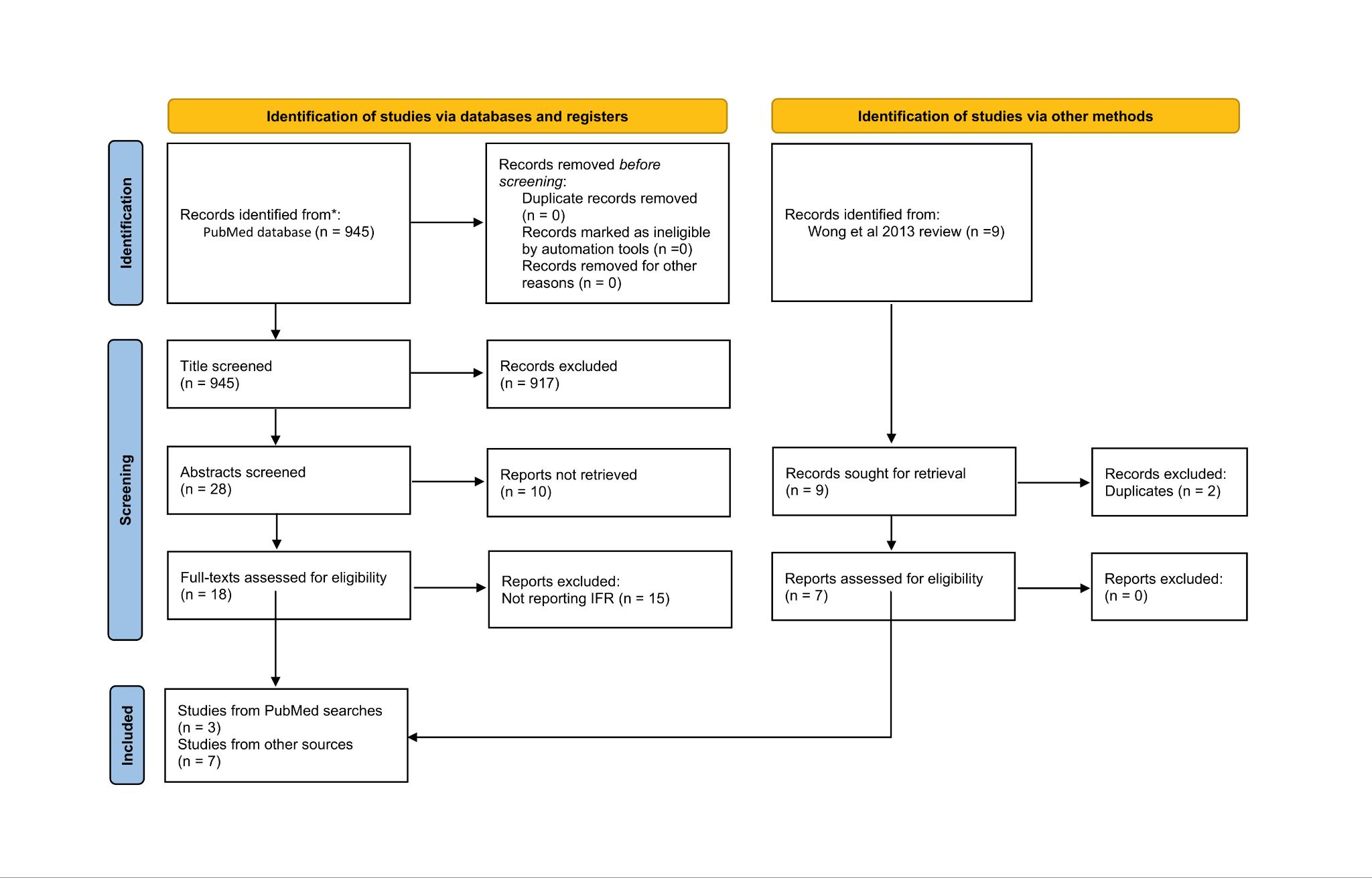

**Figure S51:** PRISMA flow diagram of the selection of studies reporting infection-fatality ratios.

| **IFR seasonal** | | **Source** | **Country** | **Period** | **IFR/100,000** | **95% CI** | **Notes** |
| --- | --- | --- | --- | --- | --- | --- | --- |
| Kwok 2017 [72] | | PubMed search | Hong-Kong | Jan-09-Nov-09 | 9.3 | 6.6-13.1 | A(H1N1)2009 |
|  |  |  |  | Dec-09-Nov-10 | 30.8 | 20.9-45.5 |  |
|  |  |  |  | Dec-10-Dec-11 | 37 | 28.1-48.8 |  |
|  |  |  |  | Jan-09-Nov-09 | 46.2 | 22.7-93.0 | A(H3N2) |
|  |  |  |  | Dec-09-Nov-10 | 78.7 | 53.5-116.0 |  |
|  |  |  |  | Dec-10-Dec-11 | 54.6 | 18.9-149.0 |  |
| **IFR A(H1N1)pdm09** | | **Source** | **Country** | **Period** | **IFR/100,000** | **95% CI[75]** | **Notes** |
| Bandaranayake 2010 | | Wong 2013 [70] | New Zealand | Apr-Sep-2009 | 4.5 | NA |  |
| Chen 2011 [76] |  | PubMed search | Taiwan | Jul-09-Aug-10 | 1 | 0.6-1.4 |  |
| McVernon 2010 [77] | | Wong 2013 [70] | Australia | Apr-Dec-2009 | 10 | NA |  |
| Presanis 2011 [78] | | PubMed search | England | Jun-Aug-2009 | 5 | 4-8 |  |
|  |  |  |  | Sep-09-Feb-10 | 9 | 4-14 |  |
| Riley 2011 [79] |  | Wong 2013 [70] | Hong-Kong | Jul-09-Feb-10 | 7.6 | 6.2-9.5 |  |
| Steens 2011[80] | | Wong 2013 [70] | Netherlands | Sep-09-Apr-10 | 4.7 | 3.2-9.2 |  |
| Sypsa 2011 [81] | | Wong 2013 [70] | Greece | Aug-09-Feb-10 | 6.3 | 5.3-7.5 |  |
| Wong 2013 [73] | | PubMed search | Hong-Kong | May-Dec-2009 | 8.2 | 0.1-17.3 | Excess deaths |
|  |  |  |  | May-Dec-2009 | 5.8 | 3.9-7.8 | Confirmed deaths |
| Wu 2010 [74] |  | Wong 2013 [70]] | Hong-Kong | Apr-Dec-2009 | 4.4 | 3.2-17 |  |

**Table S17:** Characteristics of the studies included in the review.

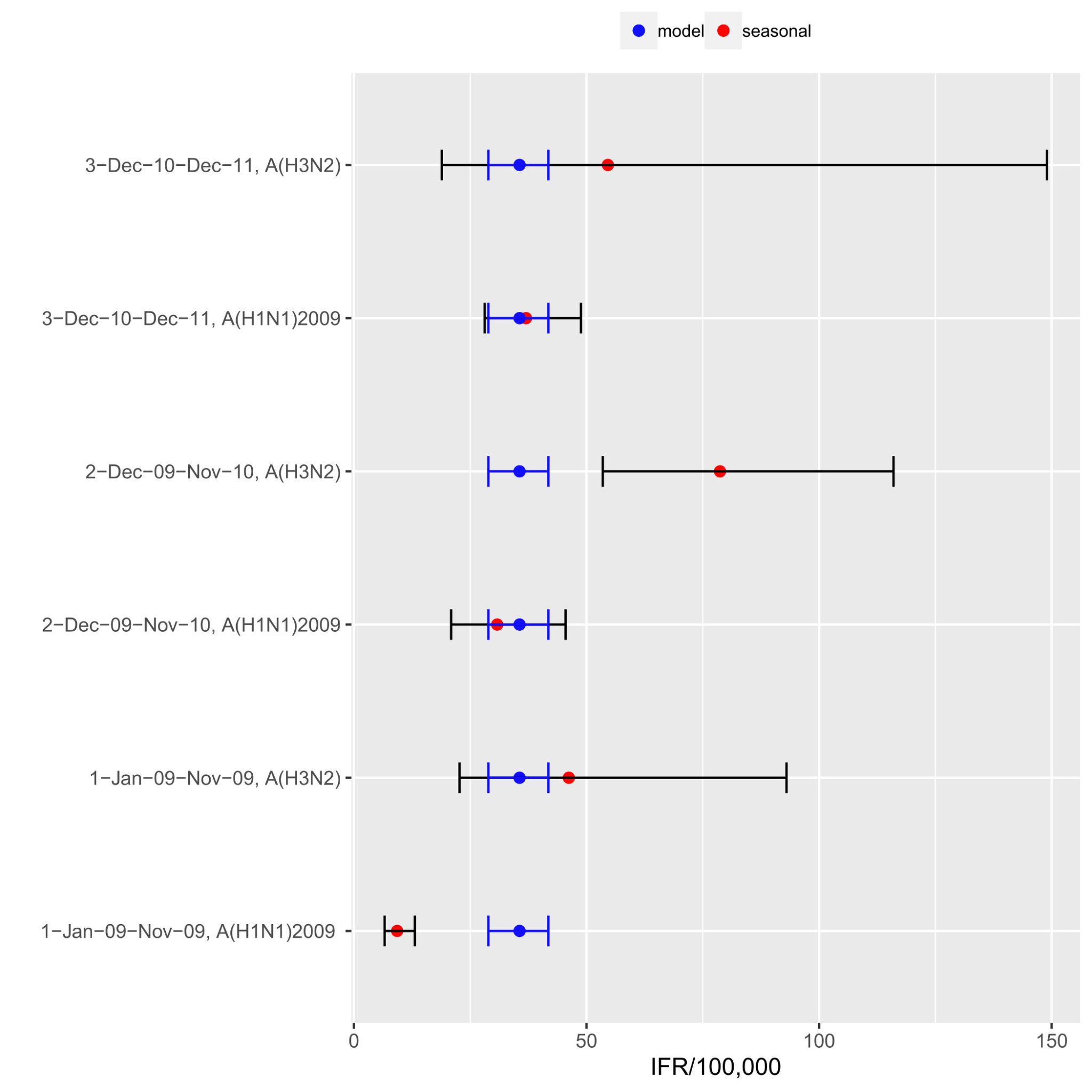

**Figure S52:** Forest plot of seasonal influenza IFR estimates from the Hong Kong study and from the model. The empirical estimates are from three different periods during 2009 through to 2011 and from two influenza strains, A(H3N2) and A(H1N1) 2009.

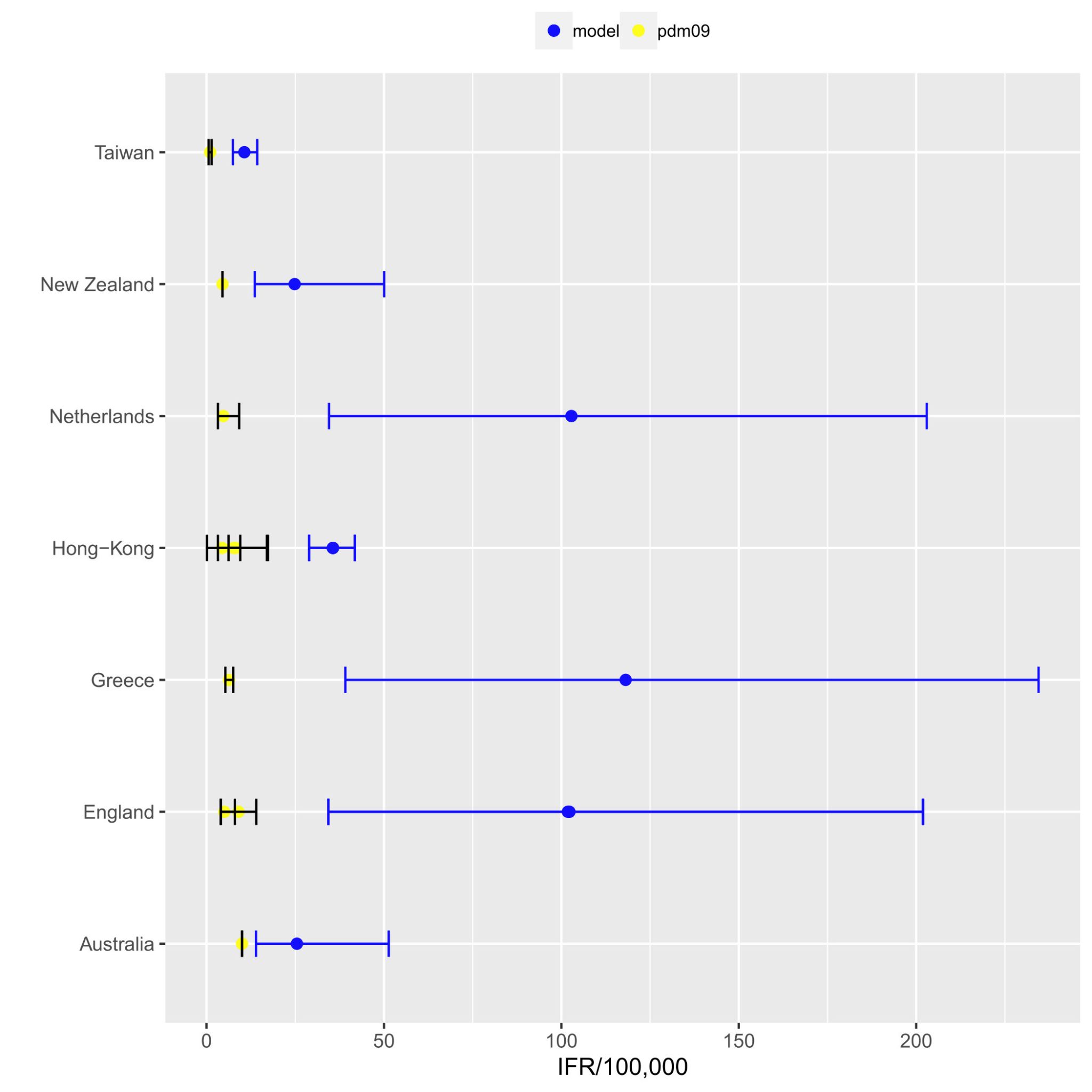

**Figure S53:** Forest plot of A(H1N1) 2009 pandemic influenza IFR estimates from empirical studies and from the seasonal influenza model.
